# A plasma-based DNA test for quantification of disease burden in acute myeloid leukemia patients undergoing bone marrow transplantation

**DOI:** 10.64898/2026.02.10.26345949

**Authors:** Yuxuan Wang, Jiajun Xie, Sergiu Pasca, Maria Popoli, Janine Ptak, Lisa Dobbyn, Natalie Silliman, Suman Paul, Richard J. Jones, Mark J. Levis, Samuel D. Curtis, Christopher Douville, Cynthia Shams, Matthew Z. Guo, Shirley Mo, Christopher D. Gocke, Sami N. Malek, Catherine M. Bollard, Chetan Bettegowda, Kenneth W. Kinzler, Bert Vogelstein, Nickolas Papadopoulos, Lukasz P. Gondek

## Abstract

Allogeneic hematopoietic cell transplantation is the only curative option for many patients with acute myeloid leukemia (AML). In the current study, we designed and implemented a personalized assay, called v96, incorporating up to 96 mutations in 30 AML patients undergoing transplantation. The assay was performed on DNA derived in cells from the bone marrow as well as in cell-free plasma. All 30 (100%) of patients harbored molecular evidence of residual leukemia during remission that was detectable by the v96 assay, while only 6 (20%) had evidence of disease as assessed by conventional clinical assays. Furthermore, cell-free DNA from plasma proved to be more sensitive than DNA from cells of the bone marrow for identifying residual leukemia. The median number of mutants was 352-fold higher in plasma taken prior to transplantation for patients who relapsed compared to those who did not relapse. At two months post-transplantation, 27 of the 30 patients still harbored detectable leukemia as assessed by the v96 assay. Twenty-two of these patients had a subsequent decrease in leukemic burden assessed by the v96 assay, usually only after immunosuppression was discontinued and supporting a graft-versus-leukemia effect. These results document the feasibility of using a relatively large panel of carefully chosen mutations and a highly specific assay as non-invasive markers of therapeutic response in AML patients, minimizing the need for multiple bone marrow biopsies.

**STATEMENT OF SIGNIFICANCE:** We report a blood test that tracks up to 96 patient-specific mutations and applied it to patients with AML who had undergone bone marrow transplantation. Using this test to evaluate cell-free plasma DNA, we found evidence of residual leukemia cells both during remission (prior to transplantation) in all patients, and two months following transplantation in 90% of patients. This test can mitigate the need for invasive bone marrow biopsies to follow patients with leukemia. Moreover, the test appears to be more accurate than standard assays for detecting residual leukemia, and has the potential to guide the timing of transplantation and subsequent therapeutic measures, thereby laying the foundation for future prospective studies.

## INTRODUCTION

Allogeneic hematopoietic cell transplantation (hereinafter denoted “transplantation”) is routinely used to treat patients with acute myeloid leukemia (AML). It is offered with curative intent for most medically fit patients with intermediate- and poor-risk disease, which comprise the majority of patients with AML (1, 2). Prior to transplantation, conditioning chemoradiotherapy is administered both to reduce the tumor burden and to prevent graft rejection, and the subsequent transplantation provides donor-derived immune cells to eliminate host leukemia cells (the graft-versus-leukemia effect, GvL) (3). Quantification of the effects of these therapeutic modalities has been hampered by the lack of broadly applicable assays to track low levels of measurable residual disease (MRD) in AML patients. The presence of MRD at the time of complete remission (CR) is associated with an increased probability of relapse after transplantation(4, 5). Nevertheless, about 30% of patients who are MRD negative using conventional assays eventually relapse after transplantation(6). MRD positivity after transplantation appears to be a more accurate prognosticator of relapse(7). Thus, dynamic MRD monitoring after transplantation is an attractive strategy for early detection of impending relapse. However, this requires frequent bone marrow aspiration, a painful and invasive procedure, limiting the frequency at which assessments can be performed.

In addition to classical histopathological review and flow cytometry, DNA can be purified from bone marrow. Such DNA has enabled sequencing-based assays for driver gene mutations in AML (8-11). Such assays are considerably more accurate than flow cytometry, but can be limited by the low mutation burden following bone marrow and the complex clonal diversity that is difficult to appreciate through evaluation of a small number of driver gene mutations (8-10). It is also not known whether biopsy of a single bone site is fully representative of the entire marrow. In the current study, we used a personalized assay, termed v96, to assess up to 96 leukemia-specific mutations in cell-free DNA from plasma. This assay can be performed as frequently as needed and without the need for bone marrow biopsies.

## RESULTS

### Patients

Thirty AML patients who underwent transplantation at Johns Hopkins between November 2020 and January of 2022 were included in this study (Materials and Methods). Twenty-nine patients underwent non-myeloablative conditioning, while one patient underwent myeloablative conditioning (AML 129). Twenty, seven, and three of the thirty patients received their transplant from haploidentical, mismatched unrelated, and fully matched unrelated donors, respectively (*SI Appendix*, Table S1). All patients received post-transplant cyclophosphamide-based graft-versus-host disease prophylaxis (12, 13). Only six of the 30 patients were MRD positive by clinical multiparameter flow cytometry (all with <1% Blasts). Baseline demographic and clinical characteristics of these patients are shown in *SI Appendix*, Table S1.

### Identification and assessment of leukemia-specific mutations prior to transplantation

An overview of the v96 assay is depicted in Fig. 1. In brief, DNA was purified from peripheral blood or bone marrow samples at diagnosis and at clinically defined CR (*SI Appendix*, Table S2). Based on whole genome sequencing of these DNA samples at a depth of 30 to 100-fold, up to 96 candidate mutations in each patient were selected and amplification primers designed for each of them. The primers for each patient were combined into a single tube and evaluated at high depth with an assay that can independently evaluate mutations on both the Watson and Crick strands using duplex sequencing. Candidate mutations that passed the criteria described in method were considered *bona fide*, and hereinafter are dubbed “mutations”. A median of 63 mutations (range: 12 to 96) per patient were thereby identified (*SI Appendix*, Table S2).

**Fig. 1:**
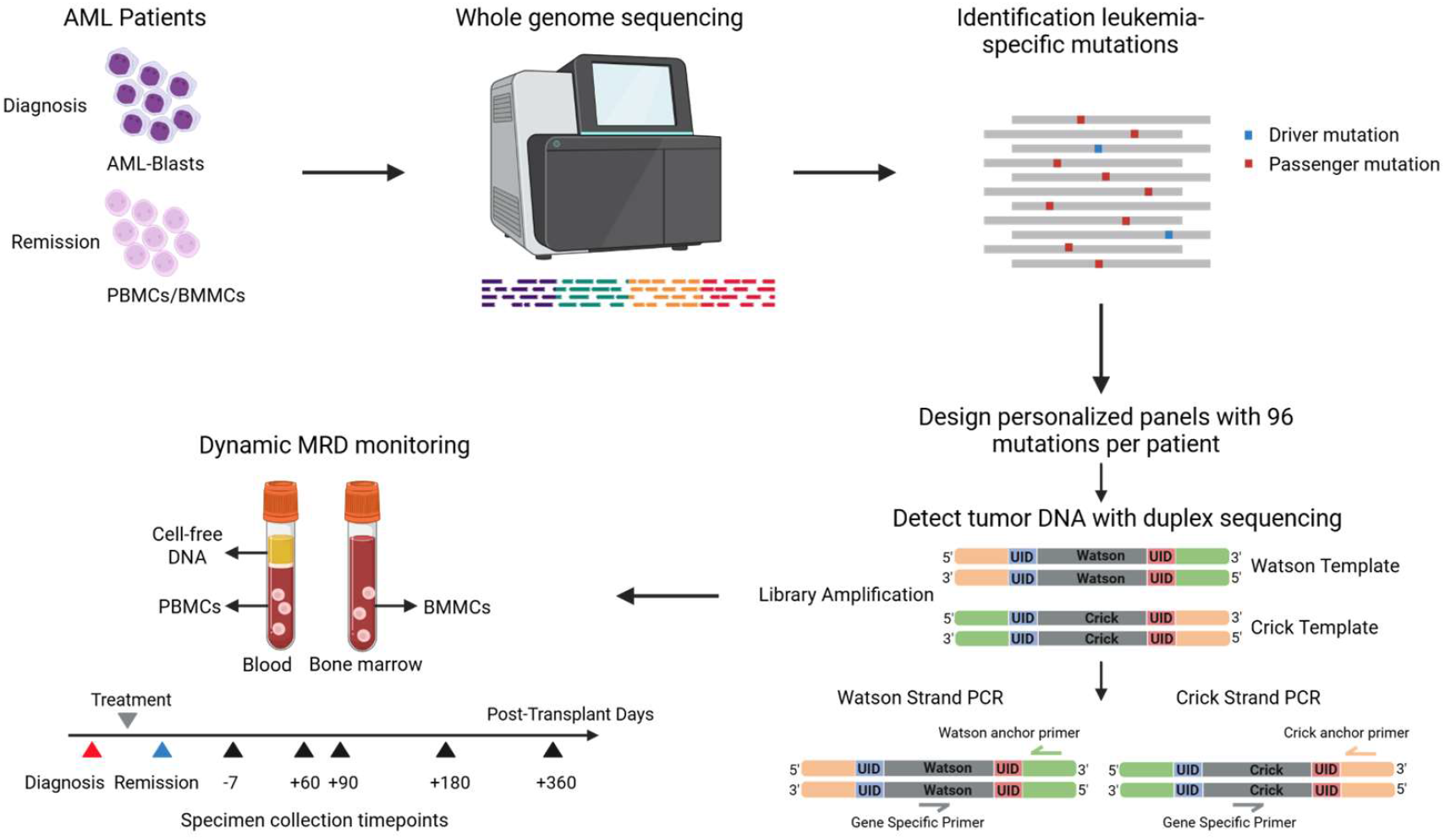
Overview of study design and the v96 assay. For each patient, whole genome sequencing was performed on DNA from bone marrow or peripheral blood at diagnosis or at complete remission (see SI Appendix, Table S2). Up to 96 mutations present at high clonal fraction in the diagnostic leukemia samples but absent or present at very low clonal fraction in the complete remission sample were subsequently selected for each patient. Primers allowing the amplification of each of the selected mutations were designed and combined into a single tube for each patient. PCR-amplification with these primers was then performed on cellular DNA or from cell-free plasma DNA collected at various time points before and after bone marrow transplantation. A mutation was scored as positive only if it was present in both the Watson and Crick strands of the same DNA molecule. PBMCs = peripheral blood mononuclear cells. BMMCs = bone marrow mononuclear cells.

CR was defined by standard clinical criteria including fewer than 5% blasts in the bone marrow, absence of circulating blasts, and absence of extramedullary disease following blood count recovery (14, 15). All 30 patients were in CR when transplanted as assessed by these clinical criteria. When cells from blood or bone marrow were assessed by flow cytometry at this time, only 20% (6 of 30) of the patients had detectable leukemic disease. In contrast, when DNA from the same cells were assessed by the v96 assay, 100% of the patients had evidence of residual leukemia (examples in Fig. 2; all data recorded in *SI Appendix* Tables S1 and S2 and Figures S1 and S2). The median number of mutant DNA molecules at CR was 281 per patient (range: 1 to 6756; *SI Appendix*, Table S2 and Fig. S1). In 21 of the 30 patients, plasma cfDNA was available after CR was obtained at 7 days prior to transplantation. In 100% of these patients (i.e., all 21 patients), evidence of residual leukemia was observed using the v96 assay in these plasma samples (*SI Appendix*, Table S2 and Figures S1 and S2). The median number of mutant DNA molecules 7 days prior to transplantation in plasma was 264 per patient (range: 1 to 84,066; *SI Appendix*, Table S2).

**Fig. 2:**
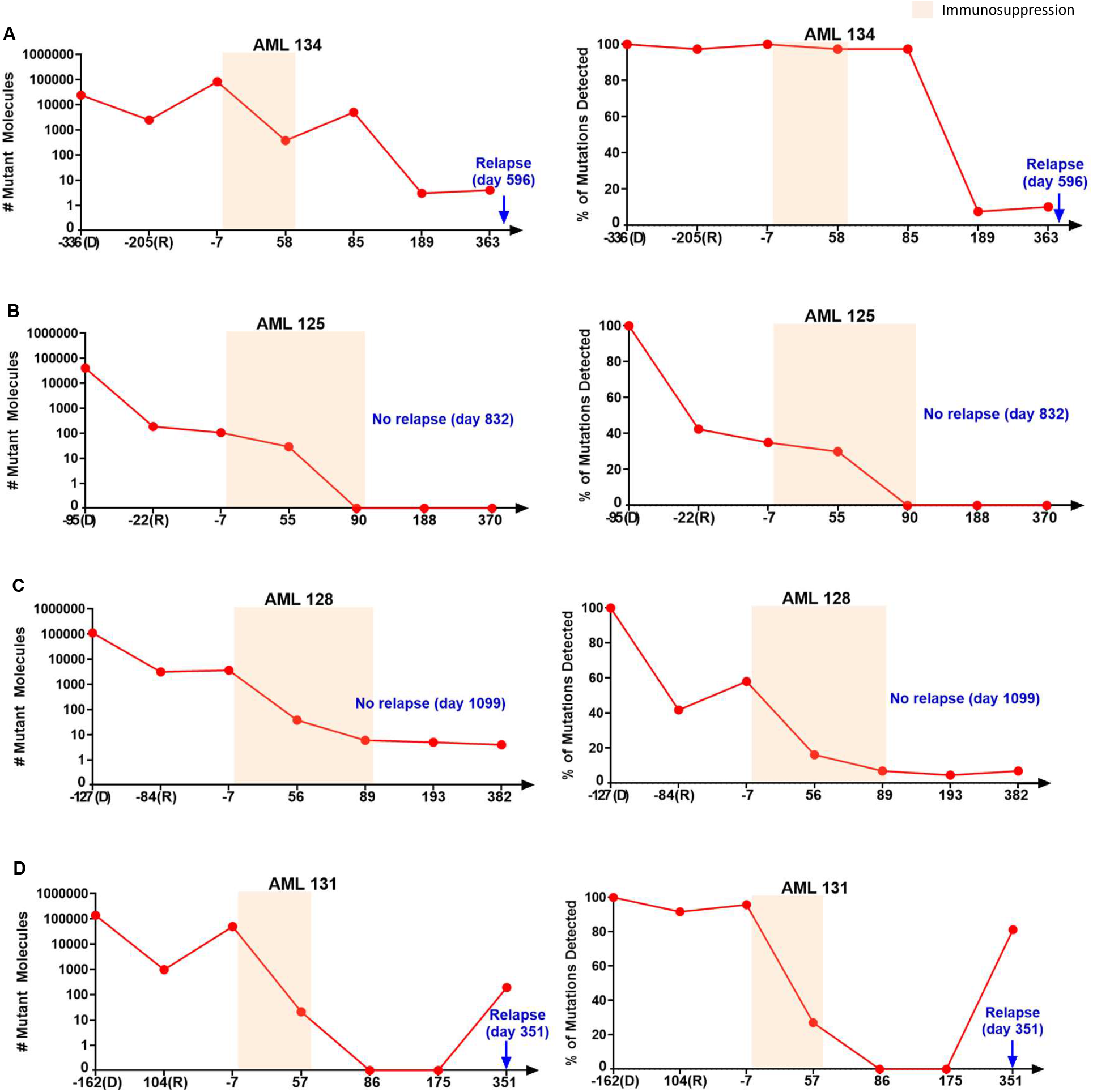
Number of mutant molecules and fraction of mutations detected before and after transplantation. Plots of number of mutant molecules detected at diagnosis (D), complete remission (R), and at other time points for four representative patients. For each patient, 150 ng of bone marrow or peripheral blood cell DNA (see SI Appendix, Table S2) was assayed by v96 for timepoints D and R, while cell-free DNA from 10-mL of plasma was assayed by v96 for the other time points. The x-axis represents the days relative to transplantation for these timepoints, with transplantation performed at day 0. Colored regions represent intervals during which immunosuppression was administered. The panels on the left represent the number of mutant molecules detected by the v96 assay. The panels on the right represent the fraction of distinct mutations found in these samples. These fractions were normalized to the number of distinct mutations detected at diagnosis (always 100%). For example, if a patient had 80 distinct mutations detected at diagnosis, and 40 of those mutations were detected at remission, the fraction on the y-axis for the remission sample would be 50%.

### Identification and assessment of leukemia-specific mutations following transplantation

Two months following transplantation, evidence for residual leukemia could still be detected in cell-free DNA from the plasma of 27 of the 30 patients using the v96 assay (examples in Fig. 2 and all data reported in *SI Appendix*, Tables S1 and S2 and Figures S1 and S2). These 27 patients included the single patient in our study that received myeloablative conditioning prior to transplantation (AML 129); all other patients received non-myeloablative conditioning. The transplantation process resulted in an median decrease of 98% in mutant molecules when the number of mutant molecules found two months after transplantation was compared to the number of mutant molecules 7 days prior to transplantation (*SI Appendix*, Table S2 and Figures S1 and S2).

Circulating DNA has a half-life of only ∼1 hour(16), so the presence of leukemia-specific mutations in plasma two or three months after transplantation provided evidence that leukemia cells were still present in the patient. The presence of even one mutant DNA molecule in 10 mL of plasma suggests that a minimum of 3,000 new leukemia cells were shedding DNA into the circulation each day (Methods). In 22 of the 27 patients with residual leukemia at two months post-transplant, the leukemia cells decreased over time (*SI Appendix*, Tables S1 and S2, Figs S1 and S2). In 13 of these 22 patients, no therapeutic agents were delivered post-transplant (*SI Appendix*, Table S1). In 12 of these 13 patients, further decreases in mutant DNA molecules were observed only *after* immunosuppressive agents for GVHD prophylaxis were discontinued (*SI Appendix*, Table S1). For example, in AML 134, the number of mutant DNA molecules at three months following transplantation was 5041, and this number decreased by 1000-fold (to 3 mutant DNA molecules) three months later (the 6-month time point in **Fig. 2A**). Another example was provided by AML 125, in whom 29 mutant DNA molecules were detected two months after transplantation, but no mutant DNA molecules were detected a month later (at the 3-month time point in **Fig. 2B**). Similar decreases were identified in other patients, such as AML 128 (**Fig. 2C**) and AML 131 (**Fig. 2D**).

### Relapse

As noted above, plasma was available 7 days prior to transplantation in 21 patients. The median number of mutant DNA molecules in plasma 7 days prior to transplantation was significantly higher in the four patients who relapsed compared to the 17 patients who did not relapse after transplantation (36,305 vs. 103; a difference of 352 fold; *SI Appendix*, Table S2). All 4 patients who relapsed had > 4000 mutant DNA molecules in plasma 7 days prior to transplantation, while only 2 of the 17 patients who did not relapse had this high level of mutant DNA molecules (p=0.0025, Fisher’s Exact Test).

Plasma was available in all 30 patients at the 2-month time point. None of the three patients without detectable mutant DNA molecules in their plasma 2 months after transplantation have relapsed (at follow-ups of 943, 1007, and 1048 days post-transplant; *SI Appendix*, Table S1 and S2). However, this low rate of relapse was not significantly different than in the 27 patients with detectable mutant DNA molecules; 7 of these 27 patients relapsed (p=1.000, Fisher’s Exact Test; *SI Appendix*, Tables S1 and S2).

In 22 patients, mutant DNA molecules *decreased* following the 2-month time point. With a median follow-up of 924 days (range 162 to 1622 days), only two of these 22 patients relapsed (*SI Appendix*, Table S1 and S2). In contrast, all five patients in whom no decrease in mutant DNA molecules was observed after the 2-month time point relapsed (p = 0.0003, Fisher’s Exact Test; *SI Appendix*, Table S1 and S2). In the seven patients who relapsed (AML 130, 131, 132, 133, 134, 236, and 247), the increase in mutant molecules preceded clinical relapse by an average of 123 days (Range 0 to 511 days; *SI Appendix*, Fig. S1 and S2).

### Comparison of DNA from bone marrow with that of plasma

DNA was available from the bone marrow as well as plasma in 15 patients during remission and prior to transplantation. These patients had a total of 643 detectable mutations in either plasma or bone marrow (**Fig. 3A**, and *SI Appendix*, Table S3). The fraction of mutant molecules was substantially higher in DNA from the plasma compared to that from the bone marrow in most cases (average MAF 2.9% vs. 0.42%; p<0.0001, two-tailed paired Student’s t-test; **Fig. 3A** and SI Appendix, Table S3). Part of the increase in mutations observed in the plasma at remission might have been due to the fact that these plasma samples were collected seven days prior to transplantation, while the remission bone marrow samples were collected earlier, at the time CR was clinically achieved (median interval between bone marrow and plasma sampling of 92 days, range 9 to 198 days, *SI Appendix*, Table S2). However, DNA samples from both plasma and bone marrow were available at the same time point, 3 months following transplantation, in a different, but overlapping set of 15 patients. Three of these 15 patients had no mutations detected at this time in either the bone marrow or plasma DNA, but the other 12 had a total of 223 detectable mutations in either the plasma or bone marrow (**Fig. 3B**, and SI Appendix, **Table S3**). As with the samples prior to transplantation, the fraction of mutant molecules was higher in DNA from the plasma compared to that from the marrow (average 0.056% vs. 0.005%; p<0.0001, two-tailed paired Student’s t-test). At the 3 month time point, three patients (AML 123, 129, and 241) were found to have mutations in their plasma (totaling 10 distinct mutations) but not in their bone marrow, while one patient (AML 131) was found to have a single mutant molecule in their bone marrow but no mutant molecules in their plasma.

**Fig. 3:**
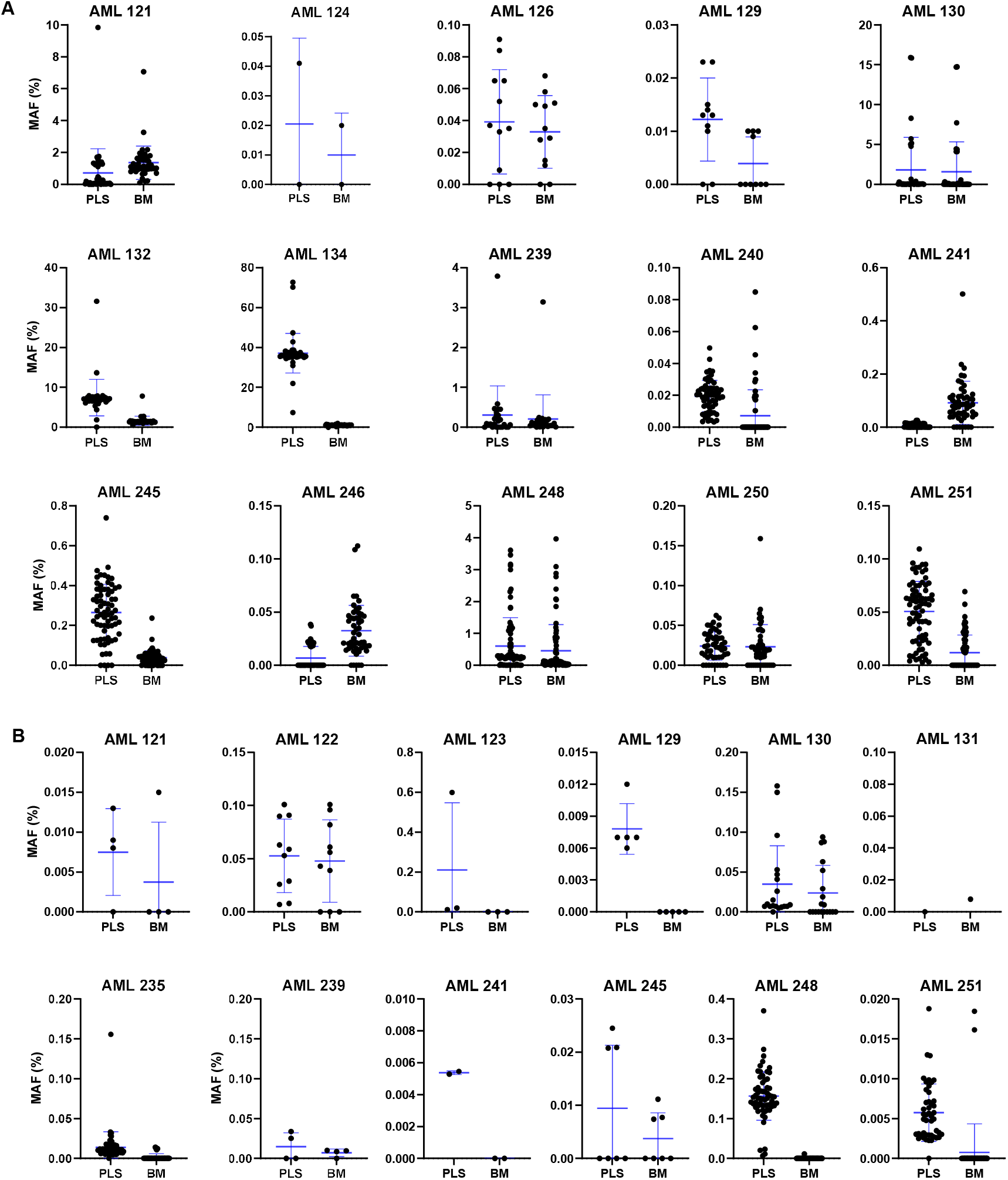
Mutant allele fraction of mutations identified in matched bone marrow and plasma samples. Eight hundred and sixty-six mutations were positive in either the bone marrow or plasma sample in 15 patients during remission prior to transplantation (A), or in 12 patients at month 3 post-transplantation (B). For those patients, the mutant allele frequencies (MAFs), defined as the ratio of mutant molecules to the total numbers of molecules evaluated in the sample, are plotted. Blue bars indicate represent +/-standard deviation.

### The v96 assay vs driver gene mutation assays

Driver gene mutations are defined as those conferring a selective growth advantage to the leukemia cells (9). The v96 assay focuses on passenger gene mutations rather than driver gene mutations because the algorithms used to select mutations eliminate those that have higher artifactual background rates (such as certain transition mutations, which are reasonably common among driver gene mutations and can lead to false positive results). Accordingly, most of the mutations (1770 of 1790, 99.4%) assessed with the v96 assay were passengers – i.e., markers of the major leukemia clone(s) at diagnostic presentation rather than functionally important genes. However, though not used for assessing MRD, we also evaluated all known driver gene mutations identified in these patients using whole genome sequencing or targeted sequencing at every time point in every patient (*SI Appendix*, Table S4; Methods).

To track driver gene mutations not included in v96 assays, we used the same basic technology employed in v96 – SaferSeqS, which evaluates both strands of DNA to achieve high sensitivity and technical specificity (<5 x 10^-7^ background mutations per bp) (17). One to four driver gene mutations were identified in each patient, and personalized SaferSeqS assays were designed for each of them (*SI Appendix*, Table S4). Major differences were observed between the results of the v96 assays and the driver gene mutation assays. In the samples drawn 7 days prior to transplantation (month 0), all 21 patients evaluated (100%) showed evidence of residual leukemia cells with the v96 assay, while a driver gene mutation was detected in only 10 of the 21 patients (48%; *SI Appendix*, Table S2 and **Figs. 4A and 5**). The median number of mutant molecules detected in the v96 assay was 264, compared to zero in the driver gene mutation assay (*SI Appendix*, Table S2 and **Figs. 4A and 5**). A total of 183,017 mutant molecules were detected in the 21 patients with the v96 assay, compared to 8,887 mutant molecules in the driver gene mutation assay – a difference of 20.6-fold (*SI Appendix*, Table S2). Of note, two of the five patients who relapsed (AML 130 and 132) had no driver mutations detected at remission prior to transplantation, while the v96 assay detected high numbers or mutations (6756 and 11 mutant molecules, respectively).

**Fig. 4:**
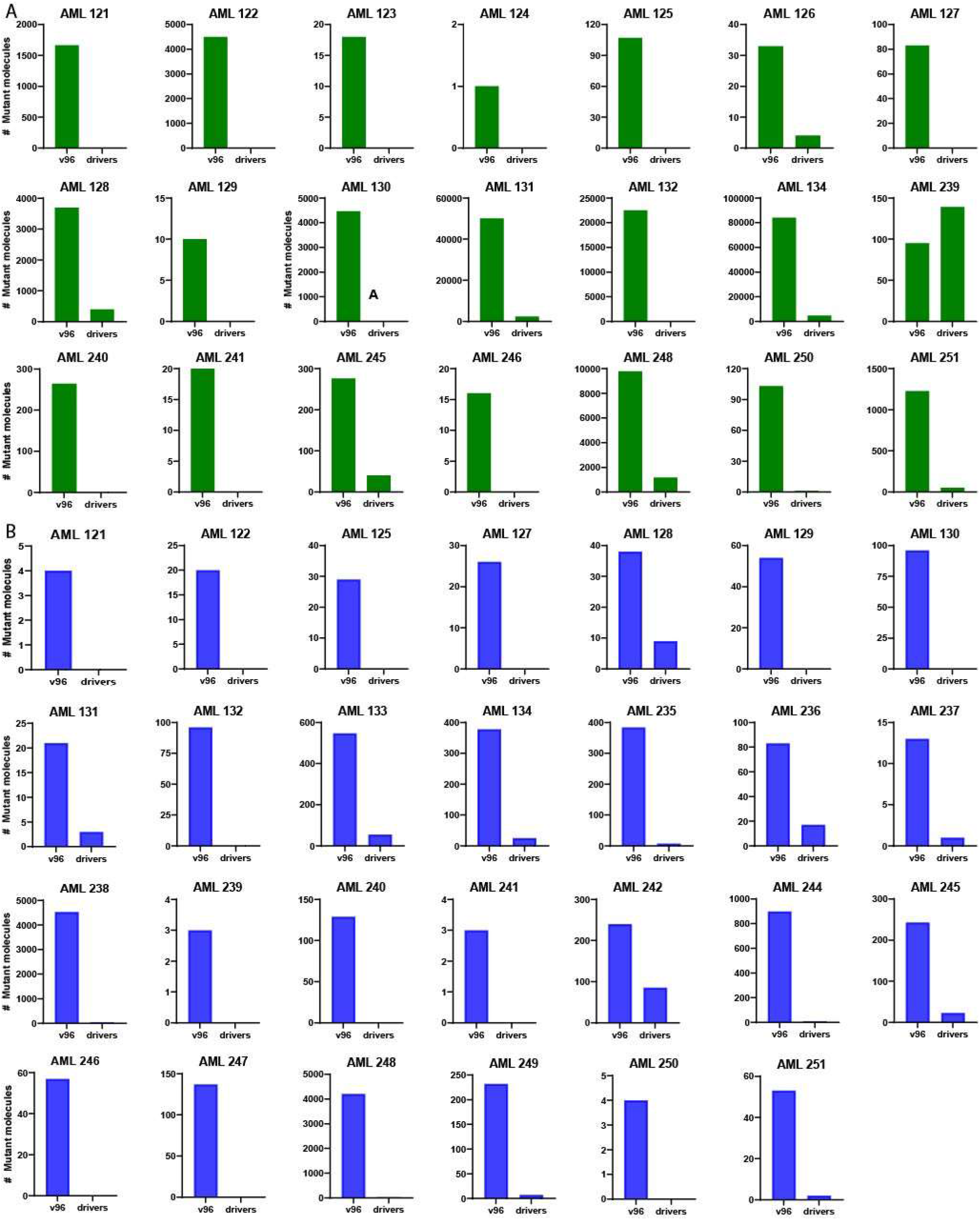
Number of mutant molecules in plasma detected through v96 or through analysis of driver mutations for each patient. Plotted are the total number of mutant molecules detected with the mutations assayed by v96 or with driver gene mutations when assayed during remission (7 days prior to transplantation; A) or at 2 months following transplantation (B). In all cases, the DNA from 10-mL of cell-free plasma was assessed. Plasma samples were available in 21 patients at 7 days prior to transplantation and in all 30 patients at 2 months following transplantation. Three of the 30 patients (AML 123, 124, and 126) had no detectable mutant molecules in plasma in either the mutations assessed by v96 or in driver gene mutations at 2 months following transplantation and are not depicted in this figure, but the corresponding values are included in SI Appendix, Table S2 and Figures S1 and S2.

Similarly significant differences between the assays of passengers vs. drivers were observed at all time points assessed. At the 2-month time point following transplantation, 27 of the 30 patients (90%) had evidence of residual leukemia in the v96 assay, while only 15 of the 30 patients (50%) had evidence of driver gene mutations (*SI Appendix*, **Table S2, Figs. 4B and 5**). This difference was also reflected in the number of mutant molecules detected (**Fig. 4B and 5**). The 3-month, 6-month, and 12-month post-transplantation timepoints also showed major differences between v96 and the driver gene mutation assays, though as the leukemia cells were eradicated, the number of mutations in the v96 assay concordantly decreased (*SI Appendix*, **Table S2 and Fig. 5**). Importantly, at no time point in any patient was a driver gene mutation detected in a sample when the corresponding v96 assay was negative, with one exception: at the 3 month time point for AML 131, one mutant molecule was detected in a driver gene (*JAK2* p.V617F), while none was detected with v96 (SI Appendix, **Table S2**). This particular mutation was also positive in a healthy plasma control, potentially indicating a high background at this position (see Methods), which is the reason it was not included in the v96 panel.

**Fig. 5:**
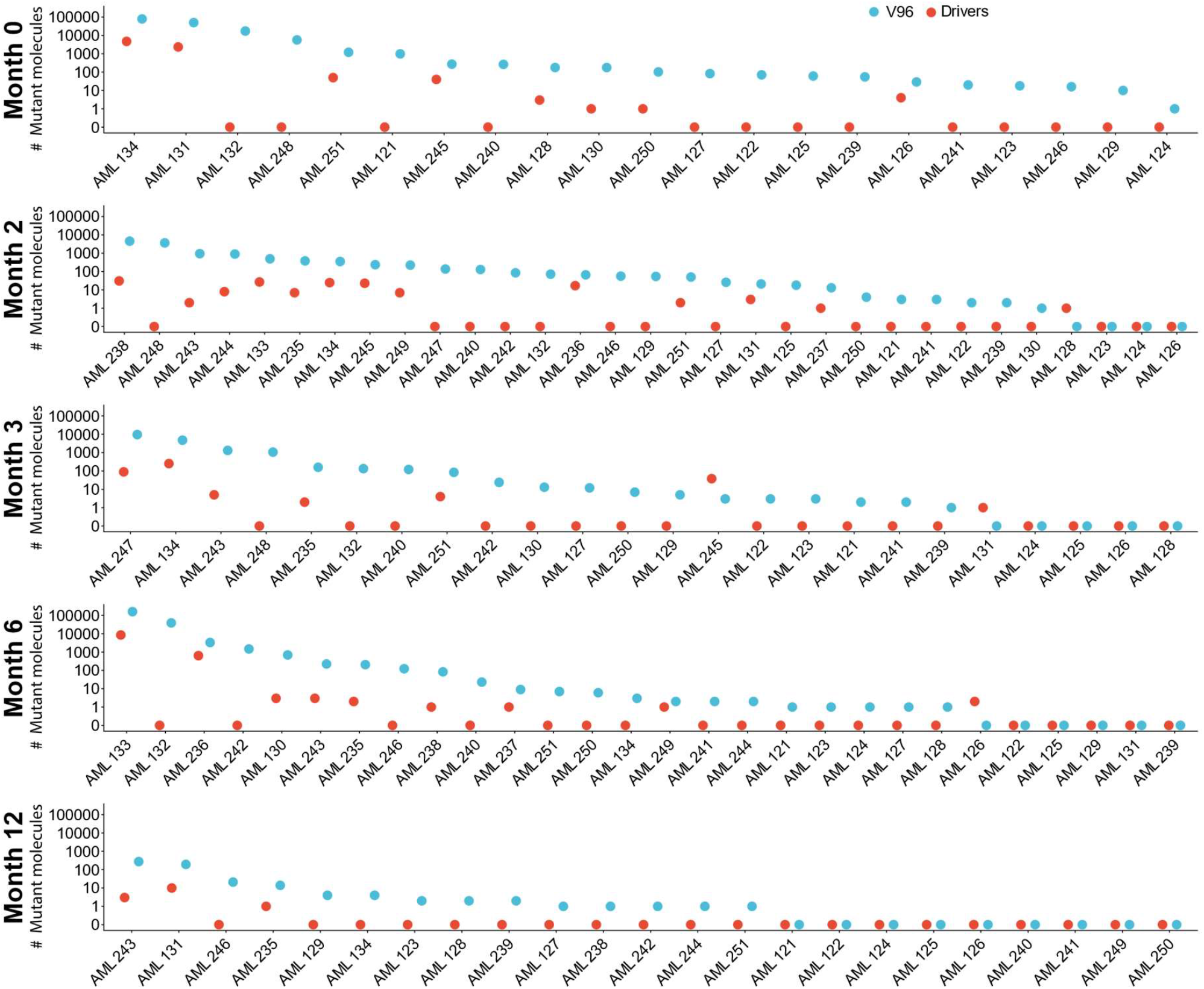
Number of mutant molecules in plasma detected through v96 or through analysis of driver gene mutations at each time point. Each panel represents a timepoint (month 0 (i.e., 7 days prior to transplantation), or 2, 3, 6, or 12 post-transplantation). Blue dots represent the number of mutant molecules detected when assessed with the mutations incorporated into v96. Red dots represent the number of mutant molecules detected when only driver mutations were assessed with the same duplex-sequencing assay used for v96 (see main text). In all cases, the DNA from 10-mL of cell-free plasma was assessed.

The relatively high number of mutations tracked by v96 allowed detection of clonal heterogeneity, represented by the presence of multiple clones. We used a simple method to infer the presence of multiple clones. We inferred that more than one clone was present if a subset of mutations had very similar mutant allele fractions and another subset of mutations had markedly different mutant allele fractions. Examples of the presence of multiple clones are shown in Fig. 6. Notably, the original driver gene mutation found at diagnosis was sometimes not present in the clone that drove relapse, such as in patient AML 132. In this patient, a *TP53* p.K120E mutation was found at diagnosis and during CR, but was eradicated thereafter, even though the patient relapsed through the expansion of a different clone.

**Fig. 6:**
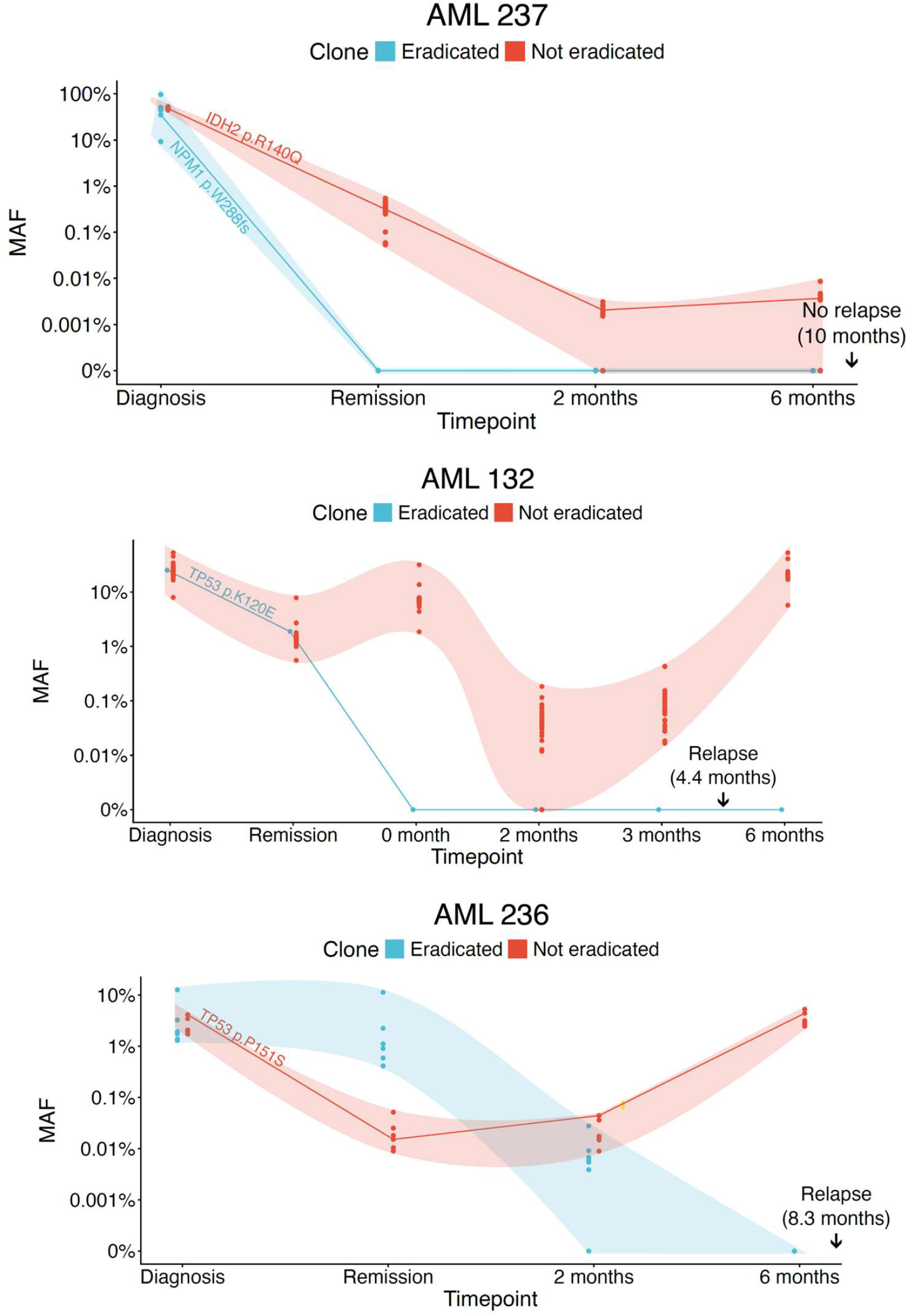
Clonal heterogeneity observed with v96. Three representative examples that demonstrate the detection of multiple clones and the change in clonal composition over time. A subset of mutations in v96 represent the clones that were apparently eradicated by transplantation (blue), while others represent clones that were persistent (red) over time. Driver mutations for each sample are depicted in addition to the mutations assessed by v96. The timepoints at diagnosis, remission, and months post-transplantation are shown on the X-axis. The shaded regions indicate 95 confidence intervals. 150 ng of bone marrow or peripheral blood cell DNA (SI Appendix, Table S2) was assayed for timepoints D and R, while cell-free DNA from 10-mL of plasma was assayed for the other time points.

## DISCUSSION

Five conclusions can be derived from the results of this study:

i. 100% of patients in complete clinical remission prior to transplantation had evidence of leukemia when a highly sensitive and specific assay for the disease was applied (*SI Appendix*, Table S2).
ii. Leukemic cells were still detectable in 90% of patients two months after transplantation.
iii. Further decreases in leukemic cell burden occurred after immunosuppression was discontinued, presumably due to GvL (*SI Appendix*, Table S2).
iv. Plasma provided a more informative and less invasive source of DNA for the analysis of residual leukemia cells than bone marrow (**Fig. 3**).
v. An assay that tracked multiple mutations, such as v96, was more informative than an assay for driver gene mutations (**Figs. 4 and 5**).

Modern techniques for detecting MRD include flow cytometry, as well as molecular genetic assays largely focused on driver gene mutations (11, 18-24). Of note, *all* the patients in the current study had easily detectable mutations at CR using v96, although only 6 of the 30 patients had flow cytometric evidence of MRD. Moreover, as detailed in *SI Appendix*, Table S2, only 19 of the 30 patients had a detectable driver mutation during CR with the highly sensitive SaferSeqS assay, the same assay used for the mutations assessed by v96 (17). The observation that the v96 assay is more informative than a state-of-the-art assay for driver gene mutations (Figs. 4 and 5) likely reflects the fact that the median number of distinct mutations assessed with v96 was 63, while the median number of driver gene mutations assessed was 2. This 32-fold difference in the number of mutations tracked is similar to the magnitude of the increased number of mutant molecules detected by the v96 assay compared to the driver gene mutation assay (21-fold, Fig. 4). It is important to point out that the passenger mutations tracked by v96, though not mechanistically responsible for the leukemia, are exquisitely specific to the leukemia cells by virtue of the way they were selected for analysis (Methods). They were all found at high clonal fractions in the leukemia cells present at diagnosis, indicating that they were present in a clone that developed into leukemia. Following transplantation, their identification indicates that a clone which previously developed into leukemia was still present in the patient. Whether that clone is considered a pre-leukemic clone, or a clonal hematopoiesis clone (25-27), is a matter of semantics. These mutations do not reflect clonal hematopoiesis of *indeterminate* potential (CHIP). The mutations present in the leukemia cells are far from *indeterminate*; they define the founder cells that evolved to become an actual malignancy. The presence of such clones presumably indicates a higher chance of subsequent leukemic development, though the magnitude of this probability is not yet known.

GvL is critical to the success of transplantation (3). However, it has heretofore been challenging to quantify and discriminate the effects of the conditioning regimen and immune-mediated effects (such as GvL) on leukemia cell reduction. In 90% (27 of 30) of the patients in our study, evidence for residual leukemia was found at 2 months following transplantation, including one patient (AML 129) who underwent a myeloablative conditioning regimen. The reductions in leukemia burden assessed by v96 generally occurred only after immunosuppression was discontinued, consistent with a GvL effect that was enhanced once the inhibitory effect of immunosuppression was eliminated.

Our observation that plasma DNA is considerably more informative than bone marrow DNA for the detection of residual leukemia (**Fig. 3**) is important to patients because a plasma-based assay offers a non-invasive option that allows more frequent monitoring. This is particularly critical for patients with bone marrow fibrosis for whom an aspiration is difficult to perform or for patients with extramedullary disease. Our new data are also consistent with previous studies that assessed driver gene mutations, and found plasma DNA to be more informative than bone marrow DNA (28-30). The ability to monitor plasma for residual disease in leukemia patients should allow a more granular view of the disease process through more frequent assessment of disease status than is currently practicable with bone marrow.

Although these data are encouraging, our study has limitations. In particular, more patients need to be assessed to validate the clinical implications of the v96 assay, particularly with respect to the probability of relapse when driver gene mutations are no longer detectable. If confirmed in larger cohorts, these data will have other important clinical implications for improving disease control for patients undergoing transplantation. The patients who relapsed had more than an order of magnitude higher (352-fold) number of mutant molecules during remission than the patients who did not relapse. Approaches to reduce tumor burden prior to transplantation, such as additional consolidation therapies or myeloablative conditioning, could be considered in these patients to optimize post-transplant outcome. There is emerging data that both immunologic and molecular targeted therapies given as maintenance after transplantation can cooperate with the GvL effect to avert relapses (31-33). A sensitive non-invasive test that quantifies leukemic burden through plasma analysis can be easily integrated into routine follow-up to inform clinical decisions regarding administration of these post-transplant maintenance therapies.

Finally, several questions related to the new data described in this paper can now be asked, and addressed, in future studies. What is the optimum time to discontinue immunosuppressive agents following transplantation? Any therapeutic effect, including those mediated by targeted therapies or T-cells, is more likely to be curative when administered to patients with less disease burden (34). Two months is now the standard time to discontinue immunosuppression in the absence of GvHD in many transplant centers, but this time may be too short or too long, depending on the patient and the degree of residual leukemia as well as the extent of GvHD. Similarly, when should a transplantation be considered a failure, and when should either a second transplantation or addition of another therapy such as targeted therapy or donor lymphocyte infusions be instituted? Longitudinal assessment of disease burdens through the evaluation of plasma with assays such as v96 could inform decisions about managing AML patients in the future.

## MATERIALS AND METHODS

### Patients

All AML patients who underwent transplantation at Johns Hopkins between November 2020 and January of 2022 were considered for participation in this study. Of them, we focused on those for whom the following samples were available: either bone marrow or peripheral blood at diagnosis, either bone marrow or peripheral blood at remission and prior to transplantation, and serial plasma samples collected before and following transplantation. Thirty patients from whom these samples were available for research purposes were included in the study (*SI Appendix*, Table S2). In total, plasma was available at 7 days before transplantation in 21 of 30 patients, 2 months after transplantation in all 30 patients, 3 months after transplantation in 23 patients, 6 months after transplantation in 27 patients, and one year after transplantation in 22 patients. The study was approved by the Institutional Review Board at The Johns Hopkins Medical Institutions. Written consent was provided by all patients.

### DNA purification and whole genome sequencing

DNA from bone marrow or peripheral blood leukocytes at diagnosis and remission were acquired from the Johns Hopkins Molecular Diagnostics Laboratory. Libraries were prepared from this DNA after shearing through sonication as described previously (17), with the following modifications. KAPA HiFi HotStart ReadyMix (Roche, Cat # 07958927001) was used to amplify the DNA following ligations using the following conditions: 98 °C for 45 s, followed by 8 cycles of 98 °C for 15 s, 60 °C for 30 s and 72 °C for 30 seconds. This produced ∼200 copies of each starting template molecule, which could be followed by the unique molecular barcode appended to each template molecule. The amplified DNA was purified using 1.8 X SPRI beads and eluted in 100 uL EB buffer (Qiagen, Cat# 19086). After seven additional PCR cycles used to add sequencing adapters to the libraries, whole genome sequencing was performed on an Illumina NovaSeq 6000 instrument or a Complete Genomics T7 instrument to a depth of ∼30-100 X. FASTQ files were generated using Illumina’s bcl2fastq or by Complete Genomic’s Ztron Lite Server, then aligned to hg38 reference genome with BWA-MEM with default settings (35). Duplicate sequencing clusters were removed with Picard (http://broadinstitute.github.io/picard). Variants in the leukemia cell-containing sample were called using Mutect2 (https://gatk.broadinstitute.org/hc/en-us/articles/360037593851-Mutect2) using remission bone marrow or peripheral blood samples as the matched normal (*SI Appendix*, Table S2).

Approximately 96 candidate, leukemia-specific mutations per patient were selected after excluding mutations in repetitive regions, regions with difficult alignments to the reference genome (hg38), regions that were difficult to amplify efficiently, regions containing single nucleotide polymorphisms, or transitions at CpG sites. These exclusions were informed through prior analysis of samples from individuals without cancer, using whole genome or targeted sequencing. For the mutations that pass these selection criteria, the ones with the highest mutant allele fractions were selected as these more likely represent truncal mutations represent in the majority of subclones.

### The v96 assay

The 96 candidate, leukemia-specific mutations described above were then used to design a personalized assay dubbed “v96” for each patient. For each of the 96 candidate mutations chosen, primers were designed as described previously (17). Primers for all 96 candidate mutations were combined into a single tube for each patient. A hemi-nested, two-stage PCR protocol was used to amplify the regions containing the candidate mutations as described previously (17), except that KAPA HiFi HotStart polymerase was used for amplification ReadyMix (Roche, Indianapolis, IN; cat # KR0370) as detailed above. Following sequencing on a NovaSeq 6000 or a Complete Genomics T7 instrument, the data were evaluated as described (17). To be considered a *bona fide* mutation, the mutation had to be present in both the Watson and Crick strands in the DNA from the leukemia cells, present at low mutant allele fraction (<20%) in DNA from the matched patient sample obtained at remission to exclude germline variants, and absent in the plasma sample of an unrelated healthy control to exclude artifactual mutation hotspots. The same v96 multiplex assay was subsequently applied to the plasma and post-transplant samples from each patient. Although approximately 96 mutations were assessed in every DNA sample from every patient, only the *bona fide* mutations among the candidate mutations were scored, which are listed in *SI Appendix*, Tables S2 and S4. The number of leukemia cells shedding ctDNA into the circulation was estimated as follows: Assuming 2500 mL of plasma in a typical individual, one mutant molecule found in 10 mL of plasma implies that there are 250 mutant molecules in the circulation at that time. The half-life of ctDNA is roughly one hour (16), so that 125 (= 250/2) mutant molecules are lost each hour. The same number of molecules are needed to maintain a steady state, i.e., number of cells being born each day = number of cells dying each day. Thus, 3000 new leukemia cells per day (125 molecules/hour x 24 hours) are required to generate one mutant molecule in 10 mL of plasma. This is a minimum estimate because not all cells that die release their DNA into the plasma (36).

### Plasma DNA isolation

Plasma was isolated from 20-mL of peripheral blood within 6 hours of venipuncture and stored at -80C. cfDNA was purified from these plasma samples using a Revolution cfDNA isolation kit (nRichDX, Irvine, CA; cat # PN 100131) according to the manufacturer’s protocol. Libraries were made from the purified cfDNA as described above for cellular DNA except that the DNA from plasma was not sheared. The personalized v96 assays were then applied to these cfDNA samples as described above. The number of mutant molecules depicted in the figures and tables of this paper refer to the number of mutant molecules found in the libraries prepared from the ∼10-mL of plasma prepared from 20-mL of peripheral blood. The total number of molecules assessed, as reported in the Tables, also refers to the number of molecules identified in the libraries prepared from 10-mL of plasma.

### Driver gene mutations

Driver gene mutations were identified by the Leukemia Panel in the Johns Hopkins Molecular Diagnostics Laboratory in a CLIA-certified test, which includes 90 hematologic malignancy-related genes (https://pathology.jhu.edu/jhml-services/test-directory/leukemia-panel-ngs-bm). Detection of FLT3-ITD in bone marrow and peripheral blood samples was performed as previously described (28). To evaluate driver gene mutations in plasma, we used the same DNA libraries and approach described above for the v96 assay, but only included primers designed to amplify the genomic regions containing the driver gene mutations. When logistically feasible, the primers used to assess driver genes were mixed into the multiplex PCRs used for v96. In all cases, 10% of the identical libraries (representing ∼20 copies of each template molecule) were used to assess the v96-based mutations and the driver gene mutations.

## Data Availability

All data produced in the present study are available upon reasonable request to the authors

https://ega-archive.org/studies/EGAS00001007969

## Data, Materials, and Software Availability

Anonymized genetic sequencing data have been deposited in EGA (Sequencing data is deposited in https://ega-archive.org/studies/EGAS00001007969)(37).

## ACKNOWLEDGEMENTS

The authors would like to thank Cherie Blair, Kathy Judge, Deborah Cummings-Thomas, and Mason Alston for technical assistance, as well as the patients in the study for their courage and generosity. This study was supported by National Institutes of Health grant R21NS113016 (CB), National Institutes of Health grant RA37CA230400 (CB), National Institutes of Health grant U01CA230691 (NP, CB), Oncology Core CA 06973 (BV, KWK, NP), The Virginia and D.K. Ludwig Fund for Cancer Research (BV, KWK, NP, CB, CD), Commonwealth Fund (NP), The Sol Goldman Sequencing Facility at Johns Hopkins (BV), The Benjamin Baker Endowment (CD, YW),, Swim Across America (CD, SP), The V Foundation for Cancer Research (YW), Break Through Cancer Foundation (LPG), National Institutes of Health grant R01HL156144 (LPG) and P01CA225618 (RJJ).

## AUTHORSHIP CONTRIBUTIONS

YW., BV., RJJ., NP., KWK., LPG. contributed to conceptualization, methodology, writing the original draft, reviewing and editing. JX., S.P. MP., JP., LD., NS., CS, MZG, SM, SNM contributed to methodology and investigation. RJJ, MJL, CDG, CB, CMB, LPG contributed to methodology and resources. YW, BV, MP, NP, SP, SDC, CD contributed to formal analysis.

## COMPETITING INTEREST STATEMENT

BV and KWK are founders of Exact Sciences. KWK, and NP are advisors to, and hold equity in, Exact Sciences. BV, KWK, and NP are founders of, and own equity in, Clasp Therapeutics and Haystack Oncology (a Quest Diagnostics company). KWK, BV, and NP are consultants to, and hold equity in, CAGE Pharma. BV is a consultant to and holds equity in Catalio Capital Management. CB is a consultant to Haystack Oncology, and Privo Technologies. CB is a co-founder of Belay Diagnostics and OrisDx. MP is a consultant to Haystack Oncology. YW is a consultant to Belay Diagnostics and Exact Sciences. LD is an employee of Haystack Oncology. CD is a consultant and holds equity in Exact Sciences. CD is a co-founder of Belay Diagnostics. SM owns equity in Abbvie. The companies named above, as well as other companies, have licensed previously described technologies related to the work described in this paper from Johns Hopkins University. BV, KWK, NP, and CB are inventors on some of these technologies. Licenses to these technologies are or will be associated with equity or royalty payments to the inventors as well as to Johns Hopkins University. Patent applications on the work described in this paper may be filed by Johns Hopkins University. The terms of all these arrangements are being managed by Johns Hopkins University in accordance with its conflict-of-interest policies.

