## Supplemental Tables 1 to 4, Supplemental Figures 1 and 2 for "A plasma-based DNA test for quantification of disease burden in acute myeloid leukemia patients undergoing bone marrow transplantation"

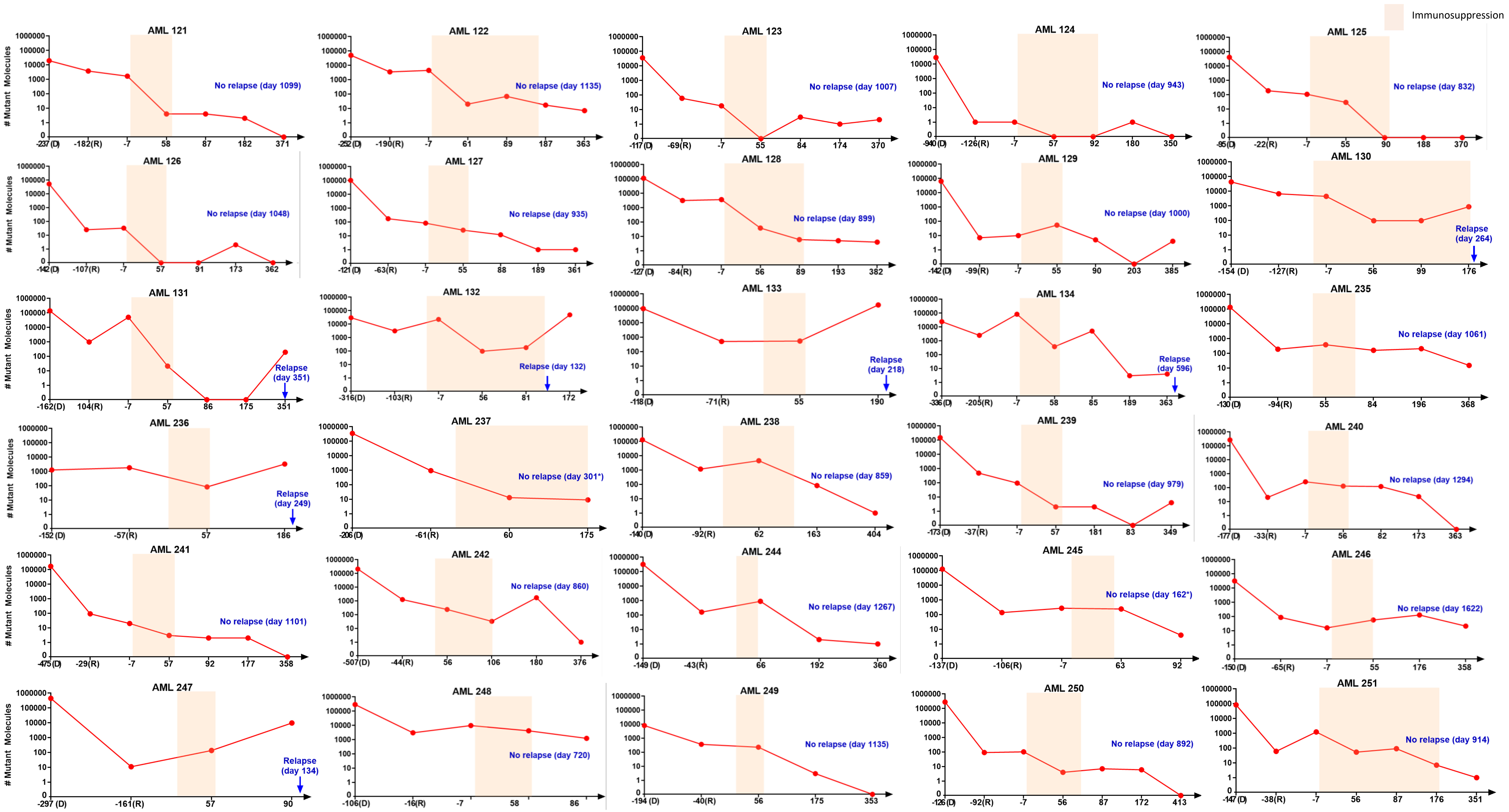


**Figure S1: Number of mutant molecules before and after transplantation.** Plots of the number of mutant molecules detected at diagnosis (D), complete remission (R), and at other time points for all 30 patients enrolled in this study. For each patient, 150 ng of bone marrow or peripheral blood cell DNA (see SI Table S1) was assayed by v96 for timepoints D and R, while cell-free DNA from 10-mL of plasma was assayed by v96 for the other time points. The x-axis represents the days relative to transplantation for these timepoints, with transplantation performed at day 0. For patients who relapsed, the post-transplantation day of relapse is indicated. For patients without relapse, the post-transplantation day of last follow-up is indicated (*Patients AML 237 and AML 245 passed away from unrelated causes without evidence of leukemic relapse). Colored regions represent intervals during which immunosuppression was administered.

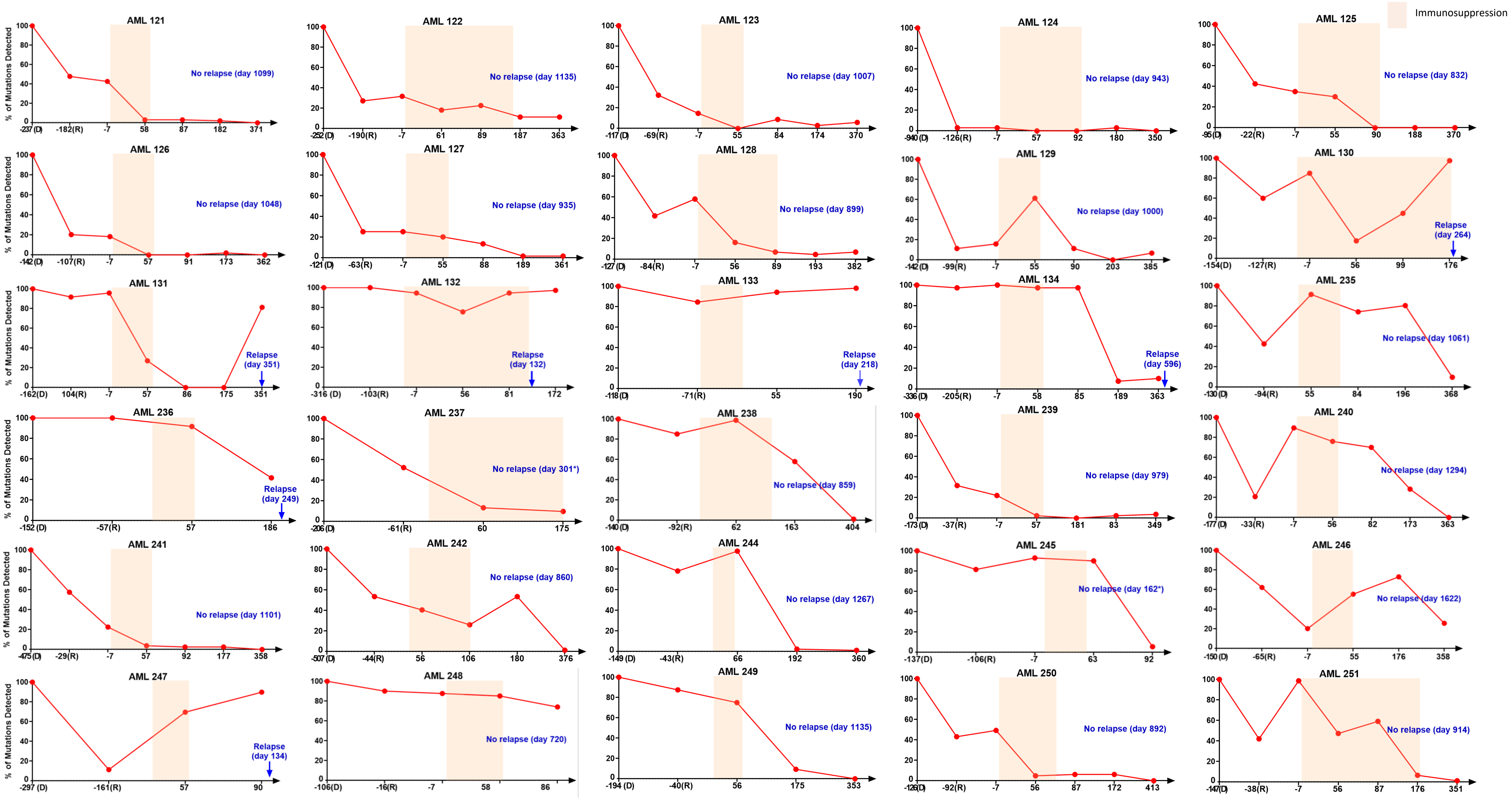


**Figure S2: Fraction of distinct mutations (rather than mutant molecules) detected before and after transplantation.** Plots of the fraction of distinct mutations detected at complete remission (R) and at other time points, relative to the total number of distinct mutations detected at diagnosis (D). This is different from the total number of mutant molecules depicted in Figure S1. The fractions in Figure S2 were normalized to the number of distinct mutations detected at diagnosis (always 100%). For example, if a patient had 80 distinct mutations detected at diagnosis, and 40 of those mutations were detected at remission, the fraction on the y-axis for the remission sample would be 50%. Bone marrow or peripheral blood cell DNA (see SI Table S2) was assayed by v96 for timepoints D and R, while cell-free DNA from plasma was assayed by v96 for the other time points. The x-axis represents the days relative to transplantation for these timepoints, with transplantation performed at day 0. For patients who relapsed, the post-transplantation day of relapse is indicated. For patients without relapse, the post-transplantation day of last follow-up is indicated (*Patients AML 237 and AML 245 passed away from unrelated causes without evidence of leukemic relapse). Colored regions represent intervals during which immunosuppression was administered.

**Table S1: Patient characteristics.**

| *Patient ID* | *Gender* | *Diagnosis* | *Cytogenetics* | *Induction therapy* | *Consolidation therapy* | *Fraction blasts by flow cytometry at remission* | *Match status (10/10, 9/10, 8/10, etc).* | *Conditioning* | *Type of transplant* | *Relapse Status* | *Immunsuppression end (Days post-alloHCT)* | *Time of relapse or last follow-up (Days post-alloHCT)* | *Decrease in mutation burden after Day 60* | *Decrease in mutation burden after immunosuppression discontinued* | *Post-transplant therapy* |
|---|---|---|---|---|---|---|---|---|---|---|---|---|---|---|---|
| AML 121 | Male | AML | Normal | HMA + venetoclax | HMA + venetoclax | negative | Unrelated (10/10) | 14.5 mg/kg day -6, -5 cyclophosphamide, 30mg/m2, day -6 to day -2 fludarabine, and TBI 400cGy | non-myeloablative fully matched unrelated donor peripheral blood stem cell transplant | No relapse | 60 | 1099 | Yes | Yes | None |
| AML 122 | Female | AML | Normal | 7+3+GO | HiDAC x2 | negative | Unrelated (8/10) | 14.5 mg/kg day -6, -5 cyclophosphamide, 30mg/m2, day -6 to day -2 fludarabine, and TBI 400cGy | non-myeloablative mismatched unrelated donor peripheral blood stem cell transplant | No relapse | 173 | 1135 | Yes | Yes | ivosidenib |
| AML 123 | Female | AML | Normal | 7+3+ midostaurin | HiDAC | negative | Haploidentical (5/10) | 14.5 mg/kg day -6, -5 cyclophosphamide, 30mg/m2, day -6 to day -2 fludarabine, and TBI 400cGy | non-myeloablative haploidentical bone marrow transplant | No relapse | 60 | 1007 | No mutation detected at Day 60 | No mutations detected prior to immunosupression cessation | gilteritinib |
| AML 124 | Female | rAML | inv(16)(p13.1q22) | 7+3+GO | HiDAC | negative | Unrelated (9/10) | 14.5 mg/kg day -6, -5 cyclophosphamide, 30mg/m2, day -6 to day -2 fludarabine, and TBI 200cGy | non-myeloablative mismatched unrelated donor peripheral blood stem cell transplant | No relapse | 92 | 943 | No mutation detected at Day 60 | No mutations detected prior to immunosupression cessation | None |
| AML 125 | Male | AML | Normal | 7+3 | none | negative | Haploidentical (5/10) | 14.5 mg/kg day -6, -5 cyclophosphamide, 30mg/m2, day -6 to day -2 fludarabine, and TBI 400cGy | non-myeloablative haploidentical bone marrow transplant | No relapse | 107 | 832 | Yes | No mutations detected prior to immunosupression cessation | enasidenib |
| AML 126 | Female | AML | Normal | 7+3 | HiDAC x2 | negative | Haploidentical (5/10) | 14.5 mg/kg day -6, -5 cyclophosphamide, 30mg/m2, day -6 to day -2 fludarabine, and TBI 400cGy | non-myeloablative haploidentical bone marrow transplant | No relapse | 60 | 1048 | No mutation detected at Day 60 | No mutations detected prior to immunosupression cessation | None |
| AML 127 | Male | AML | del(13)(q12q14) | Vyxeos+ midostaurin | Vyxeos + midostaurin | negative | Haploidentical (5/10) | 14.5 mg/kg day -6, -5 cyclophosphamide, 30mg/m2, day -6 to day -2 fludarabine, and TBI 400cGy | non-myeloablative haploidentical bone marrow transplant | No relapse | 60 | 935 | Yes | Yes | gilteritinib |
| AML 128 | Female | AML | Normal | 7+3+ midostaurin | HiDAC + gilteritinib | negative | Haploidentical (8/10) | 14.5 mg/kg day -6, -5 cyclophosphamide, 30mg/m2, day -6 to day -2 fludarabine, and TBI 400cGy | non-myeloablative haploidentical bone marrow transplant | No relapse | 100 | 899 | Yes | Yes | gilteritinib, ivosidenib |
| AML 129 | Male | AML | Normal | 7+3+ midostaurin | HiDAC + gilteritinib | negative | Unrelated (10/10) | 1 mg/kg/dose, Oral, Every 6 hours Busulphan day -8 and -7 and 1 mg/kg/dose, Oral, Every 5 hours Busulphan on day -6. 50mg/kg/dose cyclophospahmide on days -4 and -3 | myeloablative allogeneic transplant fully matched unrelated donor peripheral blood stem cell transplant | No relapse | 60 | 1000 | Yes | Yes | gilteritinib |
| AML 130 | Male | AML | Complex | HMA + venetoclax | HMA + venetoclax | negative | Unrelated (8/10) | 14.5 mg/kg day -6, -5 cyclophosphamide, 30mg/m2, day -6 to day -2 fludarabine, and TBI 400cGy | non-myeloablative mismatched unrelated donor peripheral blood stem cell transplant | Relapsed | 180 | 264 | No | Immunosupression stopped after last time point | None |
| AML 131 | Male | sAML | Complex | Vyxeos | Vyxeos | 0.8% | Haploidentical (5/10) | 14.5 mg/kg day -6, -5 cyclophosphamide, 30mg/m2, day -6 to day -2 fludarabine, and TBI 400cGy | non-myeloablative haploidentical peripheral blood stem cell transplant | Relapsed | 60 | 351 | Yes | Yes | Donor lymphocyte infusion |
| AML 132 | Female | AML-TP53 | Complex | Decitabine | none | equivocal | Haploidentical (5/10) | 14.5 mg/kg day -6, -5 cyclophosphamide, 30mg/m2, day -6 to day -2 fludarabine, and TBI 400cGy | non-myeloablative haploidentical peripheral blood stem cell transplant | Relapsed | 130 | 132 | No | No | None |
| AML 133 | Male | AML-MRC | Normal | 7+3+ midostaurin | HiDAC + midostaurin | negative | Haploidentical (5/10) | 14.5 mg/kg day -6, -5 cyclophosphamide, 30mg/m2, day -6 to day -2 fludarabine, and TBI 400cGy | non-myeloablative haploidentical peripheral blood stem cell transplant | Relapsed | 71 | 218 | No | No | None |
| AML 134 | Male | sAML | Complex | Decitabine+ ruxolitinib | none | 0.8% | Haploidentical (5/10) | 14.5 mg/kg day -6, -5 cyclophosphamide, 30mg/m2, day -6 to day -2 fludarabine, and TBI 400cGy | non-myeloablative haploidentical peripheral blood stem cell transplant | Relapsed | 60 | 596 | Yes | Yes | None |
| AML 235 | Male | AML | -Y,t(8;21)(q22;q22) | 7+3+GO | HiDAC | negative | Haploidentical( 5/10) | 14.5 mg/kg day -6, -5 cyclophosphamide, 30mg/m2,day -6 to day -2 fludarabine, and TBI 400cGy | non-myeloablative haploidentical bone marrow transplant | No relapse | 60 | 1061 | Yes | Yes | None |
| AML 236 | Male | MDS | Complex | Decitabine + venetoclax | Decitabine + venetoclax | 0.08% | Haploidentical( 5/10) | 14.5 mg/kg day -6, -5 cyclophosphamide, 30mg/m2,day -6 to day -2 fludarabine, and TBI 400cGy | non-myeloablative haploidentical peripheral blood stem cell transplant | Relapsed | 60 | 249 | No | No | aza,Decitabine |
| AML 237 | Female | AML | Normal | Vyxeo | HiDAC | negative | Unrelated (7/10) | 14.5 mg/kg day -6, -5 cyclophosphamide, 30mg/m2,day -6 to day -2 fludarabine, and TBI 400cGy | non-myeloablative mismatched unrelated donor peripheral blood stem cell transplant | No relapse | 180 | 301 | Yes | Immunosupression stopped after last time point | None |
| AML 238 | Female | AML | Normal | Vyxeos | HiDAC | negative | Unrelated (10/10) | 14.5 mg/kg day -6, -5 cyclophosphamide, 30mg/m2,day -6 to day -2 fludarabine, and TBI 400cGy | non-myeloablative fully matched unrelated donor peripheral blood stem cell transplant | No relapse | 138 | 859 | Yes | Yes | None |
| AML 239 | Female | AML | inv(9)(p12q13) | 7+3+ midostaurin | HiDAC + gilteritinib | negative | Unrelated (9/10) | 14.5 mg/kg day -6, -5 cyclophosphamide, 30mg/m2,day -6 to day -2 fludarabine, and TBI 200cGy | non-myeloablative mismatched unrelated donor peripheral blood stem cell transplant | No relapse | 90 | 979 | Yes | Yes | gilteritinib |
| AML 240 | Male | AML | Normal | Decitabine + venetoclax | Venetoclax | negative | Haploidentical( 5/10) | 14.5 mg/kg day -6, -5 cyclophosphamide, 30mg/m2,day -6 to day -2 fludarabine, and TBI 400cGy | non-myeloablative haploidentical peripheral blood stem cell transplant | No relapse | 57 | 1294 | Yes | Yes | None |

| | | | | | | | | | | | | | | | |
|---|---|---|---|---|---|---|---|---|---|---|---|---|---|---|---|
| AML 241 | Male | AML | insufficient cell growth | 7+3 | Decitabine + venetoclax | 0.5% | Haploidentical( 5/10) | 14.5 mg/kg day -6, -5 cyclophosphamide, 30mg/m2,day -6 to day -2 fludarabine, and TBI 400cGy | non-myeloablative haploidentical peripheral blood stem cell transplant | No relapse | 60 | 1101 | Yes | Yes | None |
| AML 242 | Male | AML | +8 | Azacitidine + venetoclax | Azacitidine + venetoclax | negative | Unrelated (8/10) | 14.5 mg/kg day -6, -5 cyclophosphamide, 30mg/m2,day -6 to day -2 fludarabine, and TBI 400cGy | non-myeloablative mismatched unrelated donor peripheral blood stem cell transplant | No relapse | 106 | 860 | Yes | Yes | None |
| AML 244 | Female | AML | Normal | 7+3+ midostaurin | HiDAC + midostaurin | negative | Haploidentical( 5/10) | 14.5 mg/kg day -6, -5 cyclophosphamide, 30mg/m2,day -6 to day -2 fludarabine, and TBI 400cGy | non-myeloablative haploidentical bone marrow transplant | No relapse | 60 | 1267 | Yes | Yes | None |
| AML 245 | Male | AML | Normal | 7+3+ midostaurin | HiDAC + midostaurin | negative | Haploidentical( 5/10) | 14.5 mg/kg day -6, -5 cyclophosphamide, 30mg/m2,day -6 to day -2 fludarabine, and TBI 400cGy | non-myeloablative haploidentical bone marrow transplant | No relapse | 57 | 162 | Yes | Yes | None |
| AML 246 | Male | MDS | Complex | Decitabine | Decitabine | negative | Haploidentical( 5/10) | 14.5 mg/kg day -6, -5 cyclophosphamide, 30mg/m2,day -6 to day -2 fludarabine, and TBI 400cGy | non-myeloablative haploidentical peripheral blood stem cell transplant | No relapse | 60 | 1622 | Yes | Yes | None |
| AML 247 | Female | AML | t(6:11)(q27;q23) | 7+3+GO | HiDAC | negative | Haploidentical (7/10) | 14.5 mg/kg day -6, -5 cyclophosphamide, 30mg/m2,day -6 to day -2 fludarabine, and TBI 400cGy | non-myeloablative haploidentical peripheral blood stem cell transplant | Relapsed | 60 | 134 | No | No | None |
| AML 248 | Male | AML | Normal | Vyxeos | HiDAC | negative | Haploidentical( 5/10) | 14.5 mg/kg day -6, -5 cyclophosphamide, 30mg/m2,day -6 to day -2 fludarabine, and TBI 400cGy | non-myeloablative haploidentical bone marrow transplant | No relapse | 60 | 720 | Yes | Yes | enasidenib |
| AML 249 | Male | AML | Complex | Vyxeos | Vyxeos | 0.8% | Haploidentical (5/10) | 14.5 mg/kg day -6, -5 cyclophosphamide, 30mg/m2,day -6 to day -2 fludarabine, and TBI 400cGy | non-myeloablative haploidentical peripheral blood stem cell transplant | No relapse | 66 | 1135 | Yes | Yes | None |
| AML 250 | Female | AML | Normal | 7+3+ midostaurin | HiDAC | negative | Haploidentical( 5/10) | 14.5 mg/kg day -6, -5 cyclophosphamide, 30mg/m2,day -6 to day -2 fludarabine, and TBI 400cGy | non-myeloablative haploidentical bone marrow transplant | No relapse | 70 | 892 | Yes | Yes | gilteritinib |
| AML 251 | Female | AML | Complex | 7+3 | Azacitidine + venetoclax | <0.1% | Unrelated (9/10) | 14.5 mg/kg day -6, -5 cyclophosphamide, 30mg/m2,day -6 to day -2 fludarabine, and TBI 400cGy | non-myeloablative mismatched unrelated donor peripheral blood stem cell transplant | No relapse | 180 | 914 | Yes | Yes | None |

*sAML = secondary AML, rAML = relapsed AML, AML-TP53 = TP53-mutated AML; AML-MRC = Acute Myeloid Leukemia with myelodysplasia-related changes; HMA = hypomethylating agent; GO = gemtuzumab ozogamicin, HiDAC = High-Dose Cytarabine*

**Table S2: Numbers of mutants detected and mutant allele frequencies in the mutations assessed through v96 as well as the driver gene mutations assessessed in the same DNA samples.** For each patient, 150 ng of bone marrow or peripheral blood cell DNA was assayed, and all the DNA purified from 10-mL of plasma was assayed.

| Patient | Sample Type | Time point | Day post-alloHCT | *Day on which immunsuppression ended (Days post-alloHCT)* | *Total umber of v96 mutations assessed* | *Number of v96 mutations found* | *Fraction of v96 mutations found* | *Total number of mutant molecules found in v96* | *Average number of molecules assessed in v96 (mutant + non-mutant)* | *Average Mutant Allele Frequency in v96* | *Total number of driver gene mutations assessed* | *Number of positive driver gene mutations in sample* | *Total number of mutant molecules in driver genes* | *Average number of molecules assessed in driver genes (mutant + non-mutant)* | *Average mutant allele fraction in driver genes* |
|---|---|---|---|---|---|---|---|---|---|---|---|---|---|---|---|
| AML 121 | Bone Marrow | Diagnostic | Minus 237 | 60 | 96 | 96 | 100.0% | 19672 | 427618 | 4.60% | 1 | 1 | 31 | 583 | 5.32% |
| AML 121 | Bone Marrow | Remission | Minus 182 | 60 | 96 | 46 | 47.9% | 3752 | 570555 | 0.66% | 1 | 0 | 0 | 838 | 0.00% |
| AML 121 | Plasma | 0 month | Minus 7 | 60 | 96 | 41 | 42.7% | 1664 | 399406 | 0.42% | 1 | 0 | 0 | 329 | 0.00% |
| AML 121 | Plasma | 2 months | 58 | 60 | 96 | 3 | 3.1% | 4 | 826985 | 0.00% | 1 | 0 | 0 | 597 | 0.00% |
| AML 121 | Plasma | 3 months | 87 | 60 | 96 | 3 | 3.1% | 4 | 889803 | 0.00% | 1 | 0 | 0 | 432 | 0.00% |
| AML 121 | Plasma | 6 months | 182 | 60 | 96 | 2 | 2.1% | 2 | 455120 | 0.00% | 1 | 0 | 0 | 583 | 0.00% |
| AML 121 | Plasma | 12 months | 371 | 60 | 96 | 0 | 0.0% | 0 | 459369 | 0.00% | 1 | 0 | 0 | 748 | 0.00% |
| AML 122 | Peripheral blood | Diagnostic | Minus 252 | 173 | 44 | 44 | 100.0% | 50290 | 109924 | 45.75% | 3 | 3 | 3025 | 10709 | 28.25% |
| AML 122 | Peripheral blood | Remission | Minus 190 | 173 | 44 | 12 | 27.3% | 3546 | 235788 | 1.50% | 3 | 0 | 0 | 22960 | 0.00% |
| AML 122 | Plasma | 0 month | Minus 7 | 173 | 44 | 14 | 31.8% | 4503 | 237439 | 1.90% | 3 | 0 | 0 | 24150 | 0.00% |
| AML 122 | Plasma | 2 months | 61 | 173 | 44 | 8 | 18.2% | 20 | 210861 | 0.01% | 3 | 0 | 0 | 24201 | 0.00% |
| AML 122 | Plasma | 3 months | 89 | 173 | 44 | 10 | 22.7% | 69 | 491933 | 0.01% | 3 | 0 | 0 | 47374 | 0.00% |
| AML 122 | Plasma | 6 months | 187 | 173 | 44 | 5 | 11.4% | 17 | 227614 | 0.01% | 3 | 0 | 0 | 23047 | 0.00% |
| AML 122 | Plasma | 12 months | 363 | 173 | 44 | 5 | 11.4% | 7 | 197021 | 0.00% | 3 | 0 | 0 | 17861 | 0.00% |
| AML 123 | Peripheral blood | Diagnostic | Minus 117 | 60 | 34 | 34 | 100.0% | 35926 | 82759 | 43.41% | 3 | 3 | 2280 | 5561 | 41.00% |
| AML 123 | Peripheral blood | Remission | Minus 69 | 60 | 34 | 11 | 32.4% | 59 | 137030 | 0.04% | 3 | 1 | 3 | 10446 | 0.03% |
| AML 123 | Plasma | 0 month | Minus 7 | 60 | 34 | 5 | 14.7% | 18 | 46908 | 0.04% | 3 | 0 | 0 | 2803 | 0.00% |
| AML 123 | Plasma | 2 months | 55 | 60 | 34 | 0 | 0.0% | 0 | 59172 | 0.00% | 3 | 0 | 0 | 4650 | 0.00% |
| AML 123 | Plasma | 3 months | 84 | 60 | 34 | 3 | 8.8% | 3 | 142714 | 0.00% | 3 | 0 | 0 | 8631 | 0.00% |
| AML 123 | Plasma | 6 months | 174 | 60 | 34 | 1 | 2.9% | 1 | 138898 | 0.00% | 3 | 0 | 0 | 8410 | 0.00% |
| AML 123 | Plasma | 12 months | 370 | 60 | 34 | 2 | 5.9% | 2 | 59217 | 0.00% | 3 | 0 | 0 | 3558 | 0.00% |
| AML 124 | Bone Marrow | Diagnostic | Minus 940 | 92 | 33 | 33 | 100.0% | 28684 | 72941 | 39.32% | 3 | 3 | 721 | 10805 | 6.67% |
| AML 124 | Bone Marrow | Remission | Minus 126 | 92 | 33 | 1 | 3.0% | 1 | 134353 | 0.00% | 3 | 0 | 0 | 18659 | 0.00% |
| AML 124 | Plasma | 0 month | Minus 7 | 92 | 33 | 1 | 3.0% | 1 | 56937 | 0.00% | 3 | 0 | 0 | 6499 | 0.00% |
| AML 124 | Plasma | 2 months | 57 | 92 | 33 | 0 | 0.0% | 0 | 84440 | 0.00% | 3 | 0 | 0 | 11624 | 0.00% |
| AML 124 | Plasma | 3 months | 92 | 92 | 33 | 0 | 0.0% | 0 | 134460 | 0.00% | 3 | 0 | 0 | 18492 | 0.00% |
| AML 124 | Plasma | 6 months | 180 | 92 | 33 | 1 | 3.0% | 1 | 64284 | 0.00% | 3 | 0 | 0 | 7783 | 0.00% |
| AML 124 | Plasma | 12 months | 350 | 92 | 33 | 0 | 0.0% | 0 | 84200 | 0.00% | 3 | 0 | 0 | 9742 | 0.00% |
| AML 125 | Peripheral blood | Diagnostic | Minus 95 | 107 | 40 | 40 | 100.0% | 41019 | 109490 | 37.46% | 1 | 1 | 347 | 1641 | 21.15% |
| AML 125 | Peripheral blood | Remission | Minus 22 | 107 | 40 | 17 | 42.5% | 188 | 208326 | 0.09% | 1 | 0 | 0 | 3096 | 0.00% |
| AML 125 | Plasma | 0 month | Minus 7 | 107 | 40 | 14 | 35.0% | 107 | 124905 | 0.09% | 1 | 0 | 0 | 758 | 0.00% |
| AML 125 | Plasma | 2 months | 55 | 107 | 40 | 12 | 30.0% | 29 | 441348 | 0.01% | 1 | 0 | 0 | 1204 | 0.00% |
| AML 125 | Plasma | 3 months | 90 | 107 | 40 | 0 | 0.0% | 0 | 247780 | 0.00% | 1 | 0 | 0 | 689 | 0.00% |
| AML 125 | Plasma | 6 months | 188 | 107 | 40 | 0 | 0.0% | 0 | 215457 | 0.00% | 1 | 0 | 0 | 562 | 0.00% |
| AML 125 | Plasma | 12 months | 370 | 107 | 40 | 0 | 0.0% | 0 | 75175 | 0.00% | 1 | 0 | 0 | 745 | 0.00% |
| AML 126 | Bone Marrow | Diagnostic | Minus 142 | 60 | 49 | 49 | 100.0% | 53414 | 141502 | 37.75% | 1 | 1 | 1978 | 4844 | 40.83% |
| AML 126 | Bone Marrow | Remission | Minus 107 | 60 | 49 | 10 | 20.4% | 25 | 283381 | 0.01% | 1 | 1 | 3 | 8577 | 0.03% |
| AML 126 | Plasma | 0 month | Minus 7 | 60 | 49 | 9 | 18.4% | 33 | 284688 | 0.01% | 1 | 1 | 4 | 10797 | 0.04% |
| AML 126 | Plasma | 2 months | 57 | 60 | 49 | 0 | 0.0% | 0 | 202496 | 0.00% | 1 | 0 | 0 | 7760 | 0.00% |
| AML 126 | Plasma | 3 months | 91 | 60 | 49 | 0 | 0.0% | 0 | 236950 | 0.00% | 1 | 0 | 0 | 9540 | 0.00% |
| AML 126 | Plasma | 6 months | 173 | 60 | 49 | 1 | 2.0% | 2 | 430344 | 0.00% | 1 | 1 | 2 | 17939 | 0.01% |
| AML 126 | Plasma | 12 months | 362 | 60 | 49 | 0 | 0.0% | 0 | 263317 | 0.00% | 1 | 0 | 0 | 9818 | 0.00% |
| AML 127 | Peripheral blood | Diagnostic | Minus 121 | 60 | 59 | 59 | 100.0% | 102151 | 227495 | 44.90% | 2 | 2 | 448 | 4981 | 8.99% |
| AML 127 | Peripheral blood | Remission | Minus 63 | 60 | 59 | 15 | 25.4% | 173 | 380831 | 0.05% | 2 | 0 | 0 | 6520 | 0.00% |
| AML 127 | Plasma | 0 month | Minus 7 | 60 | 59 | 15 | 25.4% | 83 | 161768 | 0.05% | 2 | 0 | 0 | 2648 | 0.00% |
| AML 127 | Plasma | 2 months | 55 | 60 | 59 | 12 | 20.3% | 26 | 697824 | 0.00% | 2 | 0 | 0 | 8906 | 0.00% |
| AML 127 | Plasma | 3 months | 88 | 60 | 59 | 8 | 13.6% | 12 | 713661 | 0.00% | 2 | 0 | 0 | 8525 | 0.00% |
| AML 127 | Plasma | 6 months | 189 | 60 | 59 | 1 | 1.7% | 1 | 785223 | 0.00% | 2 | 0 | 0 | 11039 | 0.00% |
| AML 127 | Plasma | 12 months | 361 | 60 | 59 | 1 | 1.7% | 1 | 726358 | 0.00% | 2 | 0 | 0 | 9438 | 0.00% |
| AML 128 | Peripheral blood | Diagnostic | Minus 127 | 100 | 43 | 43 | 100.0% | 112872 | 235202 | 47.99% | 3 | 3 | 6684 | 15854 | 42.16% |
| AML 128 | Peripheral blood | Remission | Minus 84 | 100 | 43 | 18 | 41.9% | 3168 | 519903 | 0.61% | 3 | 2 | 450 | 38367 | 1.17% |
| AML 128 | Plasma | 0 month | Minus 7 | 100 | 43 | 25 | 58.1% | 3691 | 572531 | 0.64% | 3 | 2 | 398 | 30114 | 1.32% |
| AML 128 | Plasma | 2 months | 56 | 100 | 43 | 7 | 16.3% | 38 | 493083 | 0.01% | 3 | 2 | 9 | 27898 | 0.03% |
| AML 128 | Plasma | 3 months | 89 | 100 | 43 | 3 | 7.0% | 6 | 283016 | 0.00% | 3 | 1 | 2 | 17322 | 0.01% |
| AML 128 | Plasma | 6 months | 193 | 100 | 43 | 2 | 4.7% | 5 | 257615 | 0.00% | 3 | 1 | 1 | 16064 | 0.01% |
| AML 128 | Plasma | 12 months | 382 | 100 | 43 | 3 | 7.0% | 4 | 257526 | 0.00% | 3 | 1 | 1 | 15564 | 0.01% |
| AML 129 | Bone Marrow | Diagnostic | Minus 142 | 60 | 44 | 44 | 100.0% | 63803 | 155167 | 41.12% | 2 | 2 | 1670 | 5481 | 30.47% |
| AML 129 | Bone Marrow | Remission | Minus 99 | 60 | 44 | 5 | 11.4% | 7 | 305357 | 0.00% | 2 | 0 | 0 | 10651 | 0.00% |

| | | | | | | | | | | | | | | | |
|---|---|---|---|---|---|---|---|---|---|---|---|---|---|---|---|
| AML 129 | Plasma | 0 month | Minus 7 | 60 | 44 | 7 | 15.9% | 10 | 262417 | 0.00% | 2 | 0 | 0 | 8635 | 0.00% |
| AML 129 | Plasma | 2 months | 55 | 60 | 44 | 27 | 61.4% | 54 | 550703 | 0.01% | 2 | 0 | 0 | 20185 | 0.00% |
| AML 129 | Plasma | 3 months | 90 | 60 | 44 | 5 | 11.4% | 5 | 567902 | 0.00% | 2 | 0 | 0 | 20299 | 0.00% |
| AML 129 | Plasma | 6 months | 203 | 60 | 44 | 0 | 0.0% | 0 | 303532 | 0.00% | 2 | 0 | 0 | 10302 | 0.00% |
| AML 129 | Plasma | 12 months | 385 | 60 | 44 | 3 | 6.8% | 4 | 423185 | 0.00% | 2 | 0 | 0 | 16063 | 0.00% |
| AML 130 | Bone Marrow | Diagnostic | Minus 154 | 180 | 40 | 40 | 100.0% | 43354 | 130268 | 33.28% | 1 | 1 | 1101 | 3788 | 29.07% |
| AML 130 | Bone Marrow | Remission | Minus 127 | 180 | 40 | 24 | 60.0% | 6756 | 370843 | 1.82% | 1 | 0 | 0 | 14528 | 0.00% |
| AML 130 | Plasma | 0 month | Minus 7 | 180 | 40 | 34 | 85.0% | 4462 | 246264 | 1.81% | 1 | 1 | 1 | 7039 | 0.01% |
| AML 130 | Plasma | 2 months | 56 | 180 | 40 | 7 | 17.5% | 96 | 629914 | 0.02% | 1 | 0 | 0 | 17638 | 0.00% |
| AML 130 | Plasma | 3 months | 99 | 180 | 40 | 18 | 45.0% | 98 | 549073 | 0.02% | 1 | 0 | 0 | 15528 | 0.00% |
| AML 130 | Plasma | 6 months | 176 | 180 | 40 | 39 | 97.5% | 874 | 776320 | 0.11% | 1 | 1 | 9 | 19040 | 0.05% |
| AML 131 | Peripheral blood | Diagnostic | Minus 162 | 60 | 48 | 48 | 100.0% | 140365 | 293596 | 47.81% | 1 | 1 | 7238 | 7541 | 95.98% |
| AML 131 | Peripheral blood | Remission | Minus 104 | 60 | 48 | 44 | 91.7% | 990 | 213964 | 0.46% | 1 | 1 | 32 | 4604 | 0.70% |
| AML 131 | Plasma | 0 month | Minus 7 | 60 | 48 | 46 | 95.8% | 50086 | 269543 | 18.58% | 1 | 1 | 2346 | 6354 | 36.92% |
| AML 131 | Plasma | 2 months | 57 | 60 | 48 | 13 | 27.1% | 21 | 531339 | 0.00% | 1 | 1 | 3 | 14469 | 0.02% |
| AML 131 | Plasma | 3 months | 86 | 60 | 48 | 0 | 0.0% | 0 | 201164 | 0.00% | 1 | 1 | 1 | 4997 | 0.02% |
| AML 131 | Plasma | 6 months | 175 | 60 | 48 | 0 | 0.0% | 0 | 164554 | 0.00% | 1 | 0 | 0 | 3901 | 0.00% |
| AML 131 | Plasma | 12 months | 351 | 60 | 48 | 39 | 81.3% | 194 | 166008 | 0.12% | 1 | 1 | 10 | 3874 | 0.26% |
| AML 132 | Bone Marrow | Diagnostic | Minus 316 | 130 | 37 | 37 | 100.0% | 30016 | 113008 | 26.56% | 1 | 1 | 1261 | 5011 | 25.16% |
| AML 132 | Bone Marrow | Remission | Minus 103 | 130 | 37 | 37 | 100.0% | 3119 | 190491 | 1.64% | 1 | 1 | 138 | 8019 | 1.72% |
| AML 132 | Plasma | 0 month | Minus 7 | 130 | 37 | 35 | 94.6% | 22523 | 295005 | 7.63% | 1 | 0 | 0 | 9845 | 0.00% |
| AML 132 | Plasma | 2 months | 56 | 130 | 37 | 28 | 75.7% | 96 | 257548 | 0.04% | 1 | 0 | 0 | 9732 | 0.00% |
| AML 132 | Plasma | 3 months | 81 | 130 | 37 | 35 | 94.6% | 179 | 197259 | 0.09% | 1 | 0 | 0 | 7144 | 0.00% |
| AML 132 | Plasma | 6 months | 172 | 130 | 37 | 36 | 97.3% | 49051 | 245092 | 20.01% | 1 | 0 | 0 | 8343 | 0.00% |
| AML 133 | Peripheral blood | Diagnostic | Minus 118 | 71 | 52 | 52 | 100.0% | 96023 | 210867 | 45.54% | 4 | 4 | 11414 | 21791 | 52.38% |
| AML 133 | Peripheral blood | Remission | Minus 71 | 71 | 52 | 44 | 84.6% | 512 | 461181 | 0.11% | 4 | 4 | 46 | 52154 | 0.09% |
| AML 133 | Plasma | 2 months | 55 | 71 | 52 | 49 | 94.2% | 547 | 851764 | 0.06% | 4 | 4 | 55 | 73162 | 0.08% |
| AML 133 | Plasma | 6 months | 190 | 71 | 52 | 51 | 98.1% | 173381 | 748863 | 23.15% | 4 | 4 | 16781 | 61899 | 27.11% |
| AML 134 | Bone Marrow | Diagnostic | Minus 336 | 60 | 39 | 39 | 100.0% | 24540 | 69658 | 35.23% | 1 | 1 | 1516 | 2122 | 71.44% |
| AML 134 | Bone Marrow | Remission | Minus 205 | 60 | 39 | 38 | 97.4% | 2474 | 228936 | 1.08% | 1 | 1 | 122 | 6396 | 1.91% |
| AML 134 | Plasma | 0 month | Minus 7 | 60 | 39 | 39 | 100.0% | 84066 | 230508 | 36.47% | 1 | 1 | 4739 | 6747 | 70.24% |
| AML 134 | Plasma | 2 months | 58 | 60 | 39 | 38 | 97.4% | 378 | 296811 | 0.13% | 1 | 1 | 25 | 9412 | 0.27% |
| AML 134 | Plasma | 3 months | 85 | 60 | 39 | 38 | 97.4% | 5041 | 297184 | 1.70% | 1 | 1 | 251 | 8823 | 2.84% |
| AML 134 | Plasma | 6 months | 189 | 60 | 39 | 3 | 7.7% | 3 | 194220 | 0.00% | 1 | 0 | 0 | 5636 | 0.00% |
| AML 134 | Plasma | 12 months | 363 | 60 | 39 | 4 | 10.3% | 4 | 555521 | 0.00% | 1 | 1 | 1 | 18022 | 0.01% |
| AML 235 | Bone marrow | Diagnostic | Minus 130 | 60 | 82 | 82 | 100.0% | 135559 | 359181 | 37.74% | 3 | 3 | 1397 | 9146 | 15.27% |
| AML 235 | Bone marrow | Remission | Minus 94 | 60 | 82 | 35 | 42.7% | 192 | 594207 | 0.03% | 3 | 0 | 0 | 15894 | 0.00% |
| AML 235 | Plasma | 2 months | 55 | 60 | 82 | 75 | 91.5% | 384 | 3573548 | 0.01% | 3 | 3 | 7 | 124829 | 0.01% |
| AML 235 | Plasma | 3 months | 84 | 60 | 82 | 61 | 74.4% | 161 | 1562501 | 0.01% | 3 | 1 | 2 | 54452 | 0.00% |
| AML 235 | Plasma | 6 months | 196 | 60 | 82 | 66 | 80.5% | 208 | 837424 | 0.02% | 3 | 1 | 2 | 28231 | 0.01% |
| AML 235 | Plasma | 12 months | 368 | 60 | 82 | 8 | 9.8% | 15 | 319604 | 0.00% | 3 | 1 | 1 | 11342 | 0.01% |
| AML 236 | Bone marrow | Diagnostic | Minus 152 | 60 | 12 | 12 | 100.0% | 1270 | 48807 | 2.60% | 1 | 1 | 136 | 3269 | 4.16% |
| AML 236 | Bone marrow | Remission | Minus 57 | 60 | 12 | 12 | 100.0% | 1833 | 135814 | 1.35% | 1 | 1 | 1 | 6581 | 0.02% |
| AML 236 | Plasma | 2 months | 57 | 60 | 12 | 11 | 91.7% | 83 | 669929 | 0.01% | 1 | 1 | 17 | 38417 | 0.04% |
| AML 236 | Plasma | 6 months | 186 | 60 | 12 | 5 | 41.7% | 3299 | 244871 | 1.35% | 1 | 1 | 637 | 14416 | 4.42% |
| AML 237 | Bone marrow | Diagnostic | Minus 206 | 180 | 82 | 82 | 100.0% | 353072 | 779580 | 45.29% | 2 | 2 | 4737 | 9884 | 47.93% |
| AML 237 | Bone marrow | Remission | Minus 61 | 180 | 82 | 43 | 52.4% | 944 | 591339 | 0.16% | 2 | 1 | 23 | 7531 | 0.31% |
| AML 237 | Plasma | 2 months | 60 | 180 | 82 | 11 | 13.4% | 13 | 4948179 | 0.00% | 2 | 1 | 1 | 50514 | 0.00% |
| AML 237 | Plasma | 6 months | 175 | 180 | 82 | 8 | 9.8% | 9 | 2470781 | 0.00% | 2 | 1 | 1 | 29587 | 0.00% |
| AML 238 | Bone marrow | Diagnostic | Minus 140 | 138 | 81 | 81 | 100.0% | 130741 | 365619 | 35.76% | 1 | 1 | 1418 | 3978 | 35.65% |
| AML 238 | Bone marrow | Remission | Minus 92 | 138 | 81 | 69 | 85.2% | 1193 | 561561 | 0.21% | 1 | 1 | 6 | 6043 | 0.10% |
| AML 238 | Plasma | 2 months | 62 | 138 | 81 | 80 | 98.8% | 4532 | 3998171 | 0.11% | 1 | 1 | 31 | 38724 | 0.08% |
| AML 238 | Plasma | 6 months | 163 | 138 | 81 | 47 | 58.0% | 83 | 668128 | 0.01% | 1 | 1 | 1 | 6664 | 0.02% |
| AML 238 | Plasma | 12 months | 404 | 138 | 81 | 1 | 1.2% | 1 | 620635 | 0.00% | 1 | 0 | 0 | 6096 | 0.00% |
| AML 239 | Bone marrow | Diagnostic | Minus 173 | 90 | 82 | 82 | 100.0% | 150649 | 365340 | 41.24% | 3 | 3 | 3100 | 7749 | 40.01% |
| AML 239 | Bone marrow | Remission | Minus 37 | 90 | 82 | 26 | 31.7% | 485 | 927231 | 0.05% | 3 | 1 | 882 | 18589 | 4.74% |
| AML 239 | Plasma | 0 month | Minus 7 | 90 | 82 | 18 | 22.0% | 95 | 119807 | 0.08% | 3 | 1 | 139 | 3208 | 4.33% |
| AML 239 | Plasma | 2 months | 57 | 90 | 82 | 2 | 2.4% | 2 | 2026974 | 0.00% | 3 | 1 | 6 | 42486 | 0.01% |
| AML 239 | Plasma | 6 months | 181 | 90 | 82 | 0 | 0.0% | 0 | 194568 | 0.00% | 3 | 0 | 0 | 3068 | 0.00% |
| AML 239 | Plasma | 3 months | 83 | 90 | 82 | 2 | 2.4% | 2 | 334186 | 0.00% | 3 | 1 | 2 | 7982 | 0.03% |
| AML 239 | Plasma | 12 months | 349 | 90 | 82 | 3 | 3.7% | 4 | 1646012 | 0.00% | 3 | 1 | 1 | 27187 | 0.00% |
| AML 240 | Bone marrow | Diagnostic | Minus 177 | 57 | 67 | 67 | 100.0% | 271154 | 636620 | 42.59% | 2 | 2 | 2859 | 7803 | 36.64% |
| AML 240 | Bone marrow | Remission | Minus 33 | 57 | 67 | 14 | 20.9% | 20 | 331094 | 0.01% | 2 | 0 | 0 | 4701 | 0.00% |
| AML 240 | Plasma | 0 month | Minus 7 | 57 | 67 | 60 | 89.6% | 264 | 1632661 | 0.02% | 2 | 0 | 0 | 19580 | 0.00% |

| | | | | | | | | | | | | | | | |
|---|---|---|---|---|---|---|---|---|---|---|---|---|---|---|---|
| AML 240 | Plasma | 2 months | 56 | 57 | 67 | 51 | 76.1% | 129 | 2492893 | 0.01% | 2 | 0 | 0 | 30332 | 0.00% |
| AML 240 | Plasma | 3 months | 82 | 57 | 67 | 47 | 70.1% | 121 | 3009828 | 0.00% | 2 | 0 | 0 | 18209 | 0.00% |
| AML 240 | Plasma | 6 months | 173 | 57 | 67 | 19 | 28.4% | 23 | 1494422 | 0.00% | 2 | 0 | 0 | 21475 | 0.00% |
| AML 240 | Plasma | 12 months | 363 | 57 | 67 | 0 | 0.0% | 0 | 713527 | 0.00% | 2 | 0 | 0 | 10154 | 0.00% |
| AML 241 | Bone marrow | Diagnostic | Minus 475 | 60 | 80 | 80 | 100.0% | 171249 | 407242 | 42.05% | 1 | 1 | 782 | 1715 | 45.60% |
| AML 241 | Bone marrow | Remission | Minus 29 | 60 | 80 | 46 | 57.5% | 92 | 165661 | 0.06% | 1 | 1 | 1 | 636 | 0.16% |
| AML 241 | Plasma | 0 month | Minus 7 | 60 | 80 | 18 | 22.5% | 20 | 525778 | 0.00% | 1 | 0 | 0 | 3767 | 0.00% |
| AML 241 | Plasma | 2 months | 57 | 60 | 80 | 3 | 3.8% | 3 | 2869033 | 0.00% | 1 | 0 | 0 | 7441 | 0.00% |
| AML 241 | Plasma | 3 months | 92 | 60 | 80 | 2 | 2.5% | 2 | 1482407 | 0.00% | 1 | 0 | 0 | 9483 | 0.00% |
| AML 241 | Plasma | 6 months | 177 | 60 | 80 | 2 | 2.5% | 2 | 2207813 | 0.00% | 1 | 0 | 0 | 11873 | 0.00% |
| AML 241 | Plasma | 12 months | 358 | 60 | 80 | 0 | 0.0% | 0 | 842984 | 0.00% | 1 | 0 | 0 | 5304 | 0.00% |
| AML 242 | Bone marrow | Diagnostic | Minus 507 | 106 | 69 | 69 | 100.0% | 211538 | 591138 | 35.78% | 7 | 7 | 7613 | 34296 | 22.20% |
| AML 242 | Bone marrow | Remission | Minus 44 | 106 | 69 | 37 | 53.6% | 1275 | 299889 | 0.43% | 7 | 1 | 938 | 20656 | 4.54% |
| AML 242 | Plasma | 2 months | 56 | 106 | 69 | 28 | 40.6% | 240 | 4658767 | 0.01% | 7 | 1 | 85 | 187692 | 0.05% |
| AML 242 | Plasma | 3 months | 106 | 106 | 69 | 18 | 26.1% | 33 | 862469 | 0.00% | 7 | 1 | 9 | 48198 | 0.02% |
| AML 242 | Plasma | 6 months | 180 | 106 | 69 | 37 | 53.6% | 1720 | 669814 | 0.26% | 7 | 1 | 12 | 11564 | 0.10% |
| AML 242 | Plasma | 12 months | 376 | 106 | 69 | 1 | 1.4% | 1 | 1640073 | 0.00% | 7 | 0 | 0 | 88203 | 0.00% |
| AML 244 | Bone marrow | Diagnostic | Minus 149 | 60 | 83 | 83 | 100.0% | 323547 | 1335278 | 24.23% | 3 | 3 | 2737 | 25698 | 10.65% |
| AML 244 | Bone marrow | Remission | Minus 43 | 60 | 83 | 65 | 78.3% | 161 | 1145059 | 0.01% | 3 | 0 | 0 | 17160 | 0.00% |
| AML 244 | Plasma | 2 months | 66 | 60 | 83 | 81 | 97.6% | 898 | 3040498 | 0.03% | 3 | 2 | 8 | 81306 | 0.01% |
| AML 244 | Plasma | 6 months | 192 | 60 | 83 | 2 | 2.4% | 2 | 1585052 | 0.00% | 3 | 0 | 0 | 44920 | 0.00% |
| AML 244 | Plasma | 12 months | 360 | 60 | 83 | 1 | 1.2% | 1 | 858010 | 0.00% | 3 | 0 | 0 | 25457 | 0.00% |
| AML 245 | Bone marrow | Diagnostic | Minus 137 | 57 | 71 | 71 | 100.0% | 126917 | 299142 | 42.43% | 4 | 4 | 4544 | 11475 | 39.60% |
| AML 245 | Bone marrow | Remission | Minus 106 | 57 | 71 | 58 | 81.7% | 138 | 411654 | 0.03% | 4 | 1 | 1 | 14206 | 0.01% |
| AML 245 | Plasma | 0 month | Minus 7 | 57 | 71 | 66 | 93.0% | 276 | 117576 | 0.23% | 4 | 4 | 40 | 6001 | 0.67% |
| AML 245 | Plasma | 2 months | 63 | 57 | 71 | 64 | 90.1% | 243 | 80238 | 0.30% | 4 | 4 | 23 | 4081 | 0.56% |
| AML 245 | Plasma | 3 months | 92 | 57 | 71 | 4 | 5.6% | 4 | 305794 | 0.00% | 4 | 1 | 38 | 11446 | 0.33% |
| AML 246 | Bone marrow | Diagnostic | Minus 150 | 60 | 74 | 74 | 100.0% | 31694 | 498167 | 6.36% | 1 | 1 | 216 | 1997 | 10.82% |
| AML 246 | Bone marrow | Remission | Minus 65 | 60 | 74 | 46 | 62.2% | 87 | 443131 | 0.02% | 1 | 1 | 1 | 1079 | 0.09% |
| AML 246 | Plasma | 0 month | Minus 7 | 60 | 74 | 15 | 20.3% | 16 | 361933 | 0.00% | 1 | 0 | 0 | 1901 | 0.00% |
| AML 246 | Plasma | 2 months | 55 | 60 | 74 | 41 | 55.4% | 57 | 4596750 | 0.00% | 1 | 0 | 0 | 15962 | 0.00% |
| AML 246 | Plasma | 6 months | 176 | 60 | 74 | 54 | 73.0% | 128 | 2274087 | 0.01% | 1 | 0 | 0 | 9745 | 0.00% |
| AML 246 | Plasma | 12 months | 358 | 60 | 74 | 19 | 25.7% | 21 | 2181994 | 0.00% | 1 | 0 | 0 | 8410 | 0.00% |
| AML 247 | Bone marrow | Diagnostic | Minus 297 | 60 | 79 | 79 | 100.0% | 447998 | 1131065 | 39.61% | 6 | 6 | 3144 | 54325 | 5.79% |
| AML 247 | Bone marrow | Remission | Minus 161 | 60 | 79 | 9 | 11.4% | 11 | 658314 | 0.00% | 6 | 0 | 0 | 31284 | 0.00% |
| AML 247 | Plasma | 2 months | 57 | 60 | 79 | 55 | 69.6% | 137 | 171274 | 0.08% | 6 | 0 | 0 | 10549 | 0.00% |
| AML 247 | Plasma | 3 months | 90 | 60 | 79 | 71 | 89.9% | 9747 | 325407 | 3.00% | 6 | 1 | 89 | 20100 | 0.44% |
| AML 248 | Bone marrow | Diagnostic | Minus 106 | 60 | 81 | 81 | 100.0% | 290830 | 722169 | 40.27% | 1 | 1 | 18 | 7317 | 0.25% |
| AML 248 | Bone marrow | Remission | Minus 16 | 60 | 81 | 73 | 90.1% | 3053 | 779472 | 0.39% | 1 | 1 | 418 | 8095 | 5.16% |
| AML 248 | Plasma | 0 month | Minus 7 | 60 | 81 | 71 | 87.7% | 9768 | 1786852 | 0.55% | 1 | 1 | 1169 | 19534 | 5.98% |
| AML 248 | Plasma | 2 months | 58 | 60 | 81 | 69 | 85.2% | 4211 | 4496633 | 0.09% | 1 | 1 | 27 | 47151 | 0.06% |
| AML 248 | Plasma | 3 months | 86 | 60 | 81 | 60 | 74.1% | 1232 | 1132386 | 0.11% | 1 | 0 | 0 | 11932 | 0.00% |
| AML 249 | Bone marrow | Diagnostic | Minus 194 | 66 | 32 | 32 | 100.0% | 8129 | 25295 | 32.14% | 4 | 4 | 1337 | 6839 | 19.55% |
| AML 249 | Bone marrow | Remission | Minus 40 | 66 | 32 | 28 | 87.5% | 369 | 319360 | 0.12% | 4 | 3 | 35 | 26977 | 0.13% |
| AML 249 | Plasma | 2 months | 56 | 66 | 32 | 24 | 75.0% | 232 | 1283117 | 0.02% | 4 | 1 | 7 | 90536 | 0.01% |
| AML 249 | Plasma | 6 months | 175 | 66 | 32 | 3 | 9.4% | 3 | 1555988 | 0.00% | 4 | 1 | 1 | 94860 | 0.00% |
| AML 249 | Plasma | 12 months | 353 | 66 | 32 | 0 | 0.0% | 0 | 503479 | 0.00% | 4 | 0 | 0 | 38113 | 0.00% |
| AML 250 | Bone marrow | Diagnostic | Minus 126 | 70 | 81 | 81 | 100.0% | 281438 | 723737 | 38.89% | 3 | 3 | 4022 | 12116 | 33.20% |
| AML 250 | Bone marrow | Remission | Minus 92 | 70 | 81 | 35 | 43.2% | 93 | 745551 | 0.01% | 3 | 1 | 3 | 11831 | 0.03% |
| AML 250 | Plasma | 0 month | Minus 7 | 70 | 81 | 40 | 49.4% | 103 | 748942 | 0.01% | 3 | 1 | 1 | 12550 | 0.01% |
| AML 250 | Plasma | 2 months | 56 | 70 | 81 | 4 | 4.9% | 4 | 635648 | 0.00% | 3 | 0 | 0 | 10785 | 0.00% |
| AML 250 | Plasma | 3 months | 87 | 70 | 81 | 5 | 6.2% | 7 | 1150663 | 0.00% | 3 | 0 | 0 | 18241 | 0.00% |
| AML 250 | Plasma | 6 months | 172 | 70 | 81 | 5 | 6.2% | 6 | 1097866 | 0.00% | 3 | 0 | 0 | 17907 | 0.00% |
| AML 250 | Plasma | 12 months | 413 | 70 | 81 | 0 | 0.0% | 0 | 1714050 | 0.00% | 3 | 0 | 0 | 28879 | 0.00% |
| AML 251 | Bone marrow | Diagnostic | Minus 147 | 180 | 76 | 76 | 100.0% | 83473 | 351865 | 23.72% | 3 | 3 | 3207 | 13603 | 23.58% |
| AML 251 | Bone marrow | Remission | Minus 38 | 180 | 76 | 32 | 42.1% | 60 | 566576 | 0.01% | 3 | 1 | 5 | 15001 | 0.03% |
| AML 251 | Plasma | 0 month | Minus 7 | 180 | 76 | 75 | 98.7% | 1228 | 2643761 | 0.05% | 3 | 3 | 50 | 84460 | 0.06% |
| AML 251 | Plasma | 2 months | 56 | 180 | 76 | 36 | 47.4% | 53 | 4523621 | 0.00% | 3 | 1 | 2 | 139909 | 0.00% |
| AML 251 | Plasma | 3 months | 87 | 180 | 76 | 45 | 59.2% | 91 | 2822566 | 0.00% | 3 | 1 | 4 | 82156 | 0.00% |
| AML 251 | Plasma | 6 months | 176 | 180 | 76 | 5 | 6.6% | 7 | 4660989 | 0.00% | 3 | 0 | 0 | 152458 | 0.00% |
| AML 251 | Plasma | 12 months | 351 | 180 | 76 | 1 | 1.3% | 1 | 1028218 | 0.00% | 3 | 0 | 0 | 37243 | 0.00% |

*NA = data not available; PB = peripheral blood; CD3 = CD3 sorted T cells*

**Table S3: Mutations identified in paired plasma and bone marrow samples at remission and ~3 months following transplantation.**

| Patient | Mutation | Timepoint | Days post-alloHCT, plasma collection | Number of mutant molecules in plasma | Total number of molecules assessed in plasma | Mutant allele frequency in plasma | Days post-alloHCT, bone marrow collection | Number of mutant molecules in bone marrow | Total number of molecules assessed in bone marrow | Mutant allele frequency in bone marrow |
|---|---|---|---|---|---|---|---|---|---|---|
| AML 121 | chr1:110245902G>A | Remission | Minus 7 | 0 | 5204 | 0.000% | Minus 182 | 127 | 6997 | 1.815% |
| AML 121 | chr10:19159747A>G | Remission | Minus 7 | 0 | 407 | 0.000% | Minus 182 | 17 | 1096 | 1.551% |
| AML 121 | chr10:63867282G>A | Remission | Minus 7 | 68 | 5125 | 1.327% | Minus 182 | 61 | 6498 | 0.939% |
| AML 121 | chr10:75654454G>A | Remission | Minus 7 | 1 | 948 | 0.105% | Minus 182 | 26 | 2005 | 1.297% |
| AML 121 | chr11:21137971G>A | Remission | Minus 7 | 5 | 4760 | 0.105% | Minus 182 | 129 | 7643 | 1.688% |
| AML 121 | chr11:28565424C>T | Remission | Minus 7 | 17 | 4246 | 0.400% | Minus 182 | 12 | 5656 | 0.212% |
| AML 121 | chr11:41711884C>T | Remission | Minus 7 | 4 | 4080 | 0.098% | Minus 182 | 98 | 6409 | 1.529% |
| AML 121 | chr11:45984682C>T | Remission | Minus 7 | 65 | 5048 | 1.288% | Minus 182 | 71 | 7039 | 1.009% |
| AML 121 | chr12:101081568A>G | Remission | Minus 7 | 1 | 4092 | 0.024% | Minus 182 | 134 | 6147 | 2.180% |
| AML 121 | chr13:113254106G>A | Remission | Minus 7 | 72 | 5643 | 1.276% | Minus 182 | 71 | 7491 | 0.948% |
| AML 121 | chr13:35209302C>T | Remission | Minus 7 | 2 | 3471 | 0.058% | Minus 182 | 92 | 4232 | 2.174% |
| AML 121 | chr14:77501038C>T | Remission | Minus 7 | 64 | 4696 | 1.363% | Minus 182 | 86 | 7123 | 1.207% |
| AML 121 | chr15:41955386G>A | Remission | Minus 7 | 3 | 5751 | 0.052% | Minus 182 | 111 | 7682 | 1.445% |
| AML 121 | chr15:42068543C>T | Remission | Minus 7 | 1 | 6588 | 0.015% | Minus 182 | 112 | 7333 | 1.527% |
| AML 121 | chr15:59271645T>C | Remission | Minus 7 | 2 | 3236 | 0.062% | Minus 182 | 52 | 4896 | 1.062% |
| AML 121 | chr16:61878846T>G | Remission | Minus 7 | 1 | 4312 | 0.023% | Minus 182 | 89 | 6904 | 1.289% |
| AML 121 | chr16:79606865T>C | Remission | Minus 7 | 48 | 3762 | 1.276% | Minus 182 | 47 | 5303 | 0.886% |
| AML 121 | chr17:12323012T>C | Remission | Minus 7 | 2 | 4410 | 0.045% | Minus 182 | 116 | 7428 | 1.562% |
| AML 121 | chr18:33220934C>T | Remission | Minus 7 | 1 | 5154 | 0.019% | Minus 182 | 114 | 6997 | 1.629% |
| AML 121 | chr18:51651640C>G | Remission | Minus 7 | 71 | 5045 | 1.407% | Minus 182 | 67 | 6904 | 0.970% |
| AML 121 | chr2:104729661C>G | Remission | Minus 7 | 666 | 6770 | 9.838% | Minus 182 | 477 | 6757 | 7.059% |
| AML 121 | chr2:71464109C>T | Remission | Minus 7 | 42 | 3768 | 1.115% | Minus 182 | 58 | 5819 | 0.997% |
| AML 121 | chr20:57195801C>T | Remission | Minus 7 | 104 | 6100 | 1.705% | Minus 182 | 94 | 8271 | 1.137% |
| AML 121 | chr20:62775175G>A | Remission | Minus 7 | 104 | 7068 | 1.471% | Minus 182 | 96 | 9027 | 1.063% |
| AML 121 | chr3:115810896G>A | Remission | Minus 7 | 3 | 4860 | 0.062% | Minus 182 | 117 | 7111 | 1.645% |
| AML 121 | chr3:176246793T>C | Remission | Minus 7 | 2 | 3722 | 0.054% | Minus 182 | 113 | 5945 | 1.901% |
| AML 121 | chr3:94302682C>T | Remission | Minus 7 | 0 | 4229 | 0.000% | Minus 182 | 102 | 5765 | 1.769% |
| AML 121 | chr4:134547370G>C | Remission | Minus 7 | 3 | 4592 | 0.065% | Minus 182 | 53 | 6600 | 0.803% |
| AML 121 | chr4:147093498G>A | Remission | Minus 7 | 2 | 2943 | 0.068% | Minus 182 | 33 | 3370 | 0.979% |
| AML 121 | chr4:154043601G>C | Remission | Minus 7 | 1 | 5603 | 0.018% | Minus 182 | 61 | 6597 | 0.925% |
| AML 121 | chr5:15219770G>A | Remission | Minus 7 | 0 | 5108 | 0.000% | Minus 182 | 131 | 6703 | 1.954% |
| AML 121 | chr5:2216594G>A | Remission | Minus 7 | 50 | 3719 | 1.344% | Minus 182 | 50 | 4528 | 1.104% |
| AML 121 | chr5:8331558G>A | Remission | Minus 7 | 8 | 637 | 1.256% | Minus 182 | 12 | 1492 | 0.804% |
| AML 121 | chr5:89300749C>G | Remission | Minus 7 | 1 | 3874 | 0.026% | Minus 182 | 100 | 5559 | 1.799% |
| AML 121 | chr7:11262755A>C | Remission | Minus 7 | 3 | 4981 | 0.060% | Minus 182 | 42 | 6015 | 0.698% |
| AML 121 | chr7:130141280T>C | Remission | Minus 7 | 4 | 1539 | 0.260% | Minus 182 | 3 | 2907 | 0.103% |
| AML 121 | chr7:154705495C>T | Remission | Minus 7 | 18 | 5553 | 0.324% | Minus 182 | 25 | 6864 | 0.364% |
| AML 121 | chr7:35356833C>T | Remission | Minus 7 | 2 | 3430 | 0.058% | Minus 182 | 35 | 5360 | 0.653% |
| AML 121 | chr8:102896093A>G | Remission | Minus 7 | 3 | 3908 | 0.077% | Minus 182 | 42 | 5999 | 0.700% |
| AML 121 | chr8:127644825G>A | Remission | Minus 7 | 85 | 6923 | 1.228% | Minus 182 | 83 | 8411 | 0.987% |
| AML 121 | chr8:22624337T>C | Remission | Minus 7 | 16 | 1017 | 1.573% | Minus 182 | 22 | 2166 | 1.016% |
| AML 121 | chr9:135723685C>T | Remission | Minus 7 | 26 | 5384 | 0.483% | Minus 182 | 16 | 7183 | 0.223% |
| AML 121 | chr9:68940919G>C | Remission | Minus 7 | 85 | 4870 | 1.745% | Minus 182 | 72 | 6527 | 1.103% |
| AML 121 | chrX:50994220G>A | Remission | Minus 7 | 6 | 3027 | 0.198% | Minus 182 | 128 | 3923 | 3.263% |

| | | | | | | | | | | |
|---|---|---|---|---|---|---|---|---|---|---|
| AML 121 | chrX:64270054C>T | Remission | Minus 7 | 2 | 1547 | 0.129% | Minus 182 | 37 | 3058 | 1.210% |
| AML 124 | chr1:193497400C>G | Remission | Minus 7 | 1 | 2419 | 0.041% | Minus 126 | 0 | 5683 | 0.000% |
| AML 124 | chr3:191874226T>C | Remission | Minus 7 | 0 | 1899 | 0.000% | Minus 126 | 1 | 4961 | 0.020% |
| AML 126 | chr1:219518740C>G | Remission | Minus 7 | 0 | 6962 | 0.000% | Minus 107 | 5 | 7364 | 0.068% |
| AML 126 | chr10:1167403T>C | Remission | Minus 7 | 5 | 9605 | 0.052% | Minus 107 | 1 | 6756 | 0.015% |
| AML 126 | chr16:79739452A>G | Remission | Minus 7 | 0 | 966 | 0.000% | Minus 107 | 1 | 1975 | 0.051% |
| AML 126 | chr19:46015947T>C | Remission | Minus 7 | 0 | 8405 | 0.000% | Minus 107 | 1 | 8394 | 0.012% |
| AML 126 | chr2:160373535C>G | Remission | Minus 7 | 4 | 4756 | 0.084% | Minus 107 | 0 | 7446 | 0.000% |
| AML 126 | chr2:25234373C>T | Remission | Minus 7 | 4 | 10797 | 0.037% | Minus 107 | 3 | 8577 | 0.035% |
| AML 126 | chr2:60499249C>G | Remission | Minus 7 | 8 | 12400 | 0.065% | Minus 107 | 5 | 8674 | 0.058% |
| AML 126 | chr5:139697924T>G | Remission | Minus 7 | 1 | 11249 | 0.009% | Minus 107 | 2 | 7117 | 0.028% |
| AML 126 | chr5:156738149A>G | Remission | Minus 7 | 1 | 2995 | 0.033% | Minus 107 | 0 | 4196 | 0.000% |
| AML 126 | chr6:82647125A>G | Remission | Minus 7 | 1 | 1100 | 0.091% | Minus 107 | 1 | 2031 | 0.049% |
| AML 126 | chr7:154503700G>- | Remission | Minus 7 | 3 | 8651 | 0.035% | Minus 107 | 2 | 6994 | 0.029% |
| AML 126 | chr7:68016075C>G | Remission | Minus 7 | 6 | 9231 | 0.065% | Minus 107 | 4 | 7943 | 0.050% |
| AML 129 | chr10:124991078G>C | Remission | Minus 7 | 1 | 7061 | 0.014% | Minus 99 | 0 | 6849 | 0.000% |
| AML 129 | chr11:133840616A>G | Remission | Minus 7 | 1 | 6893 | 0.015% | Minus 99 | 0 | 5966 | 0.000% |
| AML 129 | chr19:10877193A>G | Remission | Minus 7 | 1 | 9546 | 0.010% | Minus 99 | 0 | 9034 | 0.000% |
| AML 129 | chr21:34880577A>C | Remission | Minus 7 | 0 | 8160 | 0.000% | Minus 99 | 1 | 9757 | 0.010% |
| AML 129 | chr4:105272753G>T | Remission | Minus 7 | 2 | 8667 | 0.023% | Minus 99 | 0 | 10015 | 0.000% |
| AML 129 | chr4:105272754C>A | Remission | Minus 7 | 2 | 8586 | 0.023% | Minus 99 | 0 | 9943 | 0.000% |
| AML 129 | chr5:173670181T>G | Remission | Minus 7 | 0 | 7083 | 0.000% | Minus 99 | 1 | 10722 | 0.009% |
| AML 129 | chr6:104047120C>G | Remission | Minus 7 | 1 | 7413 | 0.013% | Minus 99 | 1 | 10150 | 0.010% |
| AML 129 | chr6:86337232A>C | Remission | Minus 7 | 1 | 7935 | 0.013% | Minus 99 | 1 | 10019 | 0.010% |
| AML 129 | chr9:129875614A>G | Remission | Minus 7 | 1 | 9279 | 0.011% | Minus 99 | 0 | 7287 | 0.000% |
| AML 130 | chr1:106285578T>G | Remission | Minus 7 | 3 | 6302 | 0.048% | Minus 127 | 0 | 9672 | 0.000% |
| AML 130 | chr1:166135924A>G | Remission | Minus 7 | 4 | 7216 | 0.055% | Minus 127 | 1 | 8201 | 0.012% |
| AML 130 | chr10:125125474T>C | Remission | Minus 7 | 2 | 6219 | 0.032% | Minus 127 | 0 | 7788 | 0.000% |
| AML 130 | chr10:34621587A>T | Remission | Minus 7 | 334 | 6990 | 4.778% | Minus 127 | 514 | 12517 | 4.106% |
| AML 130 | chr10:85852159T>C | Remission | Minus 7 | 6 | 8882 | 0.068% | Minus 127 | 1 | 10500 | 0.010% |
| AML 130 | chr11:122692233A>G | Remission | Minus 7 | 325 | 6301 | 5.158% | Minus 127 | 368 | 8267 | 4.451% |
| AML 130 | chr11:38802150A>G | Remission | Minus 7 | 3 | 6678 | 0.045% | Minus 127 | 0 | 4780 | 0.000% |
| AML 130 | chr11:64395060A>G | Remission | Minus 7 | 2 | 5929 | 0.034% | Minus 127 | 2 | 8617 | 0.023% |
| AML 130 | chr12:104741654A>G | Remission | Minus 7 | 7 | 7523 | 0.093% | Minus 127 | 11 | 7364 | 0.149% |
| AML 130 | chr12:13896169A>G | Remission | Minus 7 | 3 | 7374 | 0.041% | Minus 127 | 0 | 9672 | 0.000% |
| AML 130 | chr12:92614953T>G | Remission | Minus 7 | 12 | 7668 | 0.156% | Minus 127 | 14 | 10509 | 0.133% |
| AML 130 | chr13:55240615T>C | Remission | Minus 7 | 2 | 6405 | 0.031% | Minus 127 | 0 | 8319 | 0.000% |
| AML 130 | chr14:26683707T>C | Remission | Minus 7 | 356 | 6216 | 5.727% | Minus 127 | 385 | 8789 | 4.380% |
| AML 130 | chr14:48973412A>G | Remission | Minus 7 | 1 | 6629 | 0.015% | Minus 127 | 0 | 9297 | 0.000% |
| AML 130 | chr14:90423441C>T | Remission | Minus 7 | 1193 | 7498 | 15.911% | Minus 127 | 2251 | 15277 | 14.735% |
| AML 130 | chr14:98255192T>C | Remission | Minus 7 | 1350 | 8521 | 15.843% | Minus 127 | 1952 | 13276 | 14.703% |
| AML 130 | chr16:46743605A>G | Remission | Minus 7 | 163 | 3242 | 5.028% | Minus 127 | 152 | 3698 | 4.110% |
| AML 130 | chr16:65528696G>C | Remission | Minus 7 | 563 | 6785 | 8.298% | Minus 127 | 990 | 12806 | 7.731% |
| AML 130 | chr17:59314673A>G | Remission | Minus 7 | 4 | 6075 | 0.066% | Minus 127 | 0 | 6309 | 0.000% |
| AML 130 | chr18:51567834A>T | Remission | Minus 7 | 42 | 6420 | 0.654% | Minus 127 | 55 | 9928 | 0.554% |
| AML 130 | chr2:119166886A>C | Remission | Minus 7 | 3 | 5311 | 0.056% | Minus 127 | 1 | 11752 | 0.009% |
| AML 130 | chr2:80091791A>G | Remission | Minus 7 | 4 | 6877 | 0.058% | Minus 127 | 2 | 9251 | 0.022% |
| AML 130 | chr20:11727205C>G | Remission | Minus 7 | 24 | 6066 | 0.396% | Minus 127 | 32 | 10081 | 0.317% |
| AML 130 | chr3:128483911->TA | Remission | Minus 7 | 1 | 7039 | 0.014% | Minus 127 | 0 | 14528 | 0.000% |
| AML 130 | chr3:66126348G>C | Remission | Minus 7 | 6 | 7792 | 0.077% | Minus 127 | 1 | 12776 | 0.008% |

| | | | | | | | | | | |
|---|---|---|---|---|---|---|---|---|---|---|
| AML 130 | chr4:64227897T>A | Remission | Minus 7 | 0 | 6023 | 0.000% | Minus 127 | 2 | 9441 | 0.021% |
| AML 130 | chr4:94032696A>G | Remission | Minus 7 | 3 | 7286 | 0.041% | Minus 127 | 0 | 8280 | 0.000% |
| AML 130 | chr5:178289367T>C | Remission | Minus 7 | 5 | 9386 | 0.053% | Minus 127 | 1 | 5948 | 0.017% |
| AML 130 | chr5:56376041C>G | Remission | Minus 7 | 25 | 8134 | 0.307% | Minus 127 | 14 | 9666 | 0.145% |
| AML 130 | chr6:2469835T>C | Remission | Minus 7 | 5 | 8899 | 0.056% | Minus 127 | 3 | 12716 | 0.024% |
| AML 130 | chr6:722843T>C | Remission | Minus 7 | 3 | 10034 | 0.030% | Minus 127 | 2 | 9509 | 0.021% |
| AML 130 | chr6:84468378G>C | Remission | Minus 7 | 1 | 6187 | 0.016% | Minus 127 | 0 | 10085 | 0.000% |
| AML 130 | chr7:11319387A>G | Remission | Minus 7 | 1 | 7663 | 0.013% | Minus 127 | 0 | 10053 | 0.000% |
| AML 130 | chr7:135976147G>C | Remission | Minus 7 | 2 | 6097 | 0.033% | Minus 127 | 1 | 9134 | 0.011% |
| AML 130 | chr9:78680128G>A | Remission | Minus 7 | 4 | 8018 | 0.050% | Minus 127 | 0 | 9861 | 0.000% |
| AML 132 | chr1:117114058A>G | Remission | Minus 7 | 510 | 6968 | 7.319% | Minus 103 | 47 | 2634 | 1.784% |
| AML 132 | chr1:14930392A>G | Remission | Minus 7 | 473 | 7448 | 6.351% | Minus 103 | 31 | 3132 | 0.990% |
| AML 132 | chr1:188189173A>G | Remission | Minus 7 | 500 | 7272 | 6.876% | Minus 103 | 73 | 4589 | 1.591% |
| AML 132 | chr1:53650861A>G | Remission | Minus 7 | 613 | 8661 | 7.078% | Minus 103 | 73 | 5438 | 1.342% |
| AML 132 | chr1:86765225T>C | Remission | Minus 7 | 609 | 8444 | 7.212% | Minus 103 | 85 | 6104 | 1.393% |
| AML 132 | chr10:127095321C>G | Remission | Minus 7 | 879 | 11512 | 7.636% | Minus 103 | 105 | 7210 | 1.456% |
| AML 132 | chr11:101322094T>C | Remission | Minus 7 | 1016 | 7446 | 13.645% | Minus 103 | 122 | 4557 | 2.677% |
| AML 132 | chr11:96259212C>G | Remission | Minus 7 | 3386 | 10724 | 31.574% | Minus 103 | 445 | 5700 | 7.807% |
| AML 132 | chr14:34361569A>G | Remission | Minus 7 | 553 | 7856 | 7.039% | Minus 103 | 67 | 5505 | 1.217% |
| AML 132 | chr14:82859772T>A | Remission | Minus 7 | 521 | 8551 | 6.093% | Minus 103 | 59 | 4952 | 1.191% |
| AML 132 | chr15:69978580A>G | Remission | Minus 7 | 700 | 9580 | 7.307% | Minus 103 | 96 | 6558 | 1.464% |
| AML 132 | chr16:54566631C>G | Remission | Minus 7 | 660 | 9126 | 7.232% | Minus 103 | 87 | 7168 | 1.214% |
| AML 132 | chr17:75109062A>G | Remission | Minus 7 | 771 | 9779 | 7.884% | Minus 103 | 176 | 6503 | 2.706% |
| AML 132 | chr17:7676011T>C | Remission | Minus 7 | 0 | 9845 | 0.000% | Minus 103 | 138 | 8019 | 1.721% |
| AML 132 | chr21:36858963T>G | Remission | Minus 7 | 630 | 9362 | 6.729% | Minus 103 | 101 | 7090 | 1.425% |
| AML 132 | chr3:150683017C>G | Remission | Minus 7 | 733 | 10241 | 7.158% | Minus 103 | 67 | 6009 | 1.115% |
| AML 132 | chr4:18228019T>A | Remission | Minus 7 | 361 | 4597 | 7.853% | Minus 103 | 41 | 2822 | 1.453% |
| AML 132 | chr4:187358097T>A | Remission | Minus 7 | 524 | 6951 | 7.538% | Minus 103 | 82 | 5621 | 1.459% |
| AML 132 | chr5:180685022C>G | Remission | Minus 7 | 639 | 8407 | 7.601% | Minus 103 | 83 | 6888 | 1.205% |
| AML 132 | chr5:30956787C>G | Remission | Minus 7 | 197 | 3676 | 5.359% | Minus 103 | 27 | 2202 | 1.226% |
| AML 132 | chr5:79668936T>C | Remission | Minus 7 | 478 | 6076 | 7.867% | Minus 103 | 54 | 3925 | 1.376% |
| AML 132 | chr5:95733819A>G | Remission | Minus 7 | 626 | 8288 | 7.553% | Minus 103 | 68 | 5372 | 1.266% |
| AML 132 | chr6:129060397A>G | Remission | Minus 7 | 488 | 7449 | 6.551% | Minus 103 | 51 | 3482 | 1.465% |
| AML 132 | chr6:14230036G>C | Remission | Minus 7 | 854 | 11514 | 7.417% | Minus 103 | 109 | 7728 | 1.410% |
| AML 132 | chr6:41418913G>C | Remission | Minus 7 | 726 | 10169 | 7.139% | Minus 103 | 84 | 5462 | 1.538% |
| AML 132 | chr7:39181631A>G | Remission | Minus 7 | 597 | 9101 | 6.560% | Minus 103 | 102 | 6667 | 1.530% |
| AML 132 | chr8:11125590A>G | Remission | Minus 7 | 179 | 9661 | 1.853% | Minus 103 | 33 | 5962 | 0.554% |
| AML 132 | chr8:35002504T>C | Remission | Minus 7 | 112 | 1874 | 5.977% | Minus 103 | 59 | 4177 | 1.412% |
| AML 132 | chr8:35006381A>G | Remission | Minus 7 | 398 | 5830 | 6.827% | Minus 103 | 61 | 4668 | 1.307% |
| AML 132 | chr8:95269782A>G | Remission | Minus 7 | 277 | 6340 | 4.369% | Minus 103 | 56 | 4460 | 1.256% |
| AML 132 | chr9:127135456A>G | Remission | Minus 7 | 605 | 8798 | 6.877% | Minus 103 | 58 | 3262 | 1.778% |
| AML 132 | chr9:79508916A>G | Remission | Minus 7 | 634 | 8992 | 7.051% | Minus 103 | 68 | 4966 | 1.369% |
| AML 132 | chr9:86678564C>G | Remission | Minus 7 | 568 | 8649 | 6.567% | Minus 103 | 81 | 5923 | 1.368% |
| AML 132 | chrX:28806706A>G | Remission | Minus 7 | 686 | 9690 | 7.079% | Minus 103 | 78 | 5400 | 1.444% |
| AML 132 | chrX:39806574T>A | Remission | Minus 7 | 402 | 6747 | 5.958% | Minus 103 | 61 | 4580 | 1.332% |
| AML 132 | chrX:44912774T>C | Remission | Minus 7 | 618 | 9381 | 6.588% | Minus 103 | 90 | 5711 | 1.576% |
| AML 134 | chr1:34198846A>G | Remission | Minus 7 | 2517 | 7093 | 35.486% | Minus 205 | 75 | 6530 | 1.149% |
| AML 134 | chr1:43339489T>C | Remission | Minus 7 | 4739 | 6747 | 70.239% | Minus 205 | 122 | 6396 | 1.907% |
| AML 134 | chr10:132641668T>A | Remission | Minus 7 | 2034 | 5714 | 35.597% | Minus 205 | 62 | 6223 | 0.996% |
| AML 134 | chr10:64087228A>G | Remission | Minus 7 | 2733 | 7456 | 36.655% | Minus 205 | 65 | 6526 | 0.996% |

| | | | | | | | | | | |
|---|---|---|---|---|---|---|---|---|---|---|
| AML 134 | chr11:126659511C>G | Remission | Minus 7 | 1826 | 5913 | 30.881% | Minus 205 | 70 | 6595 | 1.061% |
| AML 134 | chr11:30454130T>C | Remission | Minus 7 | 2111 | 6500 | 32.477% | Minus 205 | 57 | 5857 | 0.973% |
| AML 134 | chr11:84494225A>C | Remission | Minus 7 | 2314 | 6066 | 38.147% | Minus 205 | 65 | 6146 | 1.058% |
| AML 134 | chr12:12228999G>C | Remission | Minus 7 | 2387 | 6626 | 36.025% | Minus 205 | 66 | 6681 | 0.988% |
| AML 134 | chr12:17646823A>G | Remission | Minus 7 | 2525 | 6959 | 36.284% | Minus 205 | 85 | 7114 | 1.195% |
| AML 134 | chr12:30901554T>C | Remission | Minus 7 | 2299 | 6528 | 35.218% | Minus 205 | 57 | 5300 | 1.075% |
| AML 134 | chr13:20025322A>G | Remission | Minus 7 | 2338 | 6458 | 36.203% | Minus 205 | 62 | 5686 | 1.090% |
| AML 134 | chr13:41307073T>G | Remission | Minus 7 | 2155 | 5893 | 36.569% | Minus 205 | 71 | 7016 | 1.012% |
| AML 134 | chr14:31515609T>A | Remission | Minus 7 | 1876 | 5119 | 36.648% | Minus 205 | 51 | 6482 | 0.787% |
| AML 134 | chr14:82732502T>A | Remission | Minus 7 | 1883 | 5202 | 36.198% | Minus 205 | 67 | 6346 | 1.056% |
| AML 134 | chr15:95819504C>G | Remission | Minus 7 | 1906 | 5325 | 35.793% | Minus 205 | 68 | 5998 | 1.134% |
| AML 134 | chr16:27829998T>C | Remission | Minus 7 | 2474 | 6923 | 35.736% | Minus 205 | 63 | 6117 | 1.030% |
| AML 134 | chr16:49111324A>T | Remission | Minus 7 | 2701 | 7524 | 35.898% | Minus 205 | 73 | 7466 | 0.978% |
| AML 134 | chr17:62783058A>G | Remission | Minus 7 | 3034 | 7084 | 42.829% | Minus 205 | 91 | 6253 | 1.455% |
| AML 134 | chr2:110840630C>A | Remission | Minus 7 | 2634 | 7302 | 36.072% | Minus 205 | 80 | 7019 | 1.140% |
| AML 134 | chr2:136910168A>G | Remission | Minus 7 | 2196 | 5951 | 36.901% | Minus 205 | 48 | 5392 | 0.890% |
| AML 134 | chr2:150341865T>A | Remission | Minus 7 | 1608 | 4457 | 36.078% | Minus 205 | 51 | 5606 | 0.910% |
| AML 134 | chr2:177636241A>G | Remission | Minus 7 | 1942 | 5476 | 35.464% | Minus 205 | 65 | 6141 | 1.058% |
| AML 134 | chr2:191150948T>C | Remission | Minus 7 | 2119 | 6152 | 34.444% | Minus 205 | 60 | 5927 | 1.012% |
| AML 134 | chr2:192779344T>C | Remission | Minus 7 | 1640 | 4247 | 38.615% | Minus 205 | 49 | 4142 | 1.183% |
| AML 134 | chr2:206738778A>G | Remission | Minus 7 | 480 | 6493 | 7.393% | Minus 205 | 18 | 6317 | 0.285% |
| AML 134 | chr20:16765363A>G | Remission | Minus 7 | 1895 | 5309 | 35.694% | Minus 205 | 59 | 5283 | 1.117% |
| AML 134 | chr20:55996573A>T | Remission | Minus 7 | 1920 | 5176 | 37.094% | Minus 205 | 71 | 6151 | 1.154% |
| AML 134 | chr3:111893509A>G | Remission | Minus 7 | 8 | 11 | 72.727% | Minus 205 | 0 | 12 | 0.000% |
| AML 134 | chr3:146163035A>G | Remission | Minus 7 | 1781 | 5070 | 35.128% | Minus 205 | 67 | 5234 | 1.280% |
| AML 134 | chr4:163363495T>C | Remission | Minus 7 | 1539 | 4324 | 35.592% | Minus 205 | 60 | 4767 | 1.259% |
| AML 134 | chr4:180595890T>C | Remission | Minus 7 | 2602 | 6729 | 38.668% | Minus 205 | 60 | 5809 | 1.033% |
| AML 134 | chr4:2804783G>C | Remission | Minus 7 | 2832 | 7844 | 36.104% | Minus 205 | 77 | 6717 | 1.146% |
| AML 134 | chr5:15522182C>G | Remission | Minus 7 | 1011 | 4603 | 21.964% | Minus 205 | 34 | 5451 | 0.624% |
| AML 134 | chr6:107563734T>C | Remission | Minus 7 | 1619 | 4326 | 37.425% | Minus 205 | 71 | 4906 | 1.447% |
| AML 134 | chr7:28768132T>A | Remission | Minus 7 | 3928 | 8289 | 47.388% | Minus 205 | 72 | 6457 | 1.115% |
| AML 134 | chr9:27906493A>G | Remission | Minus 7 | 1877 | 5018 | 37.405% | Minus 205 | 56 | 5125 | 1.093% |
| AML 134 | chr9:70386318T>A | Remission | Minus 7 | 2020 | 5765 | 35.039% | Minus 205 | 66 | 5880 | 1.122% |
| AML 134 | chr9:81676483C>G | Remission | Minus 7 | 2057 | 5859 | 35.108% | Minus 205 | 68 | 5723 | 1.188% |
| AML 134 | chr9:92960287T>C | Remission | Minus 7 | 2506 | 6977 | 35.918% | Minus 205 | 72 | 6145 | 1.172% |
| AML 239 | chr1:110007539G>C | Remission | Minus 7 | 7 | 1507 | 0.464% | Minus 37 | 26 | 14604 | 0.178% |
| AML 239 | chr1:221759682T>C | Remission | Minus 7 | 1 | 1276 | 0.078% | Minus 37 | 7 | 10892 | 0.064% |
| AML 239 | chr1:237763374C>T | Remission | Minus 7 | 0 | 1612 | 0.000% | Minus 37 | 6 | 13152 | 0.046% |
| AML 239 | chr11:112858841G>A | Remission | Minus 7 | 0 | 1703 | 0.000% | Minus 37 | 5 | 12150 | 0.041% |
| AML 239 | chr12:115131646C>T | Remission | Minus 7 | 0 | 1278 | 0.000% | Minus 37 | 5 | 11740 | 0.043% |
| AML 239 | chr12:63013504G>A | Remission | Minus 7 | 8 | 1369 | 0.584% | Minus 37 | 21 | 11043 | 0.190% |
| AML 239 | chr12:78870588G>A | Remission | Minus 7 | 2 | 1274 | 0.157% | Minus 37 | 1 | 9825 | 0.010% |
| AML 239 | chr12:96679905G>C | Remission | Minus 7 | 3 | 1427 | 0.210% | Minus 37 | 4 | 10762 | 0.037% |
| AML 239 | chr13:99956664->T | Remission | Minus 7 | 1 | 1706 | 0.059% | Minus 37 | 6 | 15723 | 0.038% |
| AML 239 | chr16:52026432C>T | Remission | Minus 7 | 0 | 1186 | 0.000% | Minus 37 | 1 | 4710 | 0.021% |
| AML 239 | chr18:3655216G>A | Remission | Minus 7 | 0 | 1449 | 0.000% | Minus 37 | 1 | 13022 | 0.008% |
| AML 239 | chr2:232975430C>T | Remission | Minus 7 | 1 | 1261 | 0.079% | Minus 37 | 1 | 1036 | 0.097% |
| AML 239 | chr3:184236681C>T | Remission | Minus 7 | 4 | 1340 | 0.299% | Minus 37 | 23 | 11405 | 0.202% |
| AML 239 | chr3:27294537G>A | Remission | Minus 7 | 1 | 1350 | 0.074% | Minus 37 | 1 | 15432 | 0.006% |
| AML 239 | chr4:159389505A>T | Remission | Minus 7 | 7 | 1553 | 0.451% | Minus 37 | 34 | 15583 | 0.218% |

| | | | | | | | | | | |
|---|---|---|---|---|---|---|---|---|---|---|
| AML 239 | chr4:65559515A>G | Remission | Minus 7 | 1 | 1131 | 0.088% | Minus 37 | 5 | 7787 | 0.064% |
| AML 239 | chr4:71493502T>C | Remission | Minus 7 | 4 | 878 | 0.456% | Minus 37 | 17 | 12034 | 0.141% |
| AML 239 | chr5:153490230T>C | Remission | Minus 7 | 3 | 1392 | 0.216% | Minus 37 | 13 | 13141 | 0.099% |
| AML 239 | chr6:133191380C>T | Remission | Minus 7 | 0 | 1432 | 0.000% | Minus 37 | 1 | 11002 | 0.009% |
| AML 239 | chr6:70101478G>C | Remission | Minus 7 | 40 | 1056 | 3.788% | Minus 37 | 227 | 7227 | 3.141% |
| AML 239 | chr6:71924206C>T | Remission | Minus 7 | 0 | 1280 | 0.000% | Minus 37 | 5 | 11108 | 0.045% |
| AML 239 | chr8:102855596A>G | Remission | Minus 7 | 0 | 1155 | 0.000% | Minus 37 | 26 | 10709 | 0.243% |
| AML 239 | chr8:5679049T>A | Remission | Minus 7 | 2 | 1069 | 0.187% | Minus 37 | 18 | 10444 | 0.172% |
| AML 239 | chr8:98557144G>A | Remission | Minus 7 | 1 | 1491 | 0.067% | Minus 37 | 7 | 11134 | 0.063% |
| AML 239 | chr9:9417477C>T | Remission | Minus 7 | 5 | 1374 | 0.364% | Minus 37 | 11 | 9168 | 0.120% |
| AML 239 | chrX:145090722C>G | Remission | Minus 7 | 4 | 1340 | 0.299% | Minus 37 | 13 | 8214 | 0.158% |
| AML 240 | chr1:109547908GGCT>- | Remission | Minus 7 | 2 | 12715 | 0.016% | Minus 33 | 0 | 4664 | 0.000% |
| AML 240 | chr1:210812092C>T | Remission | Minus 7 | 6 | 21544 | 0.028% | Minus 33 | 0 | 2970 | 0.000% |
| AML 240 | chr1:28812945A>G | Remission | Minus 7 | 4 | 18600 | 0.022% | Minus 33 | 0 | 7524 | 0.000% |
| AML 240 | chr10:116848175G>A | Remission | Minus 7 | 1 | 28523 | 0.004% | Minus 33 | 0 | 4559 | 0.000% |
| AML 240 | chr10:13001010A>G | Remission | Minus 7 | 1 | 25956 | 0.004% | Minus 33 | 0 | 6485 | 0.000% |
| AML 240 | chr10:33978321A>T | Remission | Minus 7 | 6 | 19842 | 0.030% | Minus 33 | 0 | 3196 | 0.000% |
| AML 240 | chr10:97571292C>T | Remission | Minus 7 | 7 | 28710 | 0.024% | Minus 33 | 0 | 6828 | 0.000% |
| AML 240 | chr11:127739135G>A | Remission | Minus 7 | 4 | 21106 | 0.019% | Minus 33 | 0 | 2862 | 0.000% |
| AML 240 | chr11:33802572C>T | Remission | Minus 7 | 1 | 14303 | 0.007% | Minus 33 | 2 | 5874 | 0.034% |
| AML 240 | chr11:64766111GCGGGA>- | Remission | Minus 7 | 3 | 18043 | 0.017% | Minus 33 | 1 | 9619 | 0.010% |
| AML 240 | chr11:83954495A>- | Remission | Minus 7 | 4 | 23855 | 0.017% | Minus 33 | 0 | 1994 | 0.000% |
| AML 240 | chr11:92213273C>T | Remission | Minus 7 | 2 | 21433 | 0.009% | Minus 33 | 0 | 3486 | 0.000% |
| AML 240 | chr12:106669384A>G | Remission | Minus 7 | 5 | 27806 | 0.018% | Minus 33 | 0 | 5179 | 0.000% |
| AML 240 | chr12:115470541C>T | Remission | Minus 7 | 1 | 22933 | 0.004% | Minus 33 | 1 | 5335 | 0.019% |
| AML 240 | chr12:15164481C>T | Remission | Minus 7 | 10 | 28817 | 0.035% | Minus 33 | 1 | 4418 | 0.023% |
| AML 240 | chr13:101946254A>C | Remission | Minus 7 | 6 | 23077 | 0.026% | Minus 33 | 0 | 4033 | 0.000% |
| AML 240 | chr13:104750573G>A | Remission | Minus 7 | 4 | 21390 | 0.019% | Minus 33 | 0 | 3722 | 0.000% |
| AML 240 | chr13:32961338C>T | Remission | Minus 7 | 6 | 24491 | 0.024% | Minus 33 | 0 | 3581 | 0.000% |
| AML 240 | chr13:60030523C>T | Remission | Minus 7 | 3 | 23229 | 0.013% | Minus 33 | 1 | 2205 | 0.045% |
| AML 240 | chr13:73371923C>T | Remission | Minus 7 | 4 | 29050 | 0.014% | Minus 33 | 0 | 4701 | 0.000% |
| AML 240 | chr13:75764333C>T | Remission | Minus 7 | 3 | 24488 | 0.012% | Minus 33 | 0 | 3748 | 0.000% |
| AML 240 | chr14:24871961C>T | Remission | Minus 7 | 4 | 20495 | 0.020% | Minus 33 | 0 | 3762 | 0.000% |
| AML 240 | chr14:26199812G>A | Remission | Minus 7 | 6 | 24707 | 0.024% | Minus 33 | 0 | 4408 | 0.000% |
| AML 240 | chr14:50861796G>A | Remission | Minus 7 | 6 | 27979 | 0.021% | Minus 33 | 0 | 4386 | 0.000% |
| AML 240 | chr14:63420489T>C | Remission | Minus 7 | 2 | 21455 | 0.009% | Minus 33 | 0 | 3433 | 0.000% |
| AML 240 | chr14:69702837T>C | Remission | Minus 7 | 2 | 22798 | 0.009% | Minus 33 | 0 | 3849 | 0.000% |
| AML 240 | chr15:46572302A>T | Remission | Minus 7 | 5 | 20877 | 0.024% | Minus 33 | 0 | 5204 | 0.000% |
| AML 240 | chr16:35393916G>A | Remission | Minus 7 | 6 | 29593 | 0.020% | Minus 33 | 0 | 5788 | 0.000% |
| AML 240 | chr17:9987377T>C | Remission | Minus 7 | 6 | 25245 | 0.024% | Minus 33 | 1 | 5066 | 0.020% |
| AML 240 | chr18:29787619A>G | Remission | Minus 7 | 8 | 22765 | 0.035% | Minus 33 | 0 | 3009 | 0.000% |
| AML 240 | chr18:38657157T>C | Remission | Minus 7 | 5 | 24359 | 0.021% | Minus 33 | 0 | 3745 | 0.000% |
| AML 240 | chr18:47225717G>C | Remission | Minus 7 | 1 | 23643 | 0.004% | Minus 33 | 1 | 5840 | 0.017% |
| AML 240 | chr19:28501849G>A | Remission | Minus 7 | 4 | 19635 | 0.020% | Minus 33 | 3 | 4805 | 0.062% |
| AML 240 | chr19:31907090A>G | Remission | Minus 7 | 5 | 24369 | 0.021% | Minus 33 | 0 | 4963 | 0.000% |
| AML 240 | chr2:133680796A>T | Remission | Minus 7 | 6 | 14083 | 0.043% | Minus 33 | 0 | 2573 | 0.000% |
| AML 240 | chr2:190251432C>G | Remission | Minus 7 | 7 | 24375 | 0.029% | Minus 33 | 0 | 3723 | 0.000% |
| AML 240 | chr2:210937493T>C | Remission | Minus 7 | 5 | 23491 | 0.021% | Minus 33 | 0 | 3342 | 0.000% |
| AML 240 | chr2:229057989C>T | Remission | Minus 7 | 2 | 23430 | 0.009% | Minus 33 | 0 | 3370 | 0.000% |
| AML 240 | chr2:23320144G>C | Remission | Minus 7 | 2 | 24775 | 0.008% | Minus 33 | 1 | 5026 | 0.020% |

| | | | | | | | | | | |
|---|---|---|---|---|---|---|---|---|---|---|
| AML 240 | chr20:19049506G>C | Remission | Minus 7 | 1 | 16681 | 0.006% | Minus 33 | 0 | 4991 | 0.000% |
| AML 240 | chr20:19868114T>C | Remission | Minus 7 | 7 | 23813 | 0.029% | Minus 33 | 2 | 7012 | 0.029% |
| AML 240 | chr20:35945648G>A | Remission | Minus 7 | 1 | 30647 | 0.003% | Minus 33 | 0 | 6773 | 0.000% |
| AML 240 | chr20:38836067A>G | Remission | Minus 7 | 6 | 24637 | 0.024% | Minus 33 | 0 | 9143 | 0.000% |
| AML 240 | chr21:31807628G>A | Remission | Minus 7 | 7 | 29853 | 0.023% | Minus 33 | 1 | 5148 | 0.019% |
| AML 240 | chr22:30743396->G | Remission | Minus 7 | 4 | 24129 | 0.017% | Minus 33 | 0 | 5601 | 0.000% |
| AML 240 | chr3:59726355C>T | Remission | Minus 7 | 5 | 27261 | 0.018% | Minus 33 | 0 | 4681 | 0.000% |
| AML 240 | chr4:172581470A>G | Remission | Minus 7 | 6 | 28012 | 0.021% | Minus 33 | 0 | 4782 | 0.000% |
| AML 240 | chr4:72535542G>A | Remission | Minus 7 | 4 | 19806 | 0.020% | Minus 33 | 0 | 3127 | 0.000% |
| AML 240 | chr5:120081430T>C | Remission | Minus 7 | 6 | 19239 | 0.031% | Minus 33 | 0 | 2042 | 0.000% |
| AML 240 | chr5:36345987C>A | Remission | Minus 7 | 3 | 22655 | 0.013% | Minus 33 | 0 | 3719 | 0.000% |
| AML 240 | chr5:4175162C>T | Remission | Minus 7 | 5 | 25935 | 0.019% | Minus 33 | 1 | 5663 | 0.018% |
| AML 240 | chr6:110002085C>G | Remission | Minus 7 | 2 | 24900 | 0.008% | Minus 33 | 0 | 5041 | 0.000% |
| AML 240 | chr6:148399671C>T | Remission | Minus 7 | 2 | 22086 | 0.009% | Minus 33 | 0 | 3775 | 0.000% |
| AML 240 | chr7:25421764T>C | Remission | Minus 7 | 5 | 21007 | 0.024% | Minus 33 | 0 | 4038 | 0.000% |
| AML 240 | chr7:5747470T>C | Remission | Minus 7 | 10 | 20119 | 0.050% | Minus 33 | 0 | 3633 | 0.000% |
| AML 240 | chr8:104796591C>T | Remission | Minus 7 | 8 | 22540 | 0.035% | Minus 33 | 0 | 4022 | 0.000% |
| AML 240 | chr8:2973560G>A | Remission | Minus 7 | 4 | 27599 | 0.014% | Minus 33 | 0 | 4384 | 0.000% |
| AML 240 | chr8:34586152A>G | Remission | Minus 7 | 5 | 20540 | 0.024% | Minus 33 | 1 | 3351 | 0.030% |
| AML 240 | chrX:108962525C>T | Remission | Minus 7 | 3 | 13202 | 0.023% | Minus 33 | 0 | 1880 | 0.000% |
| AML 240 | chrX:134713782G>A | Remission | Minus 7 | 5 | 15245 | 0.033% | Minus 33 | 3 | 3537 | 0.085% |
| AML 241 | chr1:155187917C>T | Remission | Minus 7 | 1 | 7418 | 0.013% | Minus 29 | 0 | 2452 | 0.000% |
| AML 241 | chr1:167510196C>T | Remission | Minus 7 | 0 | 6082 | 0.000% | Minus 29 | 1 | 2514 | 0.040% |
| AML 241 | chr1:77979302G>A | Remission | Minus 7 | 0 | 4296 | 0.000% | Minus 29 | 2 | 4093 | 0.049% |
| AML 241 | chr1:77986583C>T | Remission | Minus 7 | 0 | 5524 | 0.000% | Minus 29 | 2 | 1488 | 0.134% |
| AML 241 | chr10:117555749C>G | Remission | Minus 7 | 0 | 5823 | 0.000% | Minus 29 | 2 | 2781 | 0.072% |
| AML 241 | chr10:14845383G>A | Remission | Minus 7 | 1 | 7060 | 0.014% | Minus 29 | 0 | 2128 | 0.000% |
| AML 241 | chr10:69689216T>C | Remission | Minus 7 | 0 | 7342 | 0.000% | Minus 29 | 4 | 1796 | 0.223% |
| AML 241 | chr10:7143027C>T | Remission | Minus 7 | 1 | 7419 | 0.013% | Minus 29 | 1 | 2519 | 0.040% |
| AML 241 | chr10:98468128A>G | Remission | Minus 7 | 1 | 6512 | 0.015% | Minus 29 | 2 | 3757 | 0.053% |
| AML 241 | chr11:115157549C>T | Remission | Minus 7 | 0 | 6395 | 0.000% | Minus 29 | 1 | 2621 | 0.038% |
| AML 241 | chr11:121134967G>C | Remission | Minus 7 | 0 | 6936 | 0.000% | Minus 29 | 1 | 2194 | 0.046% |
| AML 241 | chr11:130723284T>C | Remission | Minus 7 | 0 | 6998 | 0.000% | Minus 29 | 2 | 2208 | 0.091% |
| AML 241 | chr11:42317219C>T | Remission | Minus 7 | 0 | 7235 | 0.000% | Minus 29 | 1 | 2923 | 0.034% |
| AML 241 | chr11:61360611C>T | Remission | Minus 7 | 1 | 6535 | 0.015% | Minus 29 | 2 | 2610 | 0.077% |
| AML 241 | chr13:42762113T>G | Remission | Minus 7 | 0 | 6851 | 0.000% | Minus 29 | 2 | 1786 | 0.112% |
| AML 241 | chr14:103734851A>G | Remission | Minus 7 | 1 | 7299 | 0.014% | Minus 29 | 3 | 3510 | 0.085% |
| AML 241 | chr15:40955538G>A | Remission | Minus 7 | 0 | 6321 | 0.000% | Minus 29 | 4 | 3352 | 0.119% |
| AML 241 | chr15:63328667A>G | Remission | Minus 7 | 0 | 5760 | 0.000% | Minus 29 | 4 | 3417 | 0.117% |
| AML 241 | chr15:87942979G>A | Remission | Minus 7 | 0 | 7529 | 0.000% | Minus 29 | 1 | 1948 | 0.051% |
| AML 241 | chr16:50046356T>A | Remission | Minus 7 | 1 | 6283 | 0.016% | Minus 29 | 2 | 2718 | 0.074% |
| AML 241 | chr16:56267079C>T | Remission | Minus 7 | 0 | 7487 | 0.000% | Minus 29 | 2 | 2336 | 0.086% |
| AML 241 | chr16:65033077C>T | Remission | Minus 7 | 0 | 6474 | 0.000% | Minus 29 | 2 | 1175 | 0.170% |
| AML 241 | chr16:67989741->A | Remission | Minus 7 | 1 | 7423 | 0.013% | Minus 29 | 0 | 2156 | 0.000% |
| AML 241 | chr16:7397614T>G | Remission | Minus 7 | 0 | 5518 | 0.000% | Minus 29 | 1 | 954 | 0.105% |
| AML 241 | chr16:89873831G>A | Remission | Minus 7 | 0 | 2283 | 0.000% | Minus 29 | 7 | 6920 | 0.101% |
| AML 241 | chr2:75944816C>T | Remission | Minus 7 | 1 | 4924 | 0.020% | Minus 29 | 3 | 1537 | 0.195% |
| AML 241 | chr20:2527620A>G | Remission | Minus 7 | 0 | 6526 | 0.000% | Minus 29 | 1 | 1731 | 0.058% |
| AML 241 | chr20:50781999C>T | Remission | Minus 7 | 0 | 4392 | 0.000% | Minus 29 | 1 | 1767 | 0.057% |
| AML 241 | chr21:31966916G>T | Remission | Minus 7 | 1 | 6368 | 0.016% | Minus 29 | 0 | 1453 | 0.000% |

| | | | | | | | | | | |
|---|---|---|---|---|---|---|---|---|---|---|
| AML 241 | chr3:100352613C>G | Remission | Minus 7 | 0 | 5934 | 0.000% | Minus 29 | 3 | 1721 | 0.174% |
| AML 241 | chr3:25415772G>A | Remission | Minus 7 | 1 | 6190 | 0.016% | Minus 29 | 0 | 846 | 0.000% |
| AML 241 | chr3:74439657C>T | Remission | Minus 7 | 1 | 6849 | 0.015% | Minus 29 | 0 | 1113 | 0.000% |
| AML 241 | chr4:17013952C>T | Remission | Minus 7 | 0 | 7645 | 0.000% | Minus 29 | 2 | 1613 | 0.124% |
| AML 241 | chr4:172813639G>C | Remission | Minus 7 | 1 | 7496 | 0.013% | Minus 29 | 0 | 1635 | 0.000% |
| AML 241 | chr4:20946882A>C | Remission | Minus 7 | 0 | 4648 | 0.000% | Minus 29 | 1 | 872 | 0.115% |
| AML 241 | chr4:24124142T>C | Remission | Minus 7 | 1 | 6248 | 0.016% | Minus 29 | 1 | 1787 | 0.056% |
| AML 241 | chr4:40348954T>A | Remission | Minus 7 | 0 | 7629 | 0.000% | Minus 29 | 3 | 1906 | 0.157% |
| AML 241 | chr4:73862187G>A | Remission | Minus 7 | 0 | 7458 | 0.000% | Minus 29 | 1 | 1664 | 0.060% |
| AML 241 | chr5:42508532T>C | Remission | Minus 7 | 1 | 7419 | 0.013% | Minus 29 | 1 | 3934 | 0.025% |
| AML 241 | chr5:7758962C>G | Remission | Minus 7 | 2 | 7607 | 0.026% | Minus 29 | 2 | 2168 | 0.092% |
| AML 241 | chr6:118973072T>C | Remission | Minus 7 | 0 | 6337 | 0.000% | Minus 29 | 2 | 1388 | 0.144% |
| AML 241 | chr6:11938765C>G | Remission | Minus 7 | 0 | 8491 | 0.000% | Minus 29 | 2 | 2320 | 0.086% |
| AML 241 | chr6:134573972G>A | Remission | Minus 7 | 0 | 6854 | 0.000% | Minus 29 | 1 | 1262 | 0.079% |
| AML 241 | chr6:148002316T>C | Remission | Minus 7 | 1 | 5384 | 0.019% | Minus 29 | 1 | 939 | 0.106% |
| AML 241 | chr7:43550854T>C | Remission | Minus 7 | 0 | 6044 | 0.000% | Minus 29 | 2 | 1831 | 0.109% |
| AML 241 | chr8:132863794->AATC | Remission | Minus 7 | 0 | 6101 | 0.000% | Minus 29 | 2 | 2531 | 0.079% |
| AML 241 | chr8:137985582C>T | Remission | Minus 7 | 0 | 6637 | 0.000% | Minus 29 | 1 | 2119 | 0.047% |
| AML 241 | chr9:11704606C>T | Remission | Minus 7 | 2 | 7776 | 0.026% | Minus 29 | 1 | 2782 | 0.036% |
| AML 241 | chr9:125726985C>T | Remission | Minus 7 | 0 | 5846 | 0.000% | Minus 29 | 1 | 1036 | 0.097% |
| AML 241 | chr9:610949G>C | Remission | Minus 7 | 0 | 7402 | 0.000% | Minus 29 | 2 | 1769 | 0.113% |
| AML 241 | chr9:70944583G>A | Remission | Minus 7 | 0 | 6524 | 0.000% | Minus 29 | 3 | 1551 | 0.193% |
| AML 241 | chrX:136907566G>A | Remission | Minus 7 | 0 | 2926 | 0.000% | Minus 29 | 2 | 846 | 0.236% |
| AML 241 | chrX:29787252G>A | Remission | Minus 7 | 0 | 2997 | 0.000% | Minus 29 | 4 | 798 | 0.501% |
| AML 245 | chr1:152627679C>T | Remission | Minus 7 | 4 | 1576 | 0.254% | Minus 106 | 1 | 4202 | 0.024% |
| AML 245 | chr10:128489887A>T | Remission | Minus 7 | 4 | 2008 | 0.199% | Minus 106 | 4 | 6806 | 0.059% |
| AML 245 | chr10:64219533A>G | Remission | Minus 7 | 3 | 1358 | 0.221% | Minus 106 | 1 | 4598 | 0.022% |
| AML 245 | chr11:101016121A>G | Remission | Minus 7 | 6 | 1814 | 0.331% | Minus 106 | 4 | 4607 | 0.087% |
| AML 245 | chr11:115142013G>A | Remission | Minus 7 | 1 | 1742 | 0.057% | Minus 106 | 3 | 7765 | 0.039% |
| AML 245 | chr11:134426026C>T | Remission | Minus 7 | 5 | 1443 | 0.347% | Minus 106 | 2 | 6358 | 0.031% |
| AML 245 | chr11:23631913T>A | Remission | Minus 7 | 3 | 1504 | 0.199% | Minus 106 | 0 | 3570 | 0.000% |
| AML 245 | chr11:27580616C>T | Remission | Minus 7 | 4 | 1572 | 0.254% | Minus 106 | 2 | 5436 | 0.037% |
| AML 245 | chr11:32396398G>- | Remission | Minus 7 | 4 | 1930 | 0.207% | Minus 106 | 0 | 7555 | 0.000% |
| AML 245 | chr11:57304369G>A | Remission | Minus 7 | 0 | 1564 | 0.000% | Minus 106 | 4 | 8208 | 0.049% |
| AML 245 | chr11:60890916C>A | Remission | Minus 7 | 2 | 1448 | 0.138% | Minus 106 | 5 | 6641 | 0.075% |
| AML 245 | chr11:7763661G>A | Remission | Minus 7 | 7 | 1837 | 0.381% | Minus 106 | 1 | 3147 | 0.032% |
| AML 245 | chr12:104348944T>C | Remission | Minus 7 | 4 | 1660 | 0.241% | Minus 106 | 1 | 5722 | 0.017% |
| AML 245 | chr12:42857616T>G | Remission | Minus 7 | 0 | 1084 | 0.000% | Minus 106 | 2 | 3657 | 0.055% |
| AML 245 | chr12:43511398C>T | Remission | Minus 7 | 7 | 1544 | 0.453% | Minus 106 | 1 | 5000 | 0.020% |
| AML 245 | chr13:95797300G>A | Remission | Minus 7 | 2 | 1543 | 0.130% | Minus 106 | 1 | 3565 | 0.028% |
| AML 245 | chr14:95613620G>A | Remission | Minus 7 | 0 | 1632 | 0.000% | Minus 106 | 2 | 5723 | 0.035% |
| AML 245 | chr15:46780395C>G | Remission | Minus 7 | 2 | 1270 | 0.157% | Minus 106 | 2 | 4316 | 0.046% |
| AML 245 | chr16:48789182C>G | Remission | Minus 7 | 5 | 1622 | 0.308% | Minus 106 | 3 | 6192 | 0.048% |
| AML 245 | chr16:53732716ATAATA>- | Remission | Minus 7 | 4 | 1456 | 0.275% | Minus 106 | 0 | 5829 | 0.000% |
| AML 245 | chr16:53947425A>G | Remission | Minus 7 | 5 | 1607 | 0.311% | Minus 106 | 1 | 4912 | 0.020% |
| AML 245 | chr16:61077356T>A | Remission | Minus 7 | 5 | 1661 | 0.301% | Minus 106 | 1 | 3883 | 0.026% |
| AML 245 | chr16:63573629A>G | Remission | Minus 7 | 3 | 1459 | 0.206% | Minus 106 | 1 | 3226 | 0.031% |
| AML 245 | chr16:65028184C>T | Remission | Minus 7 | 5 | 1545 | 0.324% | Minus 106 | 3 | 5977 | 0.050% |
| AML 245 | chr16:66199465C>T | Remission | Minus 7 | 2 | 531 | 0.377% | Minus 106 | 2 | 4667 | 0.043% |
| AML 245 | chr16:70691086G>C | Remission | Minus 7 | 4 | 1851 | 0.216% | Minus 106 | 2 | 6925 | 0.029% |

| | | | | | | | | | | |
|---|---|---|---|---|---|---|---|---|---|---|
| AML 245 | chr17:69907658C>- | Remission | Minus 7 | 2 | 849 | 0.236% | Minus 106 | 4 | 5647 | 0.071% |
| AML 245 | chr18:53489923A>G | Remission | Minus 7 | 5 | 1201 | 0.416% | Minus 106 | 0 | 3827 | 0.000% |
| AML 245 | chr19:49148410C>T | Remission | Minus 7 | 2 | 1629 | 0.123% | Minus 106 | 2 | 5906 | 0.034% |
| AML 245 | chr2:104124163A>T | Remission | Minus 7 | 6 | 1621 | 0.370% | Minus 106 | 1 | 3814 | 0.026% |
| AML 245 | chr2:124516180T>C | Remission | Minus 7 | 7 | 1583 | 0.442% | Minus 106 | 2 | 4230 | 0.047% |
| AML 245 | chr2:211282603G>C | Remission | Minus 7 | 4 | 1640 | 0.244% | Minus 106 | 1 | 3950 | 0.025% |
| AML 245 | chr2:229170031C>T | Remission | Minus 7 | 2 | 1609 | 0.124% | Minus 106 | 3 | 6555 | 0.046% |
| AML 245 | chr2:237737312TT>- | Remission | Minus 7 | 5 | 1812 | 0.276% | Minus 106 | 3 | 7500 | 0.040% |
| AML 245 | chr2:27139588T>A | Remission | Minus 7 | 4 | 1679 | 0.238% | Minus 106 | 8 | 6123 | 0.131% |
| AML 245 | chr2:51368616C>G | Remission | Minus 7 | 4 | 1510 | 0.265% | Minus 106 | 2 | 4024 | 0.050% |
| AML 245 | chr20:20833493G>A | Remission | Minus 7 | 5 | 1909 | 0.262% | Minus 106 | 2 | 7715 | 0.026% |
| AML 245 | chr20:396185C>G | Remission | Minus 7 | 4 | 1207 | 0.331% | Minus 106 | 1 | 4969 | 0.020% |
| AML 245 | chr20:57951388G>C | Remission | Minus 7 | 5 | 1128 | 0.443% | Minus 106 | 4 | 6405 | 0.062% |
| AML 245 | chr3:116051378T>C | Remission | Minus 7 | 2 | 1518 | 0.132% | Minus 106 | 2 | 4766 | 0.042% |
| AML 245 | chr3:140493450A>G | Remission | Minus 7 | 7 | 1423 | 0.492% | Minus 106 | 0 | 4013 | 0.000% |
| AML 245 | chr3:15070588C>G | Remission | Minus 7 | 3 | 1530 | 0.196% | Minus 106 | 1 | 4924 | 0.020% |
| AML 245 | chr3:85935791C>T | Remission | Minus 7 | 0 | 1151 | 0.000% | Minus 106 | 2 | 2515 | 0.080% |
| AML 245 | chr3:9808903T>C | Remission | Minus 7 | 6 | 1524 | 0.394% | Minus 106 | 4 | 6662 | 0.060% |
| AML 245 | chr4:107434165T>G | Remission | Minus 7 | 2 | 1756 | 0.114% | Minus 106 | 3 | 4070 | 0.074% |
| AML 245 | chr4:119684667T>C | Remission | Minus 7 | 3 | 1349 | 0.222% | Minus 106 | 0 | 4465 | 0.000% |
| AML 245 | chr4:13280547C>T | Remission | Minus 7 | 2 | 1548 | 0.129% | Minus 106 | 1 | 6585 | 0.015% |
| AML 245 | chr4:138023841C>T | Remission | Minus 7 | 5 | 1407 | 0.355% | Minus 106 | 0 | 3557 | 0.000% |
| AML 245 | chr4:49002802G>A | Remission | Minus 7 | 5 | 1562 | 0.320% | Minus 106 | 2 | 4899 | 0.041% |
| AML 245 | chr4:666408G>A | Remission | Minus 7 | 6 | 1479 | 0.406% | Minus 106 | 5 | 8621 | 0.058% |
| AML 245 | chr4:94055532C>T | Remission | Minus 7 | 6 | 1499 | 0.400% | Minus 106 | 0 | 4851 | 0.000% |
| AML 245 | chr5:140005751T>C | Remission | Minus 7 | 7 | 1830 | 0.383% | Minus 106 | 2 | 6954 | 0.029% |
| AML 245 | chr5:153394254T>C | Remission | Minus 7 | 6 | 1378 | 0.435% | Minus 106 | 3 | 4278 | 0.070% |
| AML 245 | chr5:16077151T>G | Remission | Minus 7 | 1 | 1737 | 0.058% | Minus 106 | 1 | 3951 | 0.025% |
| AML 245 | chr5:68736259T>C | Remission | Minus 7 | 2 | 1923 | 0.104% | Minus 106 | 3 | 6906 | 0.043% |
| AML 245 | chr5:75804472C>- | Remission | Minus 7 | 6 | 1578 | 0.380% | Minus 106 | 0 | 5067 | 0.000% |
| AML 245 | chr6:7161727C>T | Remission | Minus 7 | 6 | 1711 | 0.351% | Minus 106 | 3 | 6137 | 0.049% |
| AML 245 | chr6:76697773T>C | Remission | Minus 7 | 2 | 1157 | 0.173% | Minus 106 | 2 | 3534 | 0.057% |
| AML 245 | chr6:9343408G>A | Remission | Minus 7 | 6 | 1549 | 0.387% | Minus 106 | 4 | 6710 | 0.060% |
| AML 245 | chr7:121676280C>T | Remission | Minus 7 | 3 | 1494 | 0.201% | Minus 106 | 2 | 4674 | 0.043% |
| AML 245 | chr7:156002360C>T | Remission | Minus 7 | 1 | 1829 | 0.055% | Minus 106 | 2 | 6466 | 0.031% |
| AML 245 | chr7:28631844G>A | Remission | Minus 7 | 7 | 1548 | 0.452% | Minus 106 | 0 | 7569 | 0.000% |
| AML 245 | chr7:68856218A>- | Remission | Minus 7 | 6 | 1263 | 0.475% | Minus 106 | 2 | 3883 | 0.052% |
| AML 245 | chr8:122848836G>A | Remission | Minus 7 | 7 | 1699 | 0.412% | Minus 106 | 1 | 8800 | 0.011% |
| AML 245 | chr8:139021656T>C | Remission | Minus 7 | 2 | 1594 | 0.125% | Minus 106 | 1 | 5097 | 0.020% |
| AML 245 | chr8:53174237C>T | Remission | Minus 7 | 4 | 1425 | 0.281% | Minus 106 | 2 | 5337 | 0.037% |
| AML 245 | chr8:55269024C>T | Remission | Minus 7 | 4 | 1346 | 0.297% | Minus 106 | 0 | 3580 | 0.000% |
| AML 245 | chr9:117099481A>T | Remission | Minus 7 | 5 | 1519 | 0.329% | Minus 106 | 0 | 4133 | 0.000% |
| AML 245 | chr9:86116555T>- | Remission | Minus 7 | 2 | 1932 | 0.104% | Minus 106 | 3 | 6811 | 0.044% |
| AML 245 | chrX:11582347A>G | Remission | Minus 7 | 6 | 811 | 0.740% | Minus 106 | 5 | 2110 | 0.237% |
| AML 246 | chr1:116694332G>A | Remission | Minus 7 | 0 | 6023 | 0.000% | Minus 65 | 3 | 6824 | 0.044% |
| AML 246 | chr1:203507528G>A | Remission | Minus 7 | 0 | 3666 | 0.000% | Minus 65 | 2 | 7957 | 0.025% |
| AML 246 | chr1:217913065G>A | Remission | Minus 7 | 0 | 5789 | 0.000% | Minus 65 | 1 | 5498 | 0.018% |
| AML 246 | chr1:228108394C>A | Remission | Minus 7 | 0 | 4047 | 0.000% | Minus 65 | 1 | 7580 | 0.013% |
| AML 246 | chr1:228816128C>T | Remission | Minus 7 | 0 | 5187 | 0.000% | Minus 65 | 1 | 5302 | 0.019% |
| AML 246 | chr1:36661813C>T | Remission | Minus 7 | 1 | 4602 | 0.022% | Minus 65 | 2 | 5360 | 0.037% |

| | | | | | | | | | | |
|---|---|---|---|---|---|---|---|---|---|---|
| AML 246 | chr1:5713062G>A | Remission | Minus 7 | 1 | 5596 | 0.018% | Minus 65 | 3 | 7212 | 0.042% |
| AML 246 | chr1:95368304G>A | Remission | Minus 7 | 1 | 4618 | 0.022% | Minus 65 | 0 | 5278 | 0.000% |
| AML 246 | chr11:131422364C>T | Remission | Minus 7 | 0 | 5315 | 0.000% | Minus 65 | 3 | 6255 | 0.048% |
| AML 246 | chr11:74574651C>T | Remission | Minus 7 | 0 | 3830 | 0.000% | Minus 65 | 1 | 4820 | 0.021% |
| AML 246 | chr11:87718570A>T | Remission | Minus 7 | 0 | 3731 | 0.000% | Minus 65 | 2 | 4316 | 0.046% |
| AML 246 | chr11:9087665C>T | Remission | Minus 7 | 0 | 4412 | 0.000% | Minus 65 | 3 | 6011 | 0.050% |
| AML 246 | chr11:92984341C>T | Remission | Minus 7 | 1 | 4807 | 0.021% | Minus 65 | 1 | 4892 | 0.020% |
| AML 246 | chr12:106579286C>T | Remission | Minus 7 | 1 | 5036 | 0.020% | Minus 65 | 3 | 6740 | 0.045% |
| AML 246 | chr13:101079503C>T | Remission | Minus 7 | 0 | 5334 | 0.000% | Minus 65 | 3 | 4615 | 0.065% |
| AML 246 | chr13:49537117A>T | Remission | Minus 7 | 0 | 4153 | 0.000% | Minus 65 | 2 | 4297 | 0.047% |
| AML 246 | chr15:100374022C>T | Remission | Minus 7 | 0 | 2958 | 0.000% | Minus 65 | 4 | 6595 | 0.061% |
| AML 246 | chr15:101307901C>T | Remission | Minus 7 | 1 | 4024 | 0.025% | Minus 65 | 3 | 2757 | 0.109% |
| AML 246 | chr15:93674127G>A | Remission | Minus 7 | 1 | 4503 | 0.022% | Minus 65 | 2 | 6509 | 0.031% |
| AML 246 | chr15:96286146G>A | Remission | Minus 7 | 1 | 5376 | 0.019% | Minus 65 | 2 | 5890 | 0.034% |
| AML 246 | chr16:27521585C>T | Remission | Minus 7 | 0 | 5162 | 0.000% | Minus 65 | 1 | 7306 | 0.014% |
| AML 246 | chr16:30513071G>A | Remission | Minus 7 | 2 | 5419 | 0.037% | Minus 65 | 1 | 6329 | 0.016% |
| AML 246 | chr16:4831053A>T | Remission | Minus 7 | 0 | 3682 | 0.000% | Minus 65 | 3 | 5686 | 0.053% |
| AML 246 | chr16:52601311T>C | Remission | Minus 7 | 0 | 3641 | 0.000% | Minus 65 | 2 | 4471 | 0.045% |
| AML 246 | chr16:62009465T>C | Remission | Minus 7 | 0 | 4363 | 0.000% | Minus 65 | 1 | 4776 | 0.021% |
| AML 246 | chr16:66245301C>T | Remission | Minus 7 | 0 | 4986 | 0.000% | Minus 65 | 4 | 7406 | 0.054% |
| AML 246 | chr16:66292578C>T | Remission | Minus 7 | 1 | 4713 | 0.021% | Minus 65 | 0 | 7089 | 0.000% |
| AML 246 | chr17:17465118G>A | Remission | Minus 7 | 0 | 5152 | 0.000% | Minus 65 | 1 | 5796 | 0.017% |
| AML 246 | chr17:33815526G>A | Remission | Minus 7 | 0 | 4933 | 0.000% | Minus 65 | 1 | 6394 | 0.016% |
| AML 246 | chr17:7675124T>C | Remission | Minus 7 | 0 | 5420 | 0.000% | Minus 65 | 2 | 9169 | 0.022% |
| AML 246 | hr17:79394816CGTCCCGGGC | Remission | Minus 7 | 0 | 4301 | 0.000% | Minus 65 | 1 | 7058 | 0.014% |
| AML 246 | chr18:32912067G>A | Remission | Minus 7 | 0 | 6312 | 0.000% | Minus 65 | 1 | 5872 | 0.017% |
| AML 246 | chr18:63295797C>T | Remission | Minus 7 | 0 | 5417 | 0.000% | Minus 65 | 1 | 6071 | 0.016% |
| AML 246 | chr2:133732853C>G | Remission | Minus 7 | 1 | 5516 | 0.018% | Minus 65 | 1 | 4675 | 0.021% |
| AML 246 | chr2:191837512C>T | Remission | Minus 7 | 1 | 4694 | 0.021% | Minus 65 | 0 | 6654 | 0.000% |
| AML 246 | chr2:201778150C>T | Remission | Minus 7 | 0 | 4012 | 0.000% | Minus 65 | 1 | 4718 | 0.021% |
| AML 246 | chr2:57406280G>A | Remission | Minus 7 | 0 | 4073 | 0.000% | Minus 65 | 2 | 4808 | 0.042% |
| AML 246 | chr2:81621992C>A | Remission | Minus 7 | 0 | 3727 | 0.000% | Minus 65 | 1 | 4585 | 0.022% |
| AML 246 | chr3:122980124C>T | Remission | Minus 7 | 0 | 5650 | 0.000% | Minus 65 | 2 | 6859 | 0.029% |
| AML 246 | chr3:13665878C>T | Remission | Minus 7 | 0 | 5773 | 0.000% | Minus 65 | 1 | 9553 | 0.010% |
| AML 246 | chr4:153936336G>A | Remission | Minus 7 | 0 | 4354 | 0.000% | Minus 65 | 2 | 5720 | 0.035% |
| AML 246 | chr7:132547631G>A | Remission | Minus 7 | 1 | 5676 | 0.018% | Minus 65 | 0 | 7594 | 0.000% |
| AML 246 | chr9:107074661C>A | Remission | Minus 7 | 0 | 4336 | 0.000% | Minus 65 | 2 | 4984 | 0.040% |
| AML 246 | chr9:138075085G>A | Remission | Minus 7 | 0 | 4387 | 0.000% | Minus 65 | 1 | 5428 | 0.018% |
| AML 246 | chr9:28394211T>A | Remission | Minus 7 | 1 | 4283 | 0.023% | Minus 65 | 2 | 4492 | 0.045% |
| AML 246 | chr9:70665332C>T | Remission | Minus 7 | 0 | 4586 | 0.000% | Minus 65 | 2 | 4068 | 0.049% |
| AML 246 | chr9:91843707G>A | Remission | Minus 7 | 0 | 5685 | 0.000% | Minus 65 | 1 | 7756 | 0.013% |
| AML 246 | chrX:132804311C>G | Remission | Minus 7 | 0 | 3063 | 0.000% | Minus 65 | 4 | 3561 | 0.112% |
| AML 246 | chrX:141217201T>A | Remission | Minus 7 | 0 | 2586 | 0.000% | Minus 65 | 2 | 3073 | 0.065% |
| AML 246 | chrX:17170935CCT>- | Remission | Minus 7 | 1 | 2598 | 0.038% | Minus 65 | 0 | 2730 | 0.000% |
| AML 246 | chrX:53217773C>A | Remission | Minus 7 | 0 | 2603 | 0.000% | Minus 65 | 2 | 3533 | 0.057% |
| AML 248 | chr1:14609249G>A | Remission | Minus 7 | 121 | 21032 | 0.575% | Minus 16 | 39 | 10313 | 0.378% |
| AML 248 | chr1:147656369C>T | Remission | Minus 7 | 278 | 22173 | 1.254% | Minus 16 | 66 | 7668 | 0.861% |
| AML 248 | chr1:203737152A>G | Remission | Minus 7 | 185 | 16969 | 1.090% | Minus 16 | 36 | 4145 | 0.869% |
| AML 248 | chr1:2820056G>A | Remission | Minus 7 | 53 | 22852 | 0.232% | Minus 16 | 6 | 12583 | 0.048% |
| AML 248 | chr1:40866191T>C | Remission | Minus 7 | 552 | 23906 | 2.309% | Minus 16 | 229 | 11033 | 2.076% |

| | | | | | | | | | | |
|---|---|---|---|---|---|---|---|---|---|---|
| AML 248 | chr1:7466894C>T | Remission | Minus 7 | 48 | 25539 | 0.188% | Minus 16 | 15 | 12228 | 0.123% |
| AML 248 | chr10:118507409T>C | Remission | Minus 7 | 48 | 19785 | 0.243% | Minus 16 | 7 | 9126 | 0.077% |
| AML 248 | chr10:16313939T>A | Remission | Minus 7 | 54 | 18361 | 0.294% | Minus 16 | 6 | 8097 | 0.074% |
| AML 248 | chr10:17429036->A | Remission | Minus 7 | 1 | 12801 | 0.008% | Minus 16 | 0 | 9575 | 0.000% |
| AML 248 | chr10:70267661C>T | Remission | Minus 7 | 3 | 20942 | 0.014% | Minus 16 | 0 | 10402 | 0.000% |
| AML 248 | chr11:11641040G>A | Remission | Minus 7 | 53 | 22908 | 0.231% | Minus 16 | 9 | 10830 | 0.083% |
| AML 248 | hr11:134654301->GGGTGCAT | Remission | Minus 7 | 176 | 21530 | 0.817% | Minus 16 | 71 | 11502 | 0.617% |
| AML 248 | chr11:36861290G>A | Remission | Minus 7 | 576 | 16638 | 3.462% | Minus 16 | 189 | 6111 | 3.093% |
| AML 248 | chr11:94722188G>A | Remission | Minus 7 | 57 | 19410 | 0.294% | Minus 16 | 6 | 8100 | 0.074% |
| AML 248 | chr12:130171084A>G | Remission | Minus 7 | 0 | 23482 | 0.000% | Minus 16 | 2 | 9832 | 0.020% |
| AML 248 | chr13:104615517G>A | Remission | Minus 7 | 0 | 17521 | 0.000% | Minus 16 | 2 | 7274 | 0.027% |
| AML 248 | chr13:21126550C>T | Remission | Minus 7 | 1 | 24103 | 0.004% | Minus 16 | 0 | 10454 | 0.000% |
| AML 248 | chr13:48117971A>G | Remission | Minus 7 | 111 | 24808 | 0.447% | Minus 16 | 21 | 10039 | 0.209% |
| AML 248 | chr13:52962844A>G | Remission | Minus 7 | 1 | 18438 | 0.005% | Minus 16 | 3 | 8260 | 0.036% |
| AML 248 | chr13:62224453C>T | Remission | Minus 7 | 48 | 17767 | 0.270% | Minus 16 | 2 | 6443 | 0.031% |
| AML 248 | chr13:93287606G>A | Remission | Minus 7 | 44 | 15200 | 0.289% | Minus 16 | 3 | 6104 | 0.049% |
| AML 248 | chr14:39119285G>A | Remission | Minus 7 | 117 | 19849 | 0.589% | Minus 16 | 23 | 5652 | 0.407% |
| AML 248 | chr14:95340582G>A | Remission | Minus 7 | 3 | 25745 | 0.012% | Minus 16 | 0 | 12301 | 0.000% |
| AML 248 | chr15:37780604C>T | Remission | Minus 7 | 0 | 25758 | 0.000% | Minus 16 | 3 | 9040 | 0.033% |
| AML 248 | chr15:85938083C>T | Remission | Minus 7 | 57 | 21828 | 0.261% | Minus 16 | 12 | 9781 | 0.123% |
| AML 248 | chr16:51684669A>G | Remission | Minus 7 | 2 | 23467 | 0.009% | Minus 16 | 1 | 10494 | 0.010% |
| AML 248 | chr16:86772840G>C | Remission | Minus 7 | 0 | 22087 | 0.000% | Minus 16 | 3 | 11160 | 0.027% |
| AML 248 | chr16:9871241->A | Remission | Minus 7 | 41 | 16804 | 0.244% | Minus 16 | 3 | 10639 | 0.028% |
| AML 248 | chr17:5766396T>C | Remission | Minus 7 | 67 | 28461 | 0.235% | Minus 16 | 16 | 12970 | 0.123% |
| AML 248 | chr17:70021736C>A | Remission | Minus 7 | 3 | 19140 | 0.016% | Minus 16 | 0 | 7601 | 0.000% |
| AML 248 | chr19:19096041C>G | Remission | Minus 7 | 173 | 25863 | 0.669% | Minus 16 | 55 | 14163 | 0.388% |
| AML 248 | chr19:33701142C>T | Remission | Minus 7 | 58 | 27608 | 0.210% | Minus 16 | 18 | 13416 | 0.134% |
| AML 248 | chr2:158126080G>A | Remission | Minus 7 | 55 | 18469 | 0.298% | Minus 16 | 14 | 8446 | 0.166% |
| AML 248 | chr2:18197156G>A | Remission | Minus 7 | 292 | 16260 | 1.796% | Minus 16 | 92 | 6997 | 1.315% |
| AML 248 | chr2:220747088C>G | Remission | Minus 7 | 1 | 14557 | 0.007% | Minus 16 | 0 | 6258 | 0.000% |
| AML 248 | chr2:221271014A>T | Remission | Minus 7 | 647 | 17939 | 3.607% | Minus 16 | 298 | 7513 | 3.966% |
| AML 248 | chr2:239881851A>G | Remission | Minus 7 | 80 | 23354 | 0.343% | Minus 16 | 9 | 12381 | 0.073% |
| AML 248 | chr2:3974847C>T | Remission | Minus 7 | 49 | 22321 | 0.220% | Minus 16 | 12 | 8685 | 0.138% |
| AML 248 | chr2:49176542T>C | Remission | Minus 7 | 651 | 20531 | 3.171% | Minus 16 | 250 | 8999 | 2.778% |
| AML 248 | chr2:66358307C>T | Remission | Minus 7 | 43 | 17365 | 0.248% | Minus 16 | 4 | 5614 | 0.071% |
| AML 248 | chr2:72986641A>G | Remission | Minus 7 | 0 | 20148 | 0.000% | Minus 16 | 2 | 10114 | 0.020% |
| AML 248 | chr2:9414541C>T | Remission | Minus 7 | 66 | 23009 | 0.287% | Minus 16 | 12 | 9476 | 0.127% |
| AML 248 | chr20:61123120C>T | Remission | Minus 7 | 0 | 17243 | 0.000% | Minus 16 | 4 | 9297 | 0.043% |
| AML 248 | chr21:25343636C>T | Remission | Minus 7 | 57 | 20286 | 0.281% | Minus 16 | 10 | 8122 | 0.123% |
| AML 248 | chr21:37677112C>T | Remission | Minus 7 | 380 | 20929 | 1.816% | Minus 16 | 140 | 11775 | 1.189% |
| AML 248 | chr22:33909039C>T | Remission | Minus 7 | 46 | 22502 | 0.204% | Minus 16 | 6 | 9000 | 0.067% |
| AML 248 | chr22:47378266G>A | Remission | Minus 7 | 234 | 19948 | 1.173% | Minus 16 | 107 | 10578 | 1.012% |
| AML 248 | chr3:105395780G>C | Remission | Minus 7 | 44 | 16943 | 0.260% | Minus 16 | 3 | 6216 | 0.048% |
| AML 248 | chr3:138365619T>C | Remission | Minus 7 | 58 | 22033 | 0.263% | Minus 16 | 7 | 10789 | 0.065% |
| AML 248 | chr3:144253083A>G | Remission | Minus 7 | 0 | 19951 | 0.000% | Minus 16 | 2 | 7213 | 0.028% |
| AML 248 | chr3:15268647T>C | Remission | Minus 7 | 78 | 26165 | 0.298% | Minus 16 | 5 | 11570 | 0.043% |
| AML 248 | chr3:155069427G>T | Remission | Minus 7 | 463 | 21631 | 2.140% | Minus 16 | 147 | 7138 | 2.059% |
| AML 248 | chr3:193344044G>T | Remission | Minus 7 | 565 | 23678 | 2.386% | Minus 16 | 170 | 9229 | 1.842% |
| AML 248 | chr3:36719449C>T | Remission | Minus 7 | 49 | 19841 | 0.247% | Minus 16 | 3 | 7557 | 0.040% |
| AML 248 | chr4:114087248T>C | Remission | Minus 7 | 19 | 6433 | 0.295% | Minus 16 | 3 | 1480 | 0.203% |

| | | | | | | | | | | |
|---|---|---|---|---|---|---|---|---|---|---|
| AML 248 | chr5:117359851G>A | Remission | Minus 7 | 0 | 14669 | 0.000% | Minus 16 | 3 | 6320 | 0.047% |
| AML 248 | chr5:138420903C>T | Remission | Minus 7 | 71 | 23078 | 0.308% | Minus 16 | 11 | 10798 | 0.102% |
| AML 248 | chr5:37768102G>C | Remission | Minus 7 | 260 | 23785 | 1.093% | Minus 16 | 83 | 10151 | 0.818% |
| AML 248 | chr5:38936499G>- | Remission | Minus 7 | 652 | 21759 | 2.996% | Minus 16 | 199 | 8323 | 2.391% |
| AML 248 | chr5:43420421C>T | Remission | Minus 7 | 1 | 19488 | 0.005% | Minus 16 | 0 | 7257 | 0.000% |
| AML 248 | chr5:5942386C>T | Remission | Minus 7 | 46 | 20134 | 0.228% | Minus 16 | 9 | 8454 | 0.106% |
| AML 248 | chr5:64188253A>T | Remission | Minus 7 | 6 | 17917 | 0.033% | Minus 16 | 3 | 6319 | 0.047% |
| AML 248 | chr5:80012326G>A | Remission | Minus 7 | 2 | 19677 | 0.010% | Minus 16 | 2 | 8407 | 0.024% |
| AML 248 | chr6:105590513G>A | Remission | Minus 7 | 1 | 21903 | 0.005% | Minus 16 | 2 | 9062 | 0.022% |
| AML 248 | chr6:109374931A>G | Remission | Minus 7 | 1 | 17964 | 0.006% | Minus 16 | 2 | 7128 | 0.028% |
| AML 248 | chr6:78163810G>A | Remission | Minus 7 | 1 | 10684 | 0.009% | Minus 16 | 1 | 3425 | 0.029% |
| AML 248 | chr6:84798255C>A | Remission | Minus 7 | 90 | 27774 | 0.324% | Minus 16 | 10 | 8919 | 0.112% |
| AML 248 | chr6:89931274C>G | Remission | Minus 7 | 0 | 19148 | 0.000% | Minus 16 | 1 | 6878 | 0.015% |
| AML 248 | chr7:118695138C>T | Remission | Minus 7 | 235 | 17544 | 1.339% | Minus 16 | 66 | 6061 | 1.089% |
| AML 248 | chr7:132095315C>T | Remission | Minus 7 | 191 | 19196 | 0.995% | Minus 16 | 64 | 8618 | 0.743% |
| AML 248 | chr7:132300844G>A | Remission | Minus 7 | 47 | 20958 | 0.224% | Minus 16 | 8 | 12034 | 0.066% |
| AML 248 | chr7:158236471C>T | Remission | Minus 7 | 172 | 25683 | 0.670% | Minus 16 | 61 | 12242 | 0.498% |
| AML 248 | chr7:8418476G>A | Remission | Minus 7 | 56 | 19728 | 0.284% | Minus 16 | 4 | 5679 | 0.070% |
| AML 248 | chr8:139131792G>A | Remission | Minus 7 | 33 | 14715 | 0.224% | Minus 16 | 10 | 7507 | 0.133% |
| AML 248 | chr8:32518458T>C | Remission | Minus 7 | 525 | 16814 | 3.122% | Minus 16 | 201 | 6980 | 2.880% |
| AML 248 | chr8:3460641T>C | Remission | Minus 7 | 182 | 16080 | 1.132% | Minus 16 | 49 | 5840 | 0.839% |
| AML 248 | chr8:41719460G>A | Remission | Minus 7 | 78 | 23704 | 0.329% | Minus 16 | 14 | 13034 | 0.107% |
| AML 248 | chr8:60062081G>C | Remission | Minus 7 | 49 | 21128 | 0.232% | Minus 16 | 4 | 6733 | 0.059% |
| AML 248 | chr9:132157050C>T | Remission | Minus 7 | 256 | 21486 | 1.191% | Minus 16 | 95 | 10774 | 0.882% |
| AML 248 | chrX:17613357G>A | Remission | Minus 7 | 5 | 13537 | 0.037% | Minus 16 | 5 | 5547 | 0.090% |
| AML 250 | chr1:181789617G>A | Remission | Minus 7 | 1 | 10253 | 0.010% | Minus 92 | 0 | 9823 | 0.000% |
| AML 250 | chr1:214010845G>C | Remission | Minus 7 | 3 | 8744 | 0.034% | Minus 92 | 0 | 9968 | 0.000% |
| AML 250 | chr1:221501136T>C | Remission | Minus 7 | 5 | 7986 | 0.063% | Minus 92 | 11 | 6923 | 0.159% |
| AML 250 | chr1:237282606T>C | Remission | Minus 7 | 1 | 7323 | 0.014% | Minus 92 | 0 | 8436 | 0.000% |
| AML 250 | chr1:23924096A>G | Remission | Minus 7 | 3 | 6706 | 0.045% | Minus 92 | 1 | 4760 | 0.021% |
| AML 250 | chr10:89310721G>A | Remission | Minus 7 | 2 | 8706 | 0.023% | Minus 92 | 2 | 8739 | 0.023% |
| AML 250 | chr11:131996720C>T | Remission | Minus 7 | 0 | 10096 | 0.000% | Minus 92 | 2 | 10240 | 0.020% |
| AML 250 | chr12:129949400C>T | Remission | Minus 7 | 4 | 9367 | 0.043% | Minus 92 | 1 | 10226 | 0.010% |
| AML 250 | chr12:70325021T>G | Remission | Minus 7 | 2 | 6953 | 0.029% | Minus 92 | 0 | 8193 | 0.000% |
| AML 250 | chr12:77247619T>G | Remission | Minus 7 | 6 | 11998 | 0.050% | Minus 92 | 3 | 11847 | 0.025% |
| AML 250 | chr16:12095420T>C | Remission | Minus 7 | 1 | 4017 | 0.025% | Minus 92 | 6 | 8557 | 0.070% |
| AML 250 | chr16:48954956C>G | Remission | Minus 7 | 5 | 9286 | 0.054% | Minus 92 | 4 | 8788 | 0.046% |
| AML 250 | chr16:6012946G>T | Remission | Minus 7 | 0 | 6941 | 0.000% | Minus 92 | 1 | 6793 | 0.015% |
| AML 250 | chr17:530811C>T | Remission | Minus 7 | 1 | 8407 | 0.012% | Minus 92 | 4 | 9224 | 0.043% |
| AML 250 | chr17:61306084A>G | Remission | Minus 7 | 1 | 7758 | 0.013% | Minus 92 | 2 | 8801 | 0.023% |
| AML 250 | chr17:63631651T>- | Remission | Minus 7 | 0 | 5662 | 0.000% | Minus 92 | 1 | 2751 | 0.036% |
| AML 250 | chr17:65994202C>T | Remission | Minus 7 | 1 | 9878 | 0.010% | Minus 92 | 2 | 10546 | 0.019% |
| AML 250 | chr17:66238210G>A | Remission | Minus 7 | 4 | 10409 | 0.038% | Minus 92 | 3 | 9639 | 0.031% |
| AML 250 | chr17:79808688C>T | Remission | Minus 7 | 2 | 11044 | 0.018% | Minus 92 | 2 | 9053 | 0.022% |
| AML 250 | chr19:21590627G>A | Remission | Minus 7 | 3 | 8833 | 0.034% | Minus 92 | 1 | 9157 | 0.011% |
| AML 250 | chr2:174094079G>A | Remission | Minus 7 | 0 | 10479 | 0.000% | Minus 92 | 1 | 9449 | 0.011% |
| AML 250 | chr2:50581734G>A | Remission | Minus 7 | 0 | 8822 | 0.000% | Minus 92 | 2 | 8458 | 0.024% |
| AML 250 | chr2:7813343C>T | Remission | Minus 7 | 3 | 11492 | 0.026% | Minus 92 | 2 | 11236 | 0.018% |
| AML 250 | chr20:51346180C>- | Remission | Minus 7 | 2 | 11209 | 0.018% | Minus 92 | 3 | 10491 | 0.029% |
| AML 250 | chr20:58750265C>T | Remission | Minus 7 | 3 | 10300 | 0.029% | Minus 92 | 0 | 10600 | 0.000% |

| | | | | | | | | | | |
|---|---|---|---|---|---|---|---|---|---|---|
| AML 250 | chr3:74698594C>A | Remission | Minus 7 | 1 | 6542 | 0.015% | Minus 92 | 1 | 6817 | 0.015% |
| AML 250 | chr4:160661295T>C | Remission | Minus 7 | 0 | 7841 | 0.000% | Minus 92 | 1 | 7126 | 0.014% |
| AML 250 | chr4:76710373G>A | Remission | Minus 7 | 3 | 8842 | 0.034% | Minus 92 | 0 | 12242 | 0.000% |
| AML 250 | chr5:110274545C>T | Remission | Minus 7 | 1 | 9841 | 0.010% | Minus 92 | 0 | 8370 | 0.000% |
| AML 250 | chr5:120940492C>- | Remission | Minus 7 | 3 | 6462 | 0.046% | Minus 92 | 0 | 7966 | 0.000% |
| AML 250 | chr5:138657393C>G | Remission | Minus 7 | 4 | 11137 | 0.036% | Minus 92 | 2 | 10216 | 0.020% |
| AML 250 | chr5:174502164T>G | Remission | Minus 7 | 0 | 10336 | 0.000% | Minus 92 | 6 | 10902 | 0.055% |
| AML 250 | chr5:20054573T>C | Remission | Minus 7 | 5 | 9889 | 0.051% | Minus 92 | 0 | 8529 | 0.000% |
| AML 250 | chr5:93555256A>G | Remission | Minus 7 | 3 | 7616 | 0.039% | Minus 92 | 0 | 7243 | 0.000% |
| AML 250 | chr6:12234805G>A | Remission | Minus 7 | 4 | 9579 | 0.042% | Minus 92 | 1 | 9472 | 0.011% |
| AML 250 | chr6:72862557A>G | Remission | Minus 7 | 1 | 4768 | 0.021% | Minus 92 | 2 | 3830 | 0.052% |
| AML 250 | chr7:118263231A>G | Remission | Minus 7 | 0 | 7086 | 0.000% | Minus 92 | 5 | 8503 | 0.059% |
| AML 250 | chr7:128254555A>T | Remission | Minus 7 | 3 | 11599 | 0.026% | Minus 92 | 3 | 11992 | 0.025% |
| AML 250 | chr7:135229656A>G | Remission | Minus 7 | 3 | 8458 | 0.035% | Minus 92 | 2 | 10181 | 0.020% |
| AML 250 | chr7:86748341C>G | Remission | Minus 7 | 4 | 6740 | 0.059% | Minus 92 | 5 | 8331 | 0.060% |
| AML 250 | chr7:88486002C>T | Remission | Minus 7 | 1 | 6867 | 0.015% | Minus 92 | 0 | 8910 | 0.000% |
| AML 250 | chr8:10943046G>A | Remission | Minus 7 | 2 | 10307 | 0.019% | Minus 92 | 0 | 8279 | 0.000% |
| AML 250 | chr8:124042625C>T | Remission | Minus 7 | 0 | 8193 | 0.000% | Minus 92 | 1 | 7818 | 0.013% |
| AML 250 | chr8:30743661G>C | Remission | Minus 7 | 4 | 7899 | 0.051% | Minus 92 | 4 | 8986 | 0.045% |
| AML 250 | chr8:69333573C>G | Remission | Minus 7 | 2 | 8433 | 0.024% | Minus 92 | 2 | 10084 | 0.020% |
| AML 250 | chr9:24310680A>C | Remission | Minus 7 | 2 | 6236 | 0.032% | Minus 92 | 3 | 4618 | 0.065% |
| AML 250 | chrX:101261008C>T | Remission | Minus 7 | 1 | 9622 | 0.010% | Minus 92 | 0 | 11346 | 0.000% |
| AML 250 | chrX:103772364T>C | Remission | Minus 7 | 1 | 10061 | 0.010% | Minus 92 | 1 | 8986 | 0.011% |
| AML 250 | chrX:79618247C>T | Remission | Minus 7 | 2 | 9033 | 0.022% | Minus 92 | 0 | 10713 | 0.000% |
| AML 251 | chr1:113913185G>C | Remission | Minus 7 | 29 | 32235 | 0.090% | Minus 38 | 0 | 8407 | 0.000% |
| AML 251 | chr1:155904480T>A | Remission | Minus 7 | 30 | 31612 | 0.095% | Minus 38 | 4 | 6966 | 0.057% |
| AML 251 | chr1:179669423G>C | Remission | Minus 7 | 22 | 35783 | 0.061% | Minus 38 | 0 | 7675 | 0.000% |
| AML 251 | chr1:223462447G>C | Remission | Minus 7 | 11 | 26649 | 0.041% | Minus 38 | 1 | 6194 | 0.016% |
| AML 251 | chr1:75025278C>G | Remission | Minus 7 | 10 | 29112 | 0.034% | Minus 38 | 3 | 4331 | 0.069% |
| AML 251 | chr10:106665258G>C | Remission | Minus 7 | 24 | 38284 | 0.063% | Minus 38 | 0 | 5845 | 0.000% |
| AML 251 | chr10:36621510G>C | Remission | Minus 7 | 12 | 30767 | 0.039% | Minus 38 | 0 | 5306 | 0.000% |
| AML 251 | chr10:82944086->GGGAC | Remission | Minus 7 | 2 | 13749 | 0.015% | Minus 38 | 0 | 3030 | 0.000% |
| AML 251 | chr11:100254069A>T | Remission | Minus 7 | 9 | 14816 | 0.061% | Minus 38 | 0 | 3331 | 0.000% |
| AML 251 | chr11:102346774G>T | Remission | Minus 7 | 1 | 24935 | 0.004% | Minus 38 | 0 | 7475 | 0.000% |
| AML 251 | chr11:123643117C>G | Remission | Minus 7 | 30 | 27414 | 0.109% | Minus 38 | 0 | 8904 | 0.000% |
| AML 251 | chr11:5515469G>C | Remission | Minus 7 | 2 | 36336 | 0.006% | Minus 38 | 0 | 8059 | 0.000% |
| AML 251 | chr11:81220644G>A | Remission | Minus 7 | 16 | 32851 | 0.049% | Minus 38 | 0 | 6809 | 0.000% |
| AML 251 | chr11:99020392T>C | Remission | Minus 7 | 27 | 30712 | 0.088% | Minus 38 | 1 | 7024 | 0.014% |
| AML 251 | chr12:115262331G>- | Remission | Minus 7 | 26 | 42581 | 0.061% | Minus 38 | 3 | 7440 | 0.040% |
| AML 251 | chr12:127227392G>C | Remission | Minus 7 | 5 | 25313 | 0.020% | Minus 38 | 0 | 4476 | 0.000% |
| AML 251 | chr12:49052371T>C | Remission | Minus 7 | 21 | 33591 | 0.063% | Minus 38 | 0 | 8738 | 0.000% |
| AML 251 | chr12:65004477T>A | Remission | Minus 7 | 14 | 33764 | 0.041% | Minus 38 | 2 | 6966 | 0.029% |
| AML 251 | chr12:85898023T>C | Remission | Minus 7 | 16 | 27599 | 0.058% | Minus 38 | 2 | 6406 | 0.031% |
| AML 251 | chr12:93574960->GA | Remission | Minus 7 | 19 | 31791 | 0.060% | Minus 38 | 0 | 6245 | 0.000% |
| AML 251 | chr12:97751063G>A | Remission | Minus 7 | 18 | 27874 | 0.065% | Minus 38 | 0 | 6135 | 0.000% |
| AML 251 | chr13:65393072G>T | Remission | Minus 7 | 18 | 31644 | 0.057% | Minus 38 | 2 | 6278 | 0.032% |
| AML 251 | chr13:85906764G>T | Remission | Minus 7 | 23 | 33138 | 0.069% | Minus 38 | 2 | 5133 | 0.039% |
| AML 251 | chr13:96373712C>T | Remission | Minus 7 | 21 | 29569 | 0.071% | Minus 38 | 2 | 5358 | 0.037% |
| AML 251 | chr14:29615725G>C | Remission | Minus 7 | 13 | 29467 | 0.044% | Minus 38 | 0 | 5577 | 0.000% |
| AML 251 | chr15:101389694T>C | Remission | Minus 7 | 25 | 32344 | 0.077% | Minus 38 | 1 | 8421 | 0.012% |

| | | | | | | | | | | |
|---|---|---|---|---|---|---|---|---|---|---|
| AML 251 | chr15:86080219T>C | Remission | Minus 7 | 14 | 33922 | 0.041% | Minus 38 | 0 | 7081 | 0.000% |
| AML 251 | chr16:64315580C>G | Remission | Minus 7 | 3 | 26480 | 0.011% | Minus 38 | 0 | 6085 | 0.000% |
| AML 251 | chr17:30203376A>G | Remission | Minus 7 | 20 | 30256 | 0.066% | Minus 38 | 2 | 5944 | 0.034% |
| AML 251 | chr18:11378989G>C | Remission | Minus 7 | 1 | 36982 | 0.003% | Minus 38 | 0 | 8452 | 0.000% |
| AML 251 | chr18:54001774G>C | Remission | Minus 7 | 9 | 32277 | 0.028% | Minus 38 | 1 | 7181 | 0.014% |
| AML 251 | chr18:60227705C>T | Remission | Minus 7 | 27 | 32804 | 0.082% | Minus 38 | 1 | 5816 | 0.017% |
| AML 251 | chr18:70524437A>T | Remission | Minus 7 | 34 | 38943 | 0.087% | Minus 38 | 2 | 7061 | 0.028% |
| AML 251 | chr19:33623896C>A | Remission | Minus 7 | 21 | 33715 | 0.062% | Minus 38 | 2 | 8469 | 0.024% |
| AML 251 | chr19:57411010TTAAC>- | Remission | Minus 7 | 2 | 23875 | 0.008% | Minus 38 | 0 | 7813 | 0.000% |
| AML 251 | chr2:103041103C>A | Remission | Minus 7 | 28 | 37043 | 0.076% | Minus 38 | 1 | 6408 | 0.016% |
| AML 251 | chr2:173821810C>G | Remission | Minus 7 | 8 | 33042 | 0.024% | Minus 38 | 3 | 8571 | 0.035% |
| AML 251 | chr2:20903741C>G | Remission | Minus 7 | 11 | 26802 | 0.041% | Minus 38 | 0 | 6734 | 0.000% |
| AML 251 | chr2:212491338A>G | Remission | Minus 7 | 18 | 32485 | 0.055% | Minus 38 | 0 | 4044 | 0.000% |
| AML 251 | chr2:239529022C>T | Remission | Minus 7 | 23 | 34866 | 0.066% | Minus 38 | 0 | 8640 | 0.000% |
| AML 251 | chr2:30056856G>A | Remission | Minus 7 | 33 | 36314 | 0.091% | Minus 38 | 2 | 6380 | 0.031% |
| AML 251 | chr2:80125435T>A | Remission | Minus 7 | 2 | 32234 | 0.006% | Minus 38 | 0 | 7004 | 0.000% |
| AML 251 | chr20:40220782G>T | Remission | Minus 7 | 8 | 35882 | 0.022% | Minus 38 | 1 | 7823 | 0.013% |
| AML 251 | chr21:14885818T>A | Remission | Minus 7 | 5 | 33093 | 0.015% | Minus 38 | 0 | 6415 | 0.000% |
| AML 251 | chr21:46412475C>T | Remission | Minus 7 | 20 | 31713 | 0.063% | Minus 38 | 2 | 8224 | 0.024% |
| AML 251 | chr22:46163735->CCGGT | Remission | Minus 7 | 6 | 28723 | 0.021% | Minus 38 | 0 | 8350 | 0.000% |
| AML 251 | chr3:122121049G>A | Remission | Minus 7 | 21 | 36782 | 0.057% | Minus 38 | 2 | 6750 | 0.030% |
| AML 251 | chr3:13190265->GCGCA | Remission | Minus 7 | 8 | 34257 | 0.023% | Minus 38 | 0 | 9171 | 0.000% |
| AML 251 | chr3:181814957G>A | Remission | Minus 7 | 19 | 32665 | 0.058% | Minus 38 | 2 | 6778 | 0.030% |
| AML 251 | chr3:193785856G>C | Remission | Minus 7 | 20 | 32009 | 0.062% | Minus 38 | 4 | 8754 | 0.046% |
| AML 251 | chr3:2451012G>C | Remission | Minus 7 | 3 | 33822 | 0.009% | Minus 38 | 0 | 5979 | 0.000% |
| AML 251 | chr3:48782120T>G | Remission | Minus 7 | 31 | 33733 | 0.092% | Minus 38 | 0 | 6266 | 0.000% |
| AML 251 | chr3:85974475A>T | Remission | Minus 7 | 4 | 37948 | 0.011% | Minus 38 | 0 | 5906 | 0.000% |
| AML 251 | chr4:108258111->AAG | Remission | Minus 7 | 1 | 11448 | 0.009% | Minus 38 | 0 | 2533 | 0.000% |
| AML 251 | chr4:4944193G>T | Remission | Minus 7 | 14 | 14555 | 0.096% | Minus 38 | 2 | 4876 | 0.041% |
| AML 251 | chr5:26460730A>T | Remission | Minus 7 | 1 | 31976 | 0.003% | Minus 38 | 0 | 6585 | 0.000% |
| AML 251 | chr5:52223002T>C | Remission | Minus 7 | 14 | 27382 | 0.051% | Minus 38 | 0 | 3465 | 0.000% |
| AML 251 | chr5:9783059G>A | Remission | Minus 7 | 24 | 31926 | 0.075% | Minus 38 | 1 | 7743 | 0.013% |
| AML 251 | chr6:107663343G>A | Remission | Minus 7 | 30 | 40115 | 0.075% | Minus 38 | 0 | 7713 | 0.000% |
| AML 251 | chr6:163301683T>C | Remission | Minus 7 | 38 | 41228 | 0.092% | Minus 38 | 0 | 7642 | 0.000% |
| AML 251 | chr6:163336041->GGGGCA | Remission | Minus 7 | 13 | 29668 | 0.044% | Minus 38 | 3 | 8442 | 0.036% |
| AML 251 | chr6:68956057G>C | Remission | Minus 7 | 17 | 34527 | 0.049% | Minus 38 | 0 | 8356 | 0.000% |
| AML 251 | chr7:100561558C>G | Remission | Minus 7 | 28 | 39753 | 0.070% | Minus 38 | 2 | 8596 | 0.023% |
| AML 251 | chr7:117041084G>C | Remission | Minus 7 | 21 | 26901 | 0.078% | Minus 38 | 1 | 4737 | 0.021% |
| AML 251 | chr7:143951856G>A | Remission | Minus 7 | 19 | 35260 | 0.054% | Minus 38 | 0 | 8536 | 0.000% |
| AML 251 | chr7:147509396T>C | Remission | Minus 7 | 15 | 32057 | 0.047% | Minus 38 | 1 | 5929 | 0.017% |
| AML 251 | chr7:47605953->TGCCC | Remission | Minus 7 | 10 | 34779 | 0.029% | Minus 38 | 0 | 7938 | 0.000% |
| AML 251 | chr8:102015064T>C | Remission | Minus 7 | 31 | 39753 | 0.078% | Minus 38 | 0 | 6695 | 0.000% |
| AML 251 | chr8:14238270C>A | Remission | Minus 7 | 23 | 30409 | 0.076% | Minus 38 | 0 | 5047 | 0.000% |
| AML 251 | chr8:32722302->TGA | Remission | Minus 7 | 12 | 29979 | 0.040% | Minus 38 | 0 | 4753 | 0.000% |
| AML 251 | chr9:137746715C>T | Remission | Minus 7 | 7 | 30507 | 0.023% | Minus 38 | 1 | 5735 | 0.017% |
| AML 251 | chr9:5231567T>A | Remission | Minus 7 | 33 | 34802 | 0.095% | Minus 38 | 0 | 9692 | 0.000% |
| AML 251 | chrX:152907200G>T | Remission | Minus 7 | 2 | 37336 | 0.005% | Minus 38 | 0 | 10321 | 0.000% |
| AML 251 | chrX:29072621->CCCC | Remission | Minus 7 | 9 | 34054 | 0.026% | Minus 38 | 0 | 5933 | 0.000% |
| AML 251 | chrX:6231428T>A | Remission | Minus 7 | 23 | 34753 | 0.066% | Minus 38 | 1 | 7862 | 0.013% |
| AML 121 | chr10:63867282G>A | 3 months post-transplant | 87 | 1 | 11534 | 0.009% | 87 | 0 | 6796 | 0.000% |

| | | | | | | | | | | |
|---|---|---|---|---|---|---|---|---|---|---|
| AML 121 | chr13:113254106G>A | 3 months post-transplant | 87 | 1 | 12055 | 0.008% | 87 | 0 | 6822 | 0.000% |
| AML 121 | chr2:104729661C>G | 3 months post-transplant | 87 | 2 | 15999 | 0.013% | 87 | 0 | 7920 | 0.000% |
| AML 121 | chr9:68940919G>C | 3 months post-transplant | 87 | 0 | 10311 | 0.000% | 87 | 1 | 6748 | 0.015% |
| AML 122 | chr10:129942497C>G | 3 months post-transplant | 89 | 1 | 15251 | 0.007% | 89 | 0 | 5088 | 0.000% |
| AML 122 | chr11:28544552G>C | 3 months post-transplant | 89 | 7 | 6902 | 0.101% | 89 | 4 | 3959 | 0.101% |
| AML 122 | chr11:57934563A>G | 3 months post-transplant | 89 | 10 | 10996 | 0.091% | 89 | 1 | 2547 | 0.039% |
| AML 122 | chr12:124676438T>G | 3 months post-transplant | 89 | 3 | 11615 | 0.026% | 89 | 4 | 4907 | 0.082% |
| AML 122 | chr16:2598618C>A | 3 months post-transplant | 89 | 18 | 20012 | 0.090% | 89 | 5 | 5186 | 0.096% |
| AML 122 | chr18:49843886G>- | 3 months post-transplant | 89 | 1 | 3448 | 0.029% | 89 | 0 | 4080 | 0.000% |
| AML 122 | chr2:141283092T>C | 3 months post-transplant | 89 | 1 | 12448 | 0.008% | 89 | 0 | 3009 | 0.000% |
| AML 122 | chr2:25239199A>G | 3 months post-transplant | 89 | 8 | 13482 | 0.059% | 89 | 2 | 3585 | 0.056% |
| AML 122 | chr4:138104421A>C | 3 months post-transplant | 89 | 6 | 11278 | 0.053% | 89 | 3 | 4927 | 0.061% |
| AML 122 | chr7:122044719T>A | 3 months post-transplant | 89 | 14 | 22241 | 0.063% | 89 | 2 | 4617 | 0.043% |
| AML 123 | chr2:236010356G>C | 3 months post-transplant | 84 | 1 | 8073 | 0.012% | 84 | 0 | 8875 | 0.000% |
| AML 123 | chr7:148842182T>C | 3 months post-transplant | 84 | 1 | 167 | 0.599% | 84 | 0 | 8720 | 0.000% |
| AML 123 | chrX:134750569C>G | 3 months post-transplant | 84 | 1 | 4921 | 0.020% | 84 | 0 | 4304 | 0.000% |
| AML 129 | chr11:133840616A>G | 3 months post-transplant | 90 | 1 | 14660 | 0.007% | 90 | 0 | 9937 | 0.000% |
| AML 129 | chr15:23860086C>G | 3 months post-transplant | 90 | 1 | 14655 | 0.007% | 90 | 0 | 11343 | 0.000% |
| AML 129 | chr2:142221145A>G | 3 months post-transplant | 90 | 1 | 15810 | 0.006% | 90 | 0 | 11785 | 0.000% |
| AML 129 | chr20:7940367A>G | 3 months post-transplant | 90 | 1 | 8628 | 0.012% | 90 | 0 | 10589 | 0.000% |
| AML 129 | chr9:103359925T>C | 3 months post-transplant | 90 | 1 | 14038 | 0.007% | 90 | 0 | 6720 | 0.000% |
| AML 130 | chr1:106285578T>G | 3 months post-transplant | 99 | 1 | 13841 | 0.007% | 89 | 0 | 9873 | 0.000% |
| AML 130 | chr10:34621587A>T | 3 months post-transplant | 99 | 7 | 14921 | 0.047% | 89 | 6 | 9455 | 0.063% |
| AML 130 | chr10:85852159T>C | 3 months post-transplant | 99 | 1 | 10585 | 0.009% | 89 | 0 | 11765 | 0.000% |
| AML 130 | chr11:122692233A>G | 3 months post-transplant | 99 | 4 | 15395 | 0.026% | 89 | 2 | 10447 | 0.019% |
| AML 130 | chr11:38802150A>G | 3 months post-transplant | 99 | 1 | 15179 | 0.007% | 89 | 0 | 10315 | 0.000% |
| AML 130 | chr12:104741654A>G | 3 months post-transplant | 99 | 1 | 16767 | 0.006% | 89 | 1 | 10985 | 0.009% |
| AML 130 | chr12:13896169A>G | 3 months post-transplant | 99 | 0 | 8720 | 0.000% | 89 | 1 | 10009 | 0.010% |
| AML 130 | chr14:26683707T>C | 3 months post-transplant | 99 | 3 | 7273 | 0.041% | 89 | 3 | 10226 | 0.029% |
| AML 130 | chr14:48973412A>G | 3 months post-transplant | 99 | 1 | 14850 | 0.007% | 89 | 0 | 11400 | 0.000% |
| AML 130 | chr14:90423441C>T | 3 months post-transplant | 99 | 23 | 15336 | 0.150% | 89 | 10 | 11317 | 0.088% |
| AML 130 | chr14:98255192T>C | 3 months post-transplant | 99 | 31 | 19592 | 0.158% | 89 | 11 | 11681 | 0.094% |
| AML 130 | chr16:46743605A>G | 3 months post-transplant | 99 | 3 | 5701 | 0.053% | 89 | 6 | 11439 | 0.052% |
| AML 130 | chr16:65528696G>C | 3 months post-transplant | 99 | 14 | 14634 | 0.096% | 89 | 9 | 10289 | 0.087% |
| AML 130 | chr2:119166886A>C | 3 months post-transplant | 99 | 1 | 11954 | 0.008% | 89 | 0 | 10450 | 0.000% |
| AML 130 | chr2:202559796G>C | 3 months post-transplant | 99 | 1 | 13997 | 0.007% | 89 | 0 | 7475 | 0.000% |
| AML 130 | chr20:11727205C>G | 3 months post-transplant | 99 | 2 | 13017 | 0.015% | 89 | 0 | 8953 | 0.000% |
| AML 130 | chr3:128411745G>C | 3 months post-transplant | 99 | 1 | 14549 | 0.007% | 89 | 0 | 9425 | 0.000% |
| AML 130 | chr8:68116084T>C | 3 months post-transplant | 99 | 2 | 19360 | 0.010% | 89 | 0 | 11859 | 0.000% |
| AML 130 | chr9:78680128G>A | 3 months post-transplant | 99 | 1 | 18388 | 0.005% | 89 | 0 | 9717 | 0.000% |
| AML 131 | chr2:122916745A>G | 3 months post-transplant | 86 | 0 | 5423 | 0.000% | 98 | 1 | 12304 | 0.008% |
| AML 235 | chr1:165567242T>G | 3 months post-transplant | 84 | 1 | 3136 | 0.032% | 97 | 0 | 265 | 0.000% |
| AML 235 | chr1:19857444C>G | 3 months post-transplant | 84 | 4 | 18955 | 0.021% | 97 | 0 | 7285 | 0.000% |
| AML 235 | chr1:23429400T>C | 3 months post-transplant | 84 | 1 | 19771 | 0.005% | 97 | 0 | 8192 | 0.000% |
| AML 235 | chr1:25107284G>A | 3 months post-transplant | 84 | 3 | 20580 | 0.015% | 97 | 0 | 8798 | 0.000% |
| AML 235 | chr1:87885697C>T | 3 months post-transplant | 84 | 1 | 19290 | 0.005% | 97 | 0 | 8006 | 0.000% |
| AML 235 | chr10:100081931A>G | 3 months post-transplant | 84 | 4 | 23014 | 0.017% | 97 | 0 | 7231 | 0.000% |
| AML 235 | chr10:125533657C>T | 3 months post-transplant | 84 | 1 | 16837 | 0.006% | 97 | 1 | 8066 | 0.012% |
| AML 235 | chr10:17952177G>C | 3 months post-transplant | 84 | 2 | 21195 | 0.009% | 97 | 1 | 8138 | 0.012% |
| AML 235 | chr11:118519716>GG | 3 months post-transplant | 84 | 2 | 22873 | 0.009% | 97 | 0 | 9248 | 0.000% |

| | | | | | | | | | | |
|---|---|---|---|---|---|---|---|---|---|---|
| AML 235 | chr11:123940800G>C | 3 months post-transplant | 84 | 2 | 16922 | 0.012% | 97 | 1 | 7916 | 0.013% |
| AML 235 | chr11:31249680T>C | 3 months post-transplant | 84 | 2 | 21673 | 0.009% | 97 | 0 | 6895 | 0.000% |
| AML 235 | chr12:221476C>T | 3 months post-transplant | 84 | 1 | 19926 | 0.005% | 97 | 0 | 8979 | 0.000% |
| AML 235 | chr12:25623624T>C | 3 months post-transplant | 84 | 1 | 13173 | 0.008% | 97 | 0 | 2646 | 0.000% |
| AML 235 | chr12:3529546T>C | 3 months post-transplant | 84 | 6 | 18106 | 0.033% | 97 | 0 | 6513 | 0.000% |
| AML 235 | chr12:5471503G>A | 3 months post-transplant | 84 | 1 | 16439 | 0.006% | 97 | 0 | 7235 | 0.000% |
| AML 235 | chr12:64733476T>C | 3 months post-transplant | 84 | 0 | 19483 | 0.000% | 97 | 1 | 7103 | 0.014% |
| AML 235 | chr12:96117801G>A | 3 months post-transplant | 84 | 1 | 22156 | 0.005% | 97 | 0 | 8257 | 0.000% |
| AML 235 | chr14:39769140G>C | 3 months post-transplant | 84 | 2 | 18129 | 0.011% | 97 | 0 | 6036 | 0.000% |
| AML 235 | chr14:78099338C>T | 3 months post-transplant | 84 | 1 | 15309 | 0.007% | 97 | 0 | 6780 | 0.000% |
| AML 235 | chr14:81676314G>A | 3 months post-transplant | 84 | 2 | 17255 | 0.012% | 97 | 1 | 7200 | 0.014% |
| AML 235 | chr15:31787892A>G | 3 months post-transplant | 84 | 1 | 9747 | 0.010% | 97 | 0 | 2606 | 0.000% |
| AML 235 | chr15:66677904G>A | 3 months post-transplant | 84 | 3 | 24798 | 0.012% | 97 | 0 | 8769 | 0.000% |
| AML 235 | chr15:69028002C>T | 3 months post-transplant | 84 | 4 | 23852 | 0.017% | 97 | 0 | 8735 | 0.000% |
| AML 235 | chr16:85603369>CC | 3 months post-transplant | 84 | 1 | 13648 | 0.007% | 97 | 0 | 5493 | 0.000% |
| AML 235 | chr16:87008603A>G | 3 months post-transplant | 84 | 1 | 18372 | 0.005% | 97 | 0 | 7999 | 0.000% |
| AML 235 | chr17:52250848A> | 3 months post-transplant | 84 | 1 | 17519 | 0.006% | 97 | 0 | 5959 | 0.000% |
| AML 235 | chr18:24561167G>A | 3 months post-transplant | 84 | 2 | 23054 | 0.009% | 97 | 0 | 8178 | 0.000% |
| AML 235 | chr18:40037787G>T | 3 months post-transplant | 84 | 3 | 17864 | 0.017% | 97 | 0 | 7966 | 0.000% |
| AML 235 | chr19:50383921C>G | 3 months post-transplant | 84 | 1 | 12834 | 0.008% | 97 | 0 | 8559 | 0.000% |
| AML 235 | chr2:7857516C>T | 3 months post-transplant | 84 | 2 | 24878 | 0.008% | 97 | 0 | 9208 | 0.000% |
| AML 235 | chr21:40416188G>A | 3 months post-transplant | 84 | 4 | 18896 | 0.021% | 97 | 1 | 9285 | 0.011% |
| AML 235 | chr22:25780119G>A | 3 months post-transplant | 84 | 2 | 16464 | 0.012% | 97 | 0 | 4735 | 0.000% |
| AML 235 | chr22:29698198A>G | 3 months post-transplant | 84 | 3 | 19397 | 0.015% | 97 | 0 | 7999 | 0.000% |
| AML 235 | chr22:29837510T>C | 3 months post-transplant | 84 | 3 | 16773 | 0.018% | 97 | 0 | 7568 | 0.000% |
| AML 235 | chr3:107163518T>C | 3 months post-transplant | 84 | 1 | 15717 | 0.006% | 97 | 0 | 5692 | 0.000% |
| AML 235 | chr3:175778314A>G | 3 months post-transplant | 84 | 2 | 19439 | 0.010% | 97 | 0 | 7347 | 0.000% |
| AML 235 | chr3:181066160T>C | 3 months post-transplant | 84 | 3 | 18649 | 0.016% | 97 | 0 | 7156 | 0.000% |
| AML 235 | chr3:50812223A>G | 3 months post-transplant | 84 | 5 | 17985 | 0.028% | 97 | 0 | 8316 | 0.000% |
| AML 235 | chr3:54031649G>A | 3 months post-transplant | 84 | 1 | 16456 | 0.006% | 97 | 0 | 7374 | 0.000% |
| AML 235 | chr5:3126570T>C | 3 months post-transplant | 84 | 2 | 17827 | 0.011% | 97 | 0 | 7706 | 0.000% |
| AML 235 | chr5:4306175C>T | 3 months post-transplant | 84 | 2 | 20906 | 0.010% | 97 | 0 | 8503 | 0.000% |
| AML 235 | chr5:45510620C>T | 3 months post-transplant | 84 | 1 | 17326 | 0.006% | 97 | 0 | 7411 | 0.000% |
| AML 235 | chr6:105955681A>G | 3 months post-transplant | 84 | 2 | 15549 | 0.013% | 97 | 0 | 7067 | 0.000% |
| AML 235 | chr6:126010195G>A | 3 months post-transplant | 84 | 2 | 17697 | 0.011% | 97 | 0 | 7042 | 0.000% |
| AML 235 | chr6:138865853C>G | 3 months post-transplant | 84 | 1 | 13057 | 0.008% | 97 | 0 | 6081 | 0.000% |
| AML 235 | chr6:167547928C>A | 3 months post-transplant | 84 | 3 | 21690 | 0.014% | 97 | 0 | 8111 | 0.000% |
| AML 235 | chr6:7186967T>A | 3 months post-transplant | 84 | 1 | 19420 | 0.005% | 97 | 0 | 7853 | 0.000% |
| AML 235 | chr6:85492862C>T | 3 months post-transplant | 84 | 2 | 20527 | 0.010% | 97 | 0 | 7131 | 0.000% |
| AML 235 | chr6:94066528A>T | 3 months post-transplant | 84 | 3 | 17053 | 0.018% | 97 | 0 | 7910 | 0.000% |
| AML 235 | chr7:24086625G>A | 3 months post-transplant | 84 | 1 | 13730 | 0.007% | 97 | 0 | 7391 | 0.000% |
| AML 235 | chr7:33743414C>G | 3 months post-transplant | 84 | 2 | 18095 | 0.011% | 97 | 1 | 7295 | 0.014% |
| AML 235 | chr7:3972928G>C | 3 months post-transplant | 84 | 6 | 20347 | 0.029% | 97 | 0 | 9158 | 0.000% |
| AML 235 | chr8:129629953T>G | 3 months post-transplant | 84 | 35 | 22485 | 0.156% | 97 | 0 | 8284 | 0.000% |
| AML 235 | chr8:20884829T>C | 3 months post-transplant | 84 | 2 | 17583 | 0.011% | 97 | 0 | 8081 | 0.000% |
| AML 235 | chr8:2862203C>A | 3 months post-transplant | 84 | 1 | 17416 | 0.006% | 97 | 0 | 7435 | 0.000% |
| AML 235 | chr8:73451557T>C | 3 months post-transplant | 84 | 1 | 17563 | 0.006% | 97 | 0 | 8612 | 0.000% |
| AML 235 | chr8:74066786G>A | 3 months post-transplant | 84 | 1 | 14380 | 0.007% | 97 | 0 | 7597 | 0.000% |
| AML 235 | chr9:109084857C>T | 3 months post-transplant | 84 | 2 | 18935 | 0.011% | 97 | 0 | 5980 | 0.000% |
| AML 235 | chr9:135143068T>C | 3 months post-transplant | 84 | 3 | 23615 | 0.013% | 97 | 0 | 7862 | 0.000% |

| | | | | | | | | | | |
|---|---|---|---|---|---|---|---|---|---|---|
| AML 235 | chr9:23002947C>T | 3 months post-transplant | 84 | 4 | 17679 | 0.023% | 97 | 0 | 6164 | 0.000% |
| AML 235 | chrX:107140810C>G | 3 months post-transplant | 84 | 2 | 16758 | 0.012% | 97 | 0 | 6922 | 0.000% |
| AML 235 | chrX:148725203C>T | 3 months post-transplant | 84 | 1 | 20273 | 0.005% | 97 | 1 | 8546 | 0.012% |
| AML 239 | chr11:112858841G>A | 3 months post-transplant | 83 | 0 | 4769 | 0.000% | 97 | 1 | 11743 | 0.009% |
| AML 239 | chr6:70101478G>C | 3 months post-transplant | 83 | 1 | 2966 | 0.034% | 97 | 1 | 9872 | 0.010% |
| AML 239 | chr8:5679049T>A | 3 months post-transplant | 83 | 0 | 2875 | 0.000% | 97 | 1 | 10475 | 0.010% |
| AML 239 | chr9:9417477C>T | 3 months post-transplant | 83 | 1 | 3973 | 0.025% | 97 | 0 | 10985 | 0.000% |
| AML 241 | chr13:42762113T>G | 3 months post-transplant | 92 | 1 | 18905 | 0.005% | 92 | 0 | 8553 | 0.000% |
| AML 241 | chr8:137985582C>T | 3 months post-transplant | 92 | 1 | 18342 | 0.005% | 92 | 0 | 9064 | 0.000% |
| AML 245 | chr11:32396398G> | 3 months post-transplant | 92 | 1 | 4818 | 0.021% | 98 | 0 | 14904 | 0.000% |
| AML 245 | chr11:7763661G>A | 3 months post-transplant | 92 | 1 | 4084 | 0.024% | 98 | 0 | 7661 | 0.000% |
| AML 245 | chr14:95613620G>A | 3 months post-transplant | 92 | 0 | 4086 | 0.000% | 98 | 1 | 12954 | 0.008% |
| AML 245 | chr20:20833493G>A | 3 months post-transplant | 92 | 1 | 5538 | 0.018% | 98 | 0 | 17359 | 0.000% |
| AML 245 | chr4:119684667T>C | 3 months post-transplant | 92 | 0 | 3551 | 0.000% | 98 | 1 | 8971 | 0.011% |
| AML 245 | chr5:140005751T>C | 3 months post-transplant | 92 | 1 | 4795 | 0.021% | 98 | 0 | 15289 | 0.000% |
| AML 245 | chr7:156002360C>T | 3 months post-transplant | 92 | 0 | 4798 | 0.000% | 98 | 1 | 13550 | 0.007% |
| AML 248 | chr1:14609249G>A | 3 months post-transplant | 86 | 14 | 13551 | 0.103% | 93 | 0 | 9171 | 0.000% |
| AML 248 | chr1:147656369C>T | 3 months post-transplant | 86 | 29 | 13972 | 0.208% | 93 | 0 | 6938 | 0.000% |
| AML 248 | chr1:203737152A>G | 3 months post-transplant | 86 | 12 | 8374 | 0.143% | 93 | 0 | 4345 | 0.000% |
| AML 248 | chr1:2820056G>A | 3 months post-transplant | 86 | 25 | 15844 | 0.158% | 93 | 0 | 6668 | 0.000% |
| AML 248 | chr1:40866191T>C | 3 months post-transplant | 86 | 44 | 16085 | 0.274% | 93 | 0 | 8049 | 0.000% |
| AML 248 | chr1:7466894C>T | 3 months post-transplant | 86 | 21 | 17285 | 0.121% | 93 | 0 | 7897 | 0.000% |
| AML 248 | chr10:118507409T>C | 3 months post-transplant | 86 | 14 | 13005 | 0.108% | 93 | 0 | 7468 | 0.000% |
| AML 248 | chr10:16313939T>A | 3 months post-transplant | 86 | 16 | 11619 | 0.138% | 93 | 0 | 8859 | 0.000% |
| AML 248 | chr10:17429036>A | 3 months post-transplant | 86 | 2 | 8602 | 0.023% | 93 | 0 | 8543 | 0.000% |
| AML 248 | chr11:11641040G>A | 3 months post-transplant | 86 | 22 | 15570 | 0.141% | 93 | 0 | 9108 | 0.000% |
| AML 248 | hr11:134654301>GGGTGCAT | 3 months post-transplant | 86 | 16 | 13596 | 0.118% | 93 | 0 | 8612 | 0.000% |
| AML 248 | chr11:36861290G>A | 3 months post-transplant | 86 | 23 | 10562 | 0.218% | 93 | 0 | 7833 | 0.000% |
| AML 248 | chr11:94722188G>A | 3 months post-transplant | 86 | 16 | 12057 | 0.133% | 93 | 0 | 7914 | 0.000% |
| AML 248 | chr13:48117971A>G | 3 months post-transplant | 86 | 31 | 15740 | 0.197% | 93 | 0 | 8454 | 0.000% |
| AML 248 | chr13:62224453C>T | 3 months post-transplant | 86 | 15 | 11148 | 0.135% | 93 | 0 | 8501 | 0.000% |
| AML 248 | chr13:93287606G>A | 3 months post-transplant | 86 | 16 | 9217 | 0.174% | 93 | 0 | 7852 | 0.000% |
| AML 248 | chr14:39119285G>A | 3 months post-transplant | 86 | 19 | 12296 | 0.155% | 93 | 0 | 7709 | 0.000% |
| AML 248 | chr15:37780604C>T | 3 months post-transplant | 86 | 1 | 15622 | 0.006% | 93 | 0 | 8598 | 0.000% |
| AML 248 | chr15:85938083C>T | 3 months post-transplant | 86 | 25 | 14073 | 0.178% | 93 | 0 | 8675 | 0.000% |
| AML 248 | chr16:9871241>A | 3 months post-transplant | 86 | 16 | 10128 | 0.158% | 93 | 0 | 8275 | 0.000% |
| AML 248 | chr17:5766396T>C | 3 months post-transplant | 86 | 38 | 19084 | 0.199% | 93 | 0 | 9599 | 0.000% |
| AML 248 | chr19:19096041C>G | 3 months post-transplant | 86 | 21 | 17276 | 0.122% | 93 | 0 | 9013 | 0.000% |
| AML 248 | chr19:33701142C>T | 3 months post-transplant | 86 | 22 | 18540 | 0.119% | 93 | 0 | 8866 | 0.000% |
| AML 248 | chr2:158126080G>A | 3 months post-transplant | 86 | 16 | 12134 | 0.132% | 93 | 0 | 8069 | 0.000% |
| AML 248 | chr2:18197156G>A | 3 months post-transplant | 86 | 24 | 9890 | 0.243% | 93 | 0 | 6906 | 0.000% |
| AML 248 | chr2:221271014A>T | 3 months post-transplant | 86 | 10 | 11033 | 0.091% | 93 | 1 | 8708 | 0.011% |
| AML 248 | chr2:239881851A>G | 3 months post-transplant | 86 | 24 | 15443 | 0.155% | 93 | 0 | 8978 | 0.000% |
| AML 248 | chr2:3974847C>T | 3 months post-transplant | 86 | 21 | 14270 | 0.147% | 93 | 0 | 7797 | 0.000% |
| AML 248 | chr2:49176542T>C | 3 months post-transplant | 86 | 18 | 13116 | 0.137% | 93 | 0 | 8242 | 0.000% |
| AML 248 | chr2:66358307C>T | 3 months post-transplant | 86 | 21 | 10289 | 0.204% | 93 | 0 | 6181 | 0.000% |
| AML 248 | chr2:9414541C>T | 3 months post-transplant | 86 | 20 | 14236 | 0.140% | 93 | 0 | 8195 | 0.000% |
| AML 248 | chr21:25343636C>T | 3 months post-transplant | 86 | 21 | 12399 | 0.169% | 93 | 0 | 8442 | 0.000% |
| AML 248 | chr21:37677112C>T | 3 months post-transplant | 86 | 19 | 14567 | 0.130% | 93 | 0 | 8261 | 0.000% |
| AML 248 | chr22:33909039C>T | 3 months post-transplant | 86 | 23 | 13901 | 0.165% | 93 | 0 | 7693 | 0.000% |

| | | | | | | | | | | |
|---|---|---|---|---|---|---|---|---|---|---|
| AML 248 | chr22:47378266G>A | 3 months post-transplant | 86 | 17 | 13219 | 0.129% | 93 | 0 | 7197 | 0.000% |
| AML 248 | chr3:105395780G>C | 3 months post-transplant | 86 | 27 | 10508 | 0.257% | 93 | 0 | 7937 | 0.000% |
| AML 248 | chr3:138365619T>C | 3 months post-transplant | 86 | 24 | 13650 | 0.176% | 93 | 0 | 8357 | 0.000% |
| AML 248 | chr3:15268647T>C | 3 months post-transplant | 86 | 37 | 16890 | 0.219% | 93 | 0 | 8741 | 0.000% |
| AML 248 | chr3:155069427G>T | 3 months post-transplant | 86 | 31 | 13335 | 0.232% | 93 | 0 | 8622 | 0.000% |
| AML 248 | chr3:193344044G>T | 3 months post-transplant | 86 | 21 | 15133 | 0.139% | 93 | 0 | 8790 | 0.000% |
| AML 248 | chr3:36719449C>T | 3 months post-transplant | 86 | 23 | 12948 | 0.178% | 93 | 0 | 8235 | 0.000% |
| AML 248 | chr4:114087248T>C | 3 months post-transplant | 86 | 7 | 3246 | 0.216% | 93 | 0 | 2695 | 0.000% |
| AML 248 | chr5:138420903C>T | 3 months post-transplant | 86 | 33 | 15156 | 0.218% | 93 | 0 | 8175 | 0.000% |
| AML 248 | chr5:37768102G>C | 3 months post-transplant | 86 | 20 | 15150 | 0.132% | 93 | 0 | 8904 | 0.000% |
| AML 248 | chr5:38936499G> | 3 months post-transplant | 86 | 22 | 13480 | 0.163% | 93 | 0 | 9087 | 0.000% |
| AML 248 | chr5:5942386C>T | 3 months post-transplant | 86 | 19 | 12686 | 0.150% | 93 | 0 | 9028 | 0.000% |
| AML 248 | chr6:109374931A>G | 3 months post-transplant | 86 | 2 | 9685 | 0.021% | 93 | 0 | 6685 | 0.000% |
| AML 248 | chr6:84798255C>A | 3 months post-transplant | 86 | 30 | 17876 | 0.168% | 93 | 0 | 9091 | 0.000% |
| AML 248 | chr7:118695138C>T | 3 months post-transplant | 86 | 25 | 10995 | 0.227% | 93 | 0 | 7033 | 0.000% |
| AML 248 | chr7:132095315C>T | 3 months post-transplant | 86 | 15 | 12444 | 0.121% | 93 | 0 | 6710 | 0.000% |
| AML 248 | chr7:132300844G>A | 3 months post-transplant | 86 | 22 | 14325 | 0.154% | 93 | 0 | 8123 | 0.000% |
| AML 248 | chr7:158236471C>T | 3 months post-transplant | 86 | 33 | 17353 | 0.190% | 93 | 0 | 7305 | 0.000% |
| AML 248 | chr7:8418476G>A | 3 months post-transplant | 86 | 40 | 10804 | 0.370% | 93 | 0 | 7294 | 0.000% |
| AML 248 | chr8:139131792G>A | 3 months post-transplant | 86 | 14 | 9241 | 0.151% | 93 | 0 | 8022 | 0.000% |
| AML 248 | chr8:32518458T>C | 3 months post-transplant | 86 | 16 | 10759 | 0.149% | 93 | 0 | 7591 | 0.000% |
| AML 248 | chr8:3460641T>C | 3 months post-transplant | 86 | 15 | 9912 | 0.151% | 93 | 0 | 6154 | 0.000% |
| AML 248 | chr8:41719460G>A | 3 months post-transplant | 86 | 27 | 16085 | 0.168% | 93 | 0 | 7068 | 0.000% |
| AML 248 | chr8:60062081G>C | 3 months post-transplant | 86 | 17 | 12676 | 0.134% | 93 | 0 | 7674 | 0.000% |
| AML 248 | chr9:132157050C>T | 3 months post-transplant | 86 | 19 | 14354 | 0.132% | 93 | 0 | 7673 | 0.000% |
| AML 248 | chrX:17613357G>A | 3 months post-transplant | 86 | 1 | 9019 | 0.011% | 93 | 0 | 4637 | 0.000% |
| AML 251 | chr1:155904480T>A | 3 months post-transplant | 87 | 1 | 33055 | 0.003% | 81 | 0 | 4945 | 0.000% |
| AML 251 | chr1:179669423G>C | 3 months post-transplant | 87 | 2 | 41948 | 0.005% | 81 | 0 | 5275 | 0.000% |
| AML 251 | chr1:223462447G>C | 3 months post-transplant | 87 | 2 | 30328 | 0.007% | 81 | 0 | 4764 | 0.000% |
| AML 251 | chr1:75025278C>G | 3 months post-transplant | 87 | 3 | 29815 | 0.010% | 81 | 0 | 3176 | 0.000% |
| AML 251 | chr10:106665258G>C | 3 months post-transplant | 87 | 2 | 42435 | 0.005% | 81 | 0 | 4075 | 0.000% |
| AML 251 | chr10:36621510G>C | 3 months post-transplant | 87 | 1 | 30853 | 0.003% | 81 | 0 | 3412 | 0.000% |
| AML 251 | chr11:123643117C>G | 3 months post-transplant | 87 | 1 | 33854 | 0.003% | 81 | 0 | 5771 | 0.000% |
| AML 251 | chr11:5515469G>C | 3 months post-transplant | 87 | 0 | 41721 | 0.000% | 81 | 1 | 5416 | 0.018% |
| AML 251 | chr11:81220644G>A | 3 months post-transplant | 87 | 4 | 30807 | 0.013% | 81 | 0 | 4254 | 0.000% |
| AML 251 | chr11:99020392T>C | 3 months post-transplant | 87 | 1 | 34651 | 0.003% | 81 | 0 | 4423 | 0.000% |
| AML 251 | chr12:115262331G> | 3 months post-transplant | 87 | 1 | 44373 | 0.002% | 81 | 0 | 4967 | 0.000% |
| AML 251 | chr12:49052371T>C | 3 months post-transplant | 87 | 7 | 37299 | 0.019% | 81 | 0 | 6322 | 0.000% |
| AML 251 | chr12:85898023T>C | 3 months post-transplant | 87 | 3 | 31220 | 0.010% | 81 | 0 | 4217 | 0.000% |
| AML 251 | chr12:93574960>GA | 3 months post-transplant | 87 | 2 | 36510 | 0.005% | 81 | 0 | 4330 | 0.000% |
| AML 251 | chr12:97751063G>A | 3 months post-transplant | 87 | 3 | 30411 | 0.010% | 81 | 0 | 4106 | 0.000% |
| AML 251 | chr13:65393072G>T | 3 months post-transplant | 87 | 1 | 31220 | 0.003% | 81 | 0 | 3586 | 0.000% |
| AML 251 | chr13:85906764G>T | 3 months post-transplant | 87 | 3 | 33022 | 0.009% | 81 | 0 | 3279 | 0.000% |
| AML 251 | chr13:96373712C>T | 3 months post-transplant | 87 | 2 | 32389 | 0.006% | 81 | 0 | 3827 | 0.000% |
| AML 251 | chr14:29615725G>C | 3 months post-transplant | 87 | 1 | 32112 | 0.003% | 81 | 0 | 3560 | 0.000% |
| AML 251 | chr15:101389694T>C | 3 months post-transplant | 87 | 1 | 40168 | 0.002% | 81 | 0 | 6074 | 0.000% |
| AML 251 | chr17:30203376A>G | 3 months post-transplant | 87 | 1 | 34021 | 0.003% | 81 | 0 | 4725 | 0.000% |
| AML 251 | chr18:54001774G>C | 3 months post-transplant | 87 | 2 | 40419 | 0.005% | 81 | 0 | 4731 | 0.000% |
| AML 251 | chr18:60227705C>T | 3 months post-transplant | 87 | 2 | 35395 | 0.006% | 81 | 0 | 4053 | 0.000% |
| AML 251 | chr18:70524437A>T | 3 months post-transplant | 87 | 1 | 42581 | 0.002% | 81 | 0 | 4599 | 0.000% |

| | | | | | | | | | | |
|---|---|---|---|---|---|---|---|---|---|---|
| AML 251 | chr19:33623896C>A | 3 months post-transplant | 87 | 1 | 36554 | 0.003% | 81 | 0 | 5847 | 0.000% |
| AML 251 | chr2:212491338A>G | 3 months post-transplant | 87 | 3 | 30521 | 0.010% | 81 | 0 | 3172 | 0.000% |
| AML 251 | chr2:239529022C>T | 3 months post-transplant | 87 | 1 | 37809 | 0.003% | 81 | 0 | 6012 | 0.000% |
| AML 251 | chr2:30056856G>A | 3 months post-transplant | 87 | 1 | 38941 | 0.003% | 81 | 0 | 4313 | 0.000% |
| AML 251 | chr21:46412475C>T | 3 months post-transplant | 87 | 3 | 36178 | 0.008% | 81 | 0 | 5949 | 0.000% |
| AML 251 | chr3:181814957G>A | 3 months post-transplant | 87 | 1 | 32875 | 0.003% | 81 | 0 | 4103 | 0.000% |
| AML 251 | chr3:193785856G>C | 3 months post-transplant | 87 | 2 | 37546 | 0.005% | 81 | 0 | 5810 | 0.000% |
| AML 251 | chr3:48782120T>G | 3 months post-transplant | 87 | 2 | 34866 | 0.006% | 81 | 0 | 4759 | 0.000% |
| AML 251 | chr4:4944193G>T | 3 months post-transplant | 87 | 2 | 31513 | 0.006% | 81 | 1 | 6202 | 0.016% |
| AML 251 | chr5:9783059G>A | 3 months post-transplant | 87 | 1 | 37040 | 0.003% | 81 | 0 | 4917 | 0.000% |
| AML 251 | chr6:163301683T>C | 3 months post-transplant | 87 | 2 | 40580 | 0.005% | 81 | 0 | 4989 | 0.000% |
| AML 251 | chr6:163336041>GGGGCA | 3 months post-transplant | 87 | 3 | 34762 | 0.009% | 81 | 0 | 5511 | 0.000% |
| AML 251 | chr6:68956057G>C | 3 months post-transplant | 87 | 1 | 38515 | 0.003% | 81 | 0 | 4790 | 0.000% |
| AML 251 | chr7:100561558C>G | 3 months post-transplant | 87 | 1 | 44329 | 0.002% | 81 | 0 | 6333 | 0.000% |
| AML 251 | chr7:117041084G>C | 3 months post-transplant | 87 | 1 | 28745 | 0.003% | 81 | 0 | 3183 | 0.000% |
| AML 251 | chr7:147509396T>C | 3 months post-transplant | 87 | 2 | 36020 | 0.006% | 81 | 0 | 3954 | 0.000% |
| AML 251 | chr7:47605953>TGCCC | 3 months post-transplant | 87 | 3 | 44147 | 0.007% | 81 | 0 | 5598 | 0.000% |
| AML 251 | chr8:102015064T>C | 3 months post-transplant | 87 | 3 | 42190 | 0.007% | 81 | 0 | 4695 | 0.000% |
| AML 251 | chr8:14238270C>A | 3 months post-transplant | 87 | 4 | 31014 | 0.013% | 81 | 0 | 3369 | 0.000% |
| AML 251 | chr9:137746715C>T | 3 months post-transplant | 87 | 2 | 28570 | 0.007% | 81 | 0 | 4556 | 0.000% |
| AML 251 | chr9:5231567T>A | 3 months post-transplant | 87 | 3 | 41691 | 0.007% | 81 | 0 | 6388 | 0.000% |
| AML 251 | chrX:29072621>CCCC | 3 months post-transplant | 87 | 2 | 20966 | 0.010% | 81 | 0 | 1832 | 0.000% |

*Mutation genomic coordinates refer to hg38*

**Table S4: Driver mutations identified.**

| Patient | Chromosome | Position | Base from | Base to | Gene and Amino Acid Change | Included in v96 panel? |
|---|---|---|---|---|---|---|
| AML 121 | chr9 | 5073770 | G | T | JAK2 p.V617F | No |
| AML 122 | chr2 | 208248388 | C | T | IDH1 p.R132H | Yes |
| AML 122 | chr1 | 114716123 | C | T | NRAS p.G13D | Yes |
| AML 122 | chr13 | 28028192 | G | A | FLT3 p.A680V | No |
| AML 123 | chr15 | 90088702 | C | T | IDH2 p.R140Q | Yes |
| AML 123 | chr5 | 171410539 | - | TCTG | NPM1 p.W288fs | Yes |
| AML 123 | chr13 | 28034116 | - | GAGATCATATTCATATTCTCTGAAATCAACG | FLT3 ITD | No |
| AML 124 | chr17 | 31337858 | C | T | NF1 p.Q2207* | No |
| AML 124 | chr1 | 114713909 | G | T | NRAS p.Q61K | No |
| AML 124 | chr1 | 114716123 | C | T | NRAS p.G13D | No |
| AML 125 | chr5 | 171410539 | - | TCTG | NPM1 p.W288fs | Yes |
| AML 126 | chr2 | 25234373 | C | T | DNMT3A p.R882H | Yes |
| AML 127 | chr5 | 171410541 | - | TGCA | NPM1 p.W288fs | Yes |
| AML 127 | chr13 | 28034125, 28034132 | - | TCATATTCTCTGAAATCAACGTAGAAGTACTCATTATC<br>TGAGGAGCCGGTCACCTG, TCTCTGAAATCAACGTAG | FLT3 ITD | No |
| AML 128 | chr2 | 25245303 | - | A | DNMT3A p.V502f | No |
| AML 128 | chr2 | 208248389 | G | A | IDH1 p.R132C | Yes |
| AML 128 | chr13 | 28034126 | - | TCATATTCTCTGAAATCAACGTAG | FLT3 ITD | No |
| AML 129 | chr21 | 43094670 | C | T | U2AF1 p.R156H | No |
| AML 129 | chr13 | 28034100 | - | TTGGAAACTCCCATTTGAGATCATGGGGGTCCATCT | FLT3 ITD | No |
| AML 130 | chr3 | 128483911 | - | TA | GATA2 p.H323fs | Yes |
| AML 131 | chr9 | 5073770 | G | T | JAK2 p.V617F | No |
| AML 132 | chr17 | 7676011 | T | C | TP53 p.K120E | Yes |
| AML 133 | chr21 | 34880580 | C | T | RUNX1 p.R162K | Yes |
| AML 133 | chr20 | 32434981 | CAGCCATG | - | ASXL1 p.Q757fs | Yes |
| AML 133 | chr2 | 197402981 | C | T | SF3B1 p.E592K | Yes |
| AML 133 | chr1 | 114713908 | T | C | NRAS p.Q61R | No |
| AML 134 | chr1 | 43339489 | T | C | MPL p.S204P | Yes |
| AML 235 | chr4 | 54733154 | G | C | KIT p.D816H | No |
| AML 235 | chr4 | 54733155 | A | T | KIT p.D816V | No |
| AML 235 | chr11 | 118519716 | - | GG | KMT2A p.F3750fs | No |
| AML 236 | chr17 | 7675161 | G | A | TP53 p.P151S | No |
| AML 237 | chr5 | 171410539 | - | TCTG | NPM1 p.W288fs | No |
| AML 237 | chr15 | 90088702 | C | T | IDH2 p.R140Q | No |
| AML 238 | chr1 | 114713908 | T | C | NRAS p.Q61R | No |
| AML 239 | chr2 | 25234373 | C | T | DNMT3A p.R882H | No |
| AML 239 | chr5 | 171410539 | - | TCTG | NPM1 p.W288fs | No |
| AML 239 | chr13 | 28034099 | - | TGGAAACTCCCATTTGAGATCATATTCATATTCTCTGA<br>AATCAACGTAGAAGTACTCAT | FLT3 ITD | No |
| AML 240 | chr1 | 114716126 | C | T | NRAS p.G12D | Yes |
| AML 240 | chr5 | 171410540 | - | CTGC | NPM1 p.W288fs | No |
| AML 241 | chr11 | 108329172 | A | C | ATM p.Q2414P | No |
| AML 242 | chr1 | 114716123 | C | T | NRAS p.G13D | No |
| AML 242 | chr1 | 114716124 | C | G | NRAS p.G13R | No |
| AML 242 | chr17 | 76736877 | G | T | SRSF2 p.P95H | No |
| AML 242 | chr20 | 32434638 | - | G | ASXL1 p.G646fs | No |
| AML 242 | chr21 | 34880642 | - | GAGAAATAAAGAAGAGTTT | RUNX1 p.A142fs | No |
| AML 242 | chrX | 40057168 | G | A | BCOR p.Q1494* | No |
| AML 242 | chrX | 124066193 | - | T | STAG2 p.W706S | No |
| AML 244 | chr5 | 171410541 | - | TGCA | NPM1 p.W288fs | No |
| AML 244 | chr12 | 25245347 | C | T | KRAS p.G13D | Yes |
| AML 244 | chr13 | 28018486 | T | A | FLT3 p.N841I | No |
| AML 245 | chr5 | 171410539 | - | TCTG | NPM1 p.W288fs | No |
| AML 245 | chr11 | 32396398 | G | - | WT1 p.R370fs | Yes |
| AML 245 | chr13 | 28034090 | - | AGTGGCGGAATTTTCTCTTGGAAACTCCCATTTGAGAT<br>CATATTCATATTCTCTGAAATCAACGTAGAAGTACTCA<br>TTATCTGAAGTGGCGG | FLT3 ITD | No |
| AML 245 | chr13 | 28034143 | AA | TG | FLT3 p.V592A | No |
| AML 246 | chr17 | 7675124 | T | C | TP53 p.Y163C | Yes |
| AML 247 | chr1 | 114713907 | T | G | NRAS p.Q61H | No |
| AML 247 | chr1 | 114713908 | T | A | NRAS p.Q61L | No |
| AML 247 | chr1 | 114716123 | C | T | NRAS p.G13D | Yes |
| AML 247 | chr1 | 114716126 | C | G | NRAS p.G12A | No |
| AML 247 | chr12 | 25245351 | C | A | KRAS p.G12C | No |
| AML 247 | chr13 | 28018505 | C | A | FLT3 p.D835Y | No |
| AML 248 | chr4 | 105272684 | C | T | TET2 p.Q1435* | No |
| AML 249 | chr2 | 197402635 | C | G | SF3B1 p.K666N | No |
| AML 249 | chr17 | 76736877 | G | A | SRSF2 p.P95L | No |
| AML 249 | chr21 | 34880580 | C | T | RUNX1 p.R162K | No |

| | | | | | | |
|---|---|---|---|---|---|---|
| AML 249 | chr21 | 34886952 | - | C | RUNX1 p.V81fs | No |
| AML 250 | chr5 | 171410539 | - | TCTG | NPM1 p.W288fs | No |
| AML 250 | chr13 | 28034123 | - | TATTCATATTCTCTGAAATCA | FLT3 ITD | No |
| AML 250 | chr15 | 90088702 | C | A | IDH2 p.R140L | No |
| AML 251 | chr11 | 118490196 | A | G | KMT2A Pp.Q1548R | No |
| AML 251 | chr12 | 49052371 | T | C | KMT2D p.M438V | Yes |
| AML 251 | chr15 | 90088702 | C | T | IDH2 p.R140Q | No |

*Mutation genomic coordinates refer to hg38*
